# Uptake of infant and pre-school immunisations in Scotland and England during the COVID-19 pandemic: an observational study of routinely collected data

**DOI:** 10.1101/2021.07.19.21260770

**Authors:** Fiona McQuaid, Rachel Mulholland, Yuma Sangpang Rai, Utkarsh Agrawal, Helen Bedford, J. Claire Cameron, Cheryl Gibbons, Partho Roy, Aziz Sheikh, Ting Shi, Colin R Simpson, Judith Tait, Elise Tessier, Steve Turner, Jaime Villacampa Ortega, Joanne White, Rachael Wood

## Abstract

**Background:** In 2020, the COVID-19 pandemic and control measures such as national lockdowns threatened to disrupt routine childhood immunisation programmes. Initial reports from the early weeks of lockdown in the UK and worldwide suggested that uptake could fall putting children at risk from multiple other infectious diseases. In Scotland and England, enhanced surveillance of national data for childhood immunisations was established to inform and rapidly assess the impact of the pandemic on infant and preschool immunisation uptake rates.

**Methods and findings:** We undertook an observational study using routinely collected data for the year prior to the pandemic (2019), and immediately before, during and after the first period of the UK ‘lockdown’ in 2020. Data were obtained for Scotland from the Public Health Scotland “COVID19 wider impacts on the health care system” dashboard (https://scotland.shinyapps.io/phs-covid-wider-impact/) and for England from ImmForm.

Five vaccinations delivered at different ages were evaluated; three doses of the ‘6-in-1’ DTaP/IPV/Hib/HepB vaccine and two doses of MMR. Uptake in the periods in 2020 compared to that in the baseline year of 2019 using binary logistic regression analysis. For Scotland, we analysed timely uptake of immunisations, defined as uptake within four weeks of the child becoming eligible by age for each immunisation and data were also analysed by geographical region and indices of deprivation. For both Scotland and England, we assessed whether immunisations were up to date at approximately 6 months (all doses 6-in-1) and 16-18 months (first MMR) of age.

We found that uptake rates within four weeks of eligibility in Scotland for all the five vaccine visits were higher during the 2020 lockdown period than in 2019. The difference ranged from 1.3% for the first dose of the 6-in-1 vaccine (95.3 vs 94%, OR 1.28, CI 1.18-1.39) to 14.3% for the second MMR dose (66.1 vs 51.8 %, OR 1.8, CI 1.74-1.87). Significant increases in uptake were seen across all deprivation levels, though, for MMR, there was evidence of greater improvement for children living in the least deprived areas.

In England, fewer children who had been due to receive their immunisations during the lockdown period were up to date at 6 months (6-in-1) or 18 months (first dose MMR). The fall in percentage uptake ranged from 0.5% for first 6-in1 (95.8 vs 96.3%, OR 0.89, CI 0.86-0.91) to 2.1% for third 6-in-1 (86.6 vs 88.7%, OR 0.82, CI 0.81-0.83).

**Conclusions:** This study suggests that the national lockdown in Scotland was associated with a positive effect on timely childhood immunisation uptake, however in England a lower percentage of children were up to date at 6 and 18 months. Reason for the improve uptake in Scotland may include active measures taken to promote immunisation at local and national level during this period. Promoting immunisation uptake and addressing potential vaccine hesitancy is particularly important given the ongoing pandemic and COVID-19 vaccination campaigns.

## Introduction

The COVID-19 pandemic and associated control measures such as national ‘lockdowns’, involving varying restrictions on leaving the home, work and socialising, have had a profound impact on daily life and the delivery of healthcare worldwide. In the UK, a national lockdown was announced on the 23^rd^ March 2020 with instructions that people should only leave their home for a limited number of “essential” reasons. (1) This was accompanied by the reconfiguration of acute healthcare services to support the anticipated influx of COVID-19 patients, cancellation of most elective activity and pausing of screening programmes. (2) During lockdown, there was evidence of a change in healthcare-seeking behaviour – for example, in Scotland the uptake of both emergency and elective hospital based care dropped substantially over the lockdown period. (3) However, within child health, key routine services including childhood immunisations continued through the UK. (4)

It has become increasingly apparent that younger children are at low risk of severe disease due to Severe acute respiratory syndrome coronavirus 2 (SARS-CoV-2) (5) and may be less susceptible to infection with the virus. (6) Yet the wider impact of the pandemic on children in terms of education, mental and physical health and safeguarding is not yet fully understood and is likely to be profound. (7, 8) One particular area of concern early in the lockdown period was the potential effect on the uptake of routine childhood immunisations. (9) Maintaining high population vaccine coverage is vital for both direct and indirect (via herd immunity) protection against non-COVID-19 infectious diseases.

In July 2020, the World Health Organization (WHO) warned of a potential decline in routine immunisation rates associated with the COVID-19 pandemic, citing a poll from May 2020 in which respondents from 82 countries suggested disruption to immunisation programmes was widespread (10). Initial reports from England, (11) Pakistan (12) South Africa, (13) Singapore (14) and the USA, (15) were concerning, suggesting a fall in children receiving their scheduled vaccinations in the very early weeks of national lockdowns, though the full impact has not yet been assessed. However, the English and US studies relied on surrogate measures of vaccine uptake; number of vaccines delivered/ordered (without a corresponding denominator) (11, 15) and the Singaporean study used convenience sampling with multiple assumptions for missing data (14). Longer-term data were available from the KwaZulu-Natal province of South Africa (13) and Sindh province of Pakistan (12), which appeared to show some recovery after an initial fall in uptake; however, in Pakistan, part of the lockdown restrictions involved shutting down of outreach immunisation programme. The overall impact of lockdown on immunisation uptake in higher income countries which maintained their routine immunisation programmes is unclear.

Given the prolonged, and repeated periods of lockdown, it is important to evaluate the overall effect on childhood immunisation uptake. The aim of this study was therefore to provide a longer perspective than previous studies by describing the pattern of childhood immunisation uptake in Scotland and England before, during and immediately after the first national lockdown implemented in response to the pandemic (23^rd^ March-31st July), with comparisons to baseline data from 2019, by geographical area and socio-economic deprivation.

## Methods

### Study design

This observational study took advantage of the natural experiment afforded by the COVID-19 pandemic and used routinely collected data for the year prior to the pandemic (2019) and immediately before, during and after the first period of ‘lockdown’ imposed by the United Kingdom and Scottish governments in 2020. Data were available for Scotland and England, however, as discussed below, variations in time points at which the data were collected precluded direct comparisons. Of note, this analysis relates to the first national lockdown which began on 23^rd^ March 2020 with restrictions easing gradually from over June and July 2020.

The vaccines chosen as indicators of preschool immunisation uptake were the hexavalent DTaP/IPV/Hib/HepB vaccine, (referred to here as ‘6in1’), which protects against Diphtheria, Tetanus, Pertussis, Polio, *Haemophilus influenzae* type b and Hepatitis B, and the Measles, Mumps and Rubella vaccine (referred to as ‘MMR’). In the UK, the 6in1 vaccine is recommended at age eight (‘first dose 6in1’), 12 (‘second dose 6in1’) and 16 (‘third dose 6in1’) weeks of age, and MMR is given at 12 months (‘first dose MMR’) and 3 years 4 months (‘second dose MMR’) (16). Uptake of the additional immunisations offered at the same ages (Meningococcal C, Rotavirus and Pneumococcal) was not directly measured.

For Scotland, we chose to primarily examine uptake within four weeks of eligibility as this represents timely uptake of vaccinations as per the recommended UK schedule (16) leading to the child being protected at the earliest recommended opportunity. All children living in Scotland who became eligible by age for any of the pre-school immunisations of interest from January 2019 up to and including the week beginning 28^th^ September 2020 were included. Of secondary interest, and to allow descriptive comparisons with data from England, we also analysed uptake at approximately 6 months (range 24-32 weeks) for the 6in1 and 16 months for the first dose MMR. For England, equivalent data on uptake within four weeks were not available therefore the analysis was conducted on uptake by six (6in1) or 18 months of age (first MMR) and monthly, rather than weekly, data were used.

Vaccine uptake was analysed in the following four time periods: 1^st^ January to 31^st^ December 2019 (“2019”), 1st January to week beginning 16th March 2020 (“pre-lockdown”), 23rd March to week beginning 27^th^ July (“lockdown”) and week beginning 3^rd^ August to week beginning 28^th^ September 2020 inclusive (“post-lockdown”). These time periods were chosen to correspond with the beginning of the first UK-wide lockdown as announced by the UK government on the 23rd March 2020 (1). The end of the lockdown period is less well-defined and varied both in approach and timescale between Scotland and England (17, 18). Broadly speaking, by the end of July 2020, there was a substantial reduction in ‘lockdown’ restrictions in both countries with the opening of many non-essential businesses and limited indoor meeting between households permitted, therefore a pragmatic approach was taken to define the lockdown period as 23rd March 2020 until 31st July 2020. Children were included in the time period at which they first became eligible by age. As data from England were available by month only, the pre-lockdown period included January to end of March 2020.

### Data sources

The Scottish data used in this paper were extracted in January 2021 from the PHS “COVID19 wider impacts on the health care system” dashboard, which is publicly accessible at https://scotland.shinyapps.io/phs-covid-wider-impact/. The dashboard includes aggregate information on immunisation uptake, including by the geographical area in which the child is living and the corresponding Scottish Index of Multiple Deprivation (SIMD) (both assigned based on the child’s postcode registered on the Scottish national vaccination call-recall system, SIRS). The SIMD breaks Scotland into 6976 small areas of similar population size and assigns one of five SIMD quintile based on indicators of deprivation including income, education and housing, with 1 representing the most deprived areas and 5, the least. (19) We defined geographical areas by the Health and Social Care Partnership (HSCP) in which the child lives. Within Scotland there are 31 HSCPs which provide integrated health and social care to their population.

English data were extracted from the ImmForm system, for January 2019 (representing the first extract for pre-lockdown period) to September 2020 (representing the final extract for post-lockdown period), which automatically receives monthly aggregate data on vaccine uptake from 92-95% of English GP practices, provided by GP System Suppliers. These data are validated and analysed by Public Health England to check completeness, query any anomalous results, and are used to describe epidemiological trends, as well as being used directly locally by the NHS for performance management. Data were available for immunisation uptake at age six months (each dose of the 6in1immunisation) and 18 months (first MMR) only.

### Statistical methods

The primary outcome examined was the change in percentage uptake, within four weeks of eligibility, of each of the immunisations of interest between baseline uptake rates in 2019 and during the lockdown period for Scotland as a whole and by geographical areas or level of deprivation. Of secondary interest were comparisons between 2019 and the other time points listed above. Broadly comparable analyses were conducted to examine the primary outcome for England as a whole. Due to differences in data collection methods and age of child at data extraction point, comparisons between Scottish and English data were descriptive only. The prespecified analysis plan is included within the supplementary materials.

To compare uptake rates between time periods, aggregate binary logistic regression modelling was conducted, using vaccination status (vaccinated vs unvaccinated) as the dependent variable, and time period as the explanatory variable.. Separate analyses were carried out for each vaccine and country. When comparing HSCP or deprivation, an additional interaction with HSPC or SIMD quintile was included in the model. Odds ratios with 95% confidence intervals were calculated for uptake in the period of interest compared to the 2019 baseline. Given the nature of the aggregated data available, no adjusting for potential confounders or effect modifiers was possible.

All analyses and generation of figures were performed using R/R Studio (4.0.3). All code is publicly available via the EAVE II GitHub page (https://github.com/EAVE-II) and data can be accessed via the PHS wider impacts dashboard https://scotland.shinyapps.io/phs-covid-wider-impact/. Aggregated English data could be made available if in agreement with information governance regulations and upon request from the authors. Further methodological details can be found in S2 File supplemental methods.

### Ethics and funding

Ethical approval for this specific study was not required as we have used publicly available, anonymised, aggregated data. Results have been reported using the STROBE (20) and RECORD (21) guidelines. No specific funding was received for this project, RM is funded by the Health Data Research UK BREATHE hub. ImmForm data are not publicly available, however, an agreement was accepted for England to only share the national figures and analyses outputs.

## Results

### Preschool immunisation uptake increased during the lockdown period in Scotland

Across Scotland, the percentage of preschool children receiving their immunisations within four weeks of becoming eligible increased during the lockdown period for all five immunisations (Fig1, Table 1, Table S1). The change in percentage uptake compared to the 2019 baseline ranged from 1.3% for the first dose 6in1 (OR 1.28, CI 1.18-1.39) to 14.3% or the second dose MMR (OR 1.8, CI 1.74-1.87) (Table 1). Across all the immunisations visits, this equated to an additional 7,508 infants/preschool children receiving their immunisations in a timely manner over the lockdown period compared to the baseline rates of 2019. Uptake rates dipped immediately before the announcement of a national lockdown in mid-March 2020 (Fig1), then peaked throughout June before starting to decrease. However, uptake remained significantly higher than 2019 during the post lockdown period (Fig 1, Table 1). Of note, prior to lockdown for both MMR doses, there was already a modest, increase in uptake compared to 2019 (Table 1).

**Fig 1:**
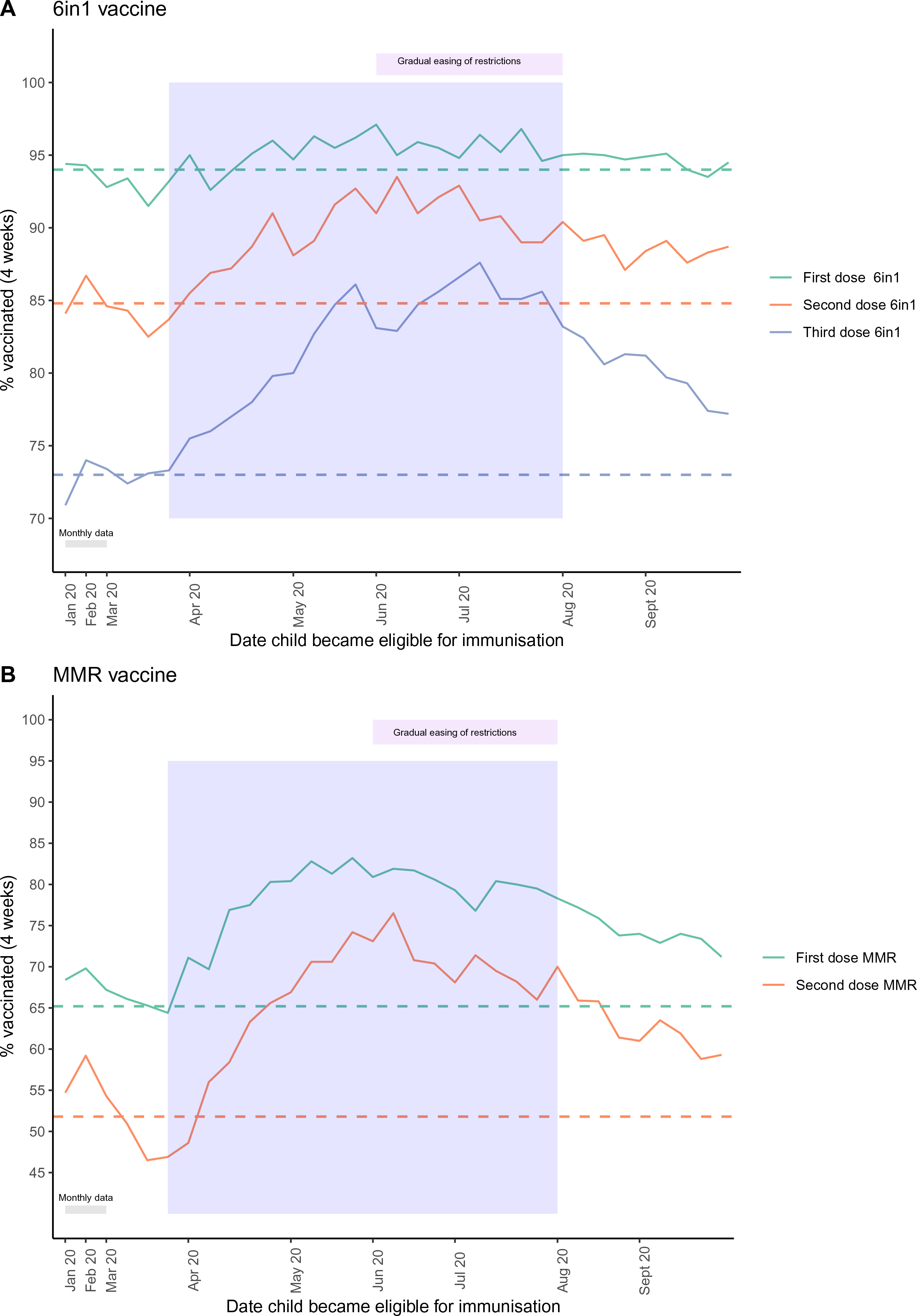
Percentage of children in Scotland immunised within 4 weeks of eligibility. (A) all doses 6in1 vaccine, (B) MMR vaccine across January to September 2020. Lockdown period = blue shaded area. For January and February, a single mean monthly value is plotted and from March onwards, weekly uptake is shown (see table S1 for full data). Dashed horizontal lines indicated the mean uptake in 2019 of the immunisation with the corresponding colour. The increase in uptake during the lockdown period was statistically significant (see Table 1 for details).

**Table 1:**
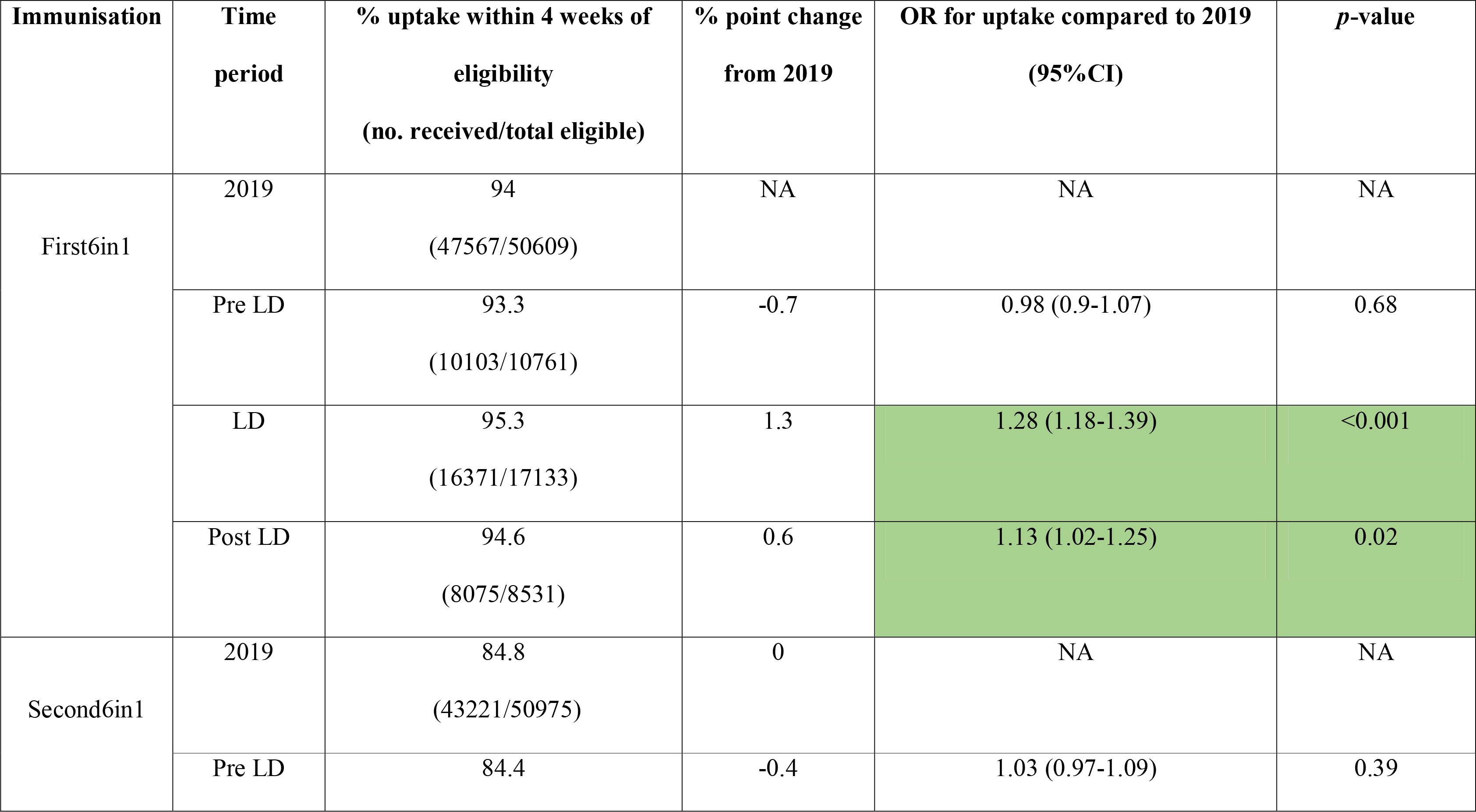
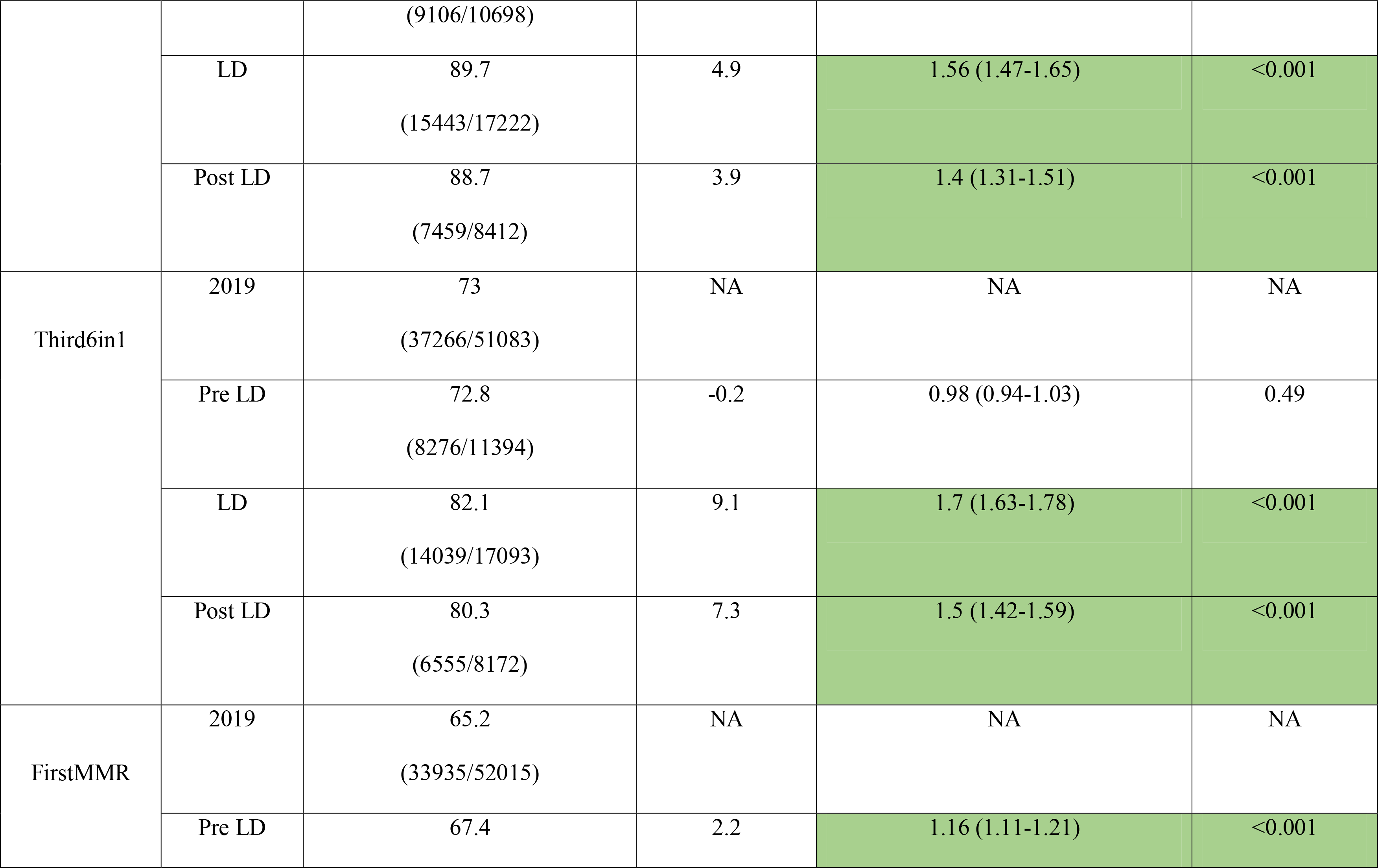
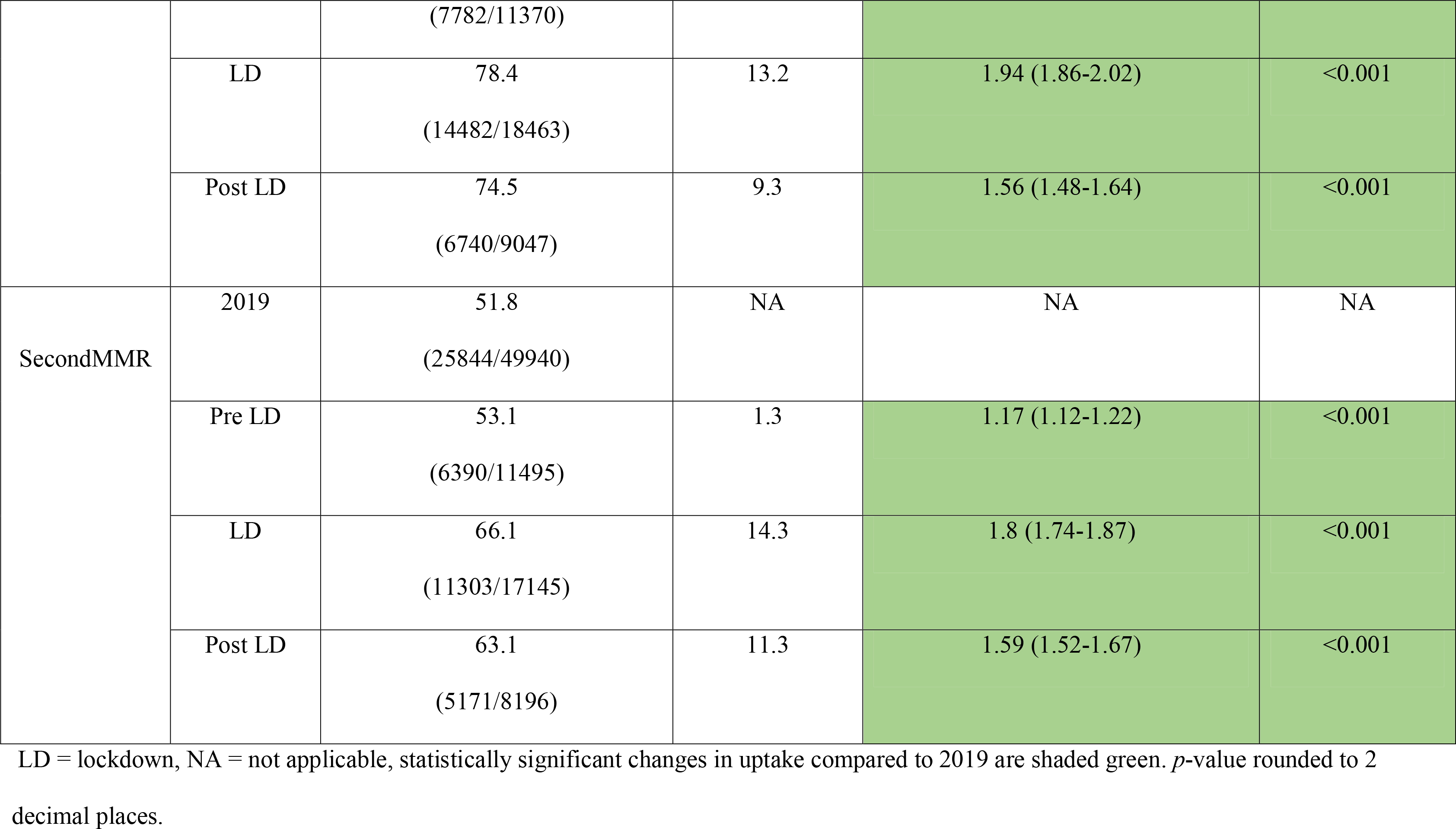
Uptake of pre-school immunisations by time-period with odds ratio compared to baseline of 2019.

### Variation in uptake of preschool immunisations by geographical area

Baseline data from 2019 showed the percentage uptake of preschool immunisations within four weeks of eligibility varied widely by geographical HSCP and immunisation (Fig2, FigS1, Table S2). In keeping with the rise in mean uptake across Scotland for all vaccines, the percentage of children immunised in most HSCP increased between 2019 and lockdown. However, not all followed this pattern with a minority demonstrating a fall in uptake (Fig2, FigS1, FigS2, Table S2). Care must be taken when interpreting percentage results from the island HSCPs (Shetland Islands, Orkney Islands and Western Isles) given the very small numbers of children involved (Table S2).

For individual HSCP, the statistical significance of the change in uptake varied by immunisation (Fig 2 and Table S2). Despite a general trend of improvement for the first 6in1 vaccine, we found a significant change for only eight of the 31 HSCP, mainly centred around the more densely populated, urban central belt of Scotland (Glasgow City, Edinburgh, Stirling and Clackmannanshire, East Dunbartonshire, Falkirk, Fife, South Lanarkshire, South Ayrshire, Fig 2). However, this pattern evolved with the different immunisation visits and with almost all HSCP showing a rise in uptake for both MMR immunisations, with percentage point increases as high as 30% (Angus, 74.1% vs 43.8%, OR 3.38, CI 2.61-4.37) (Fig 2, table S2).

**Fig 2:**
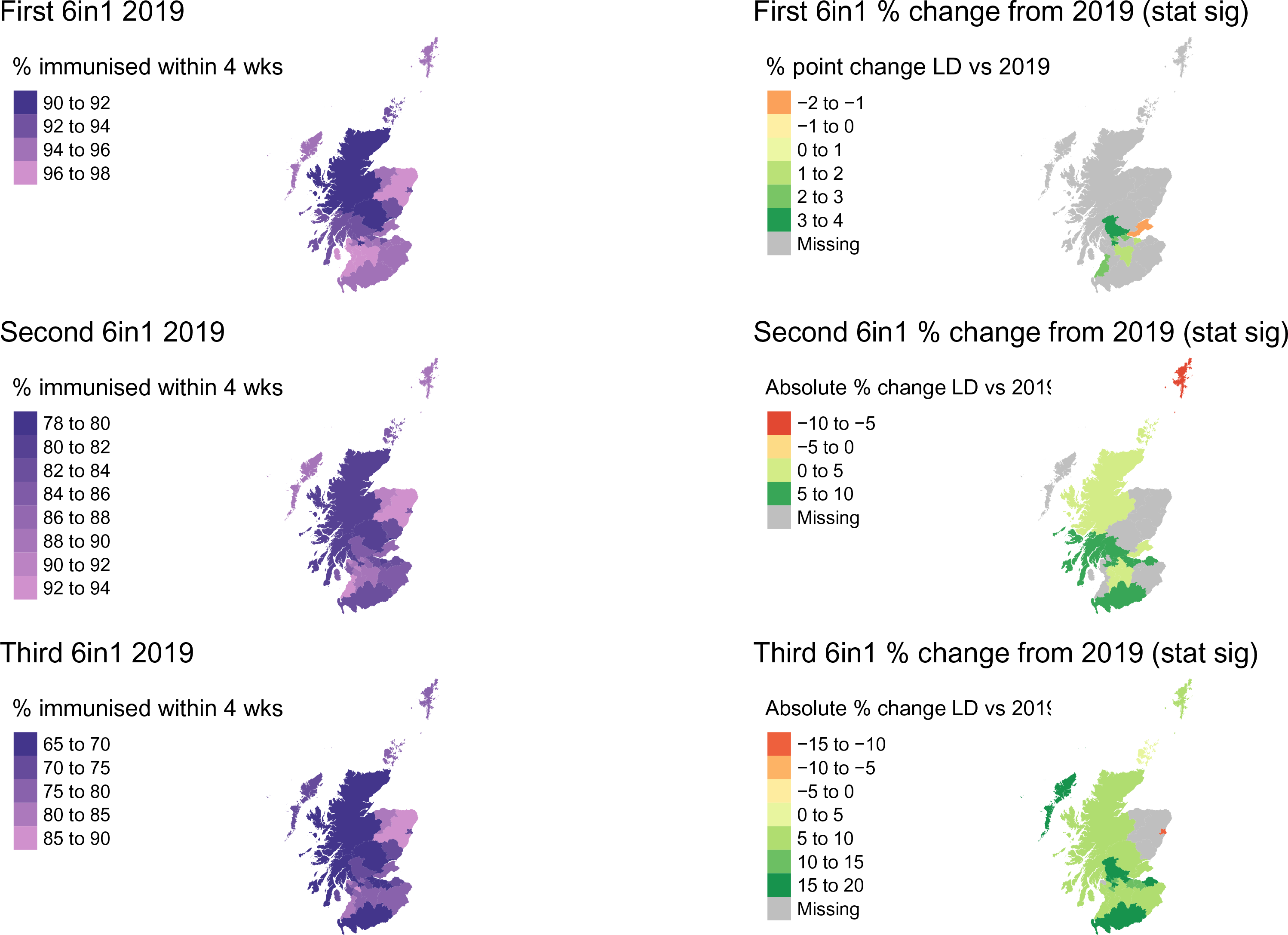
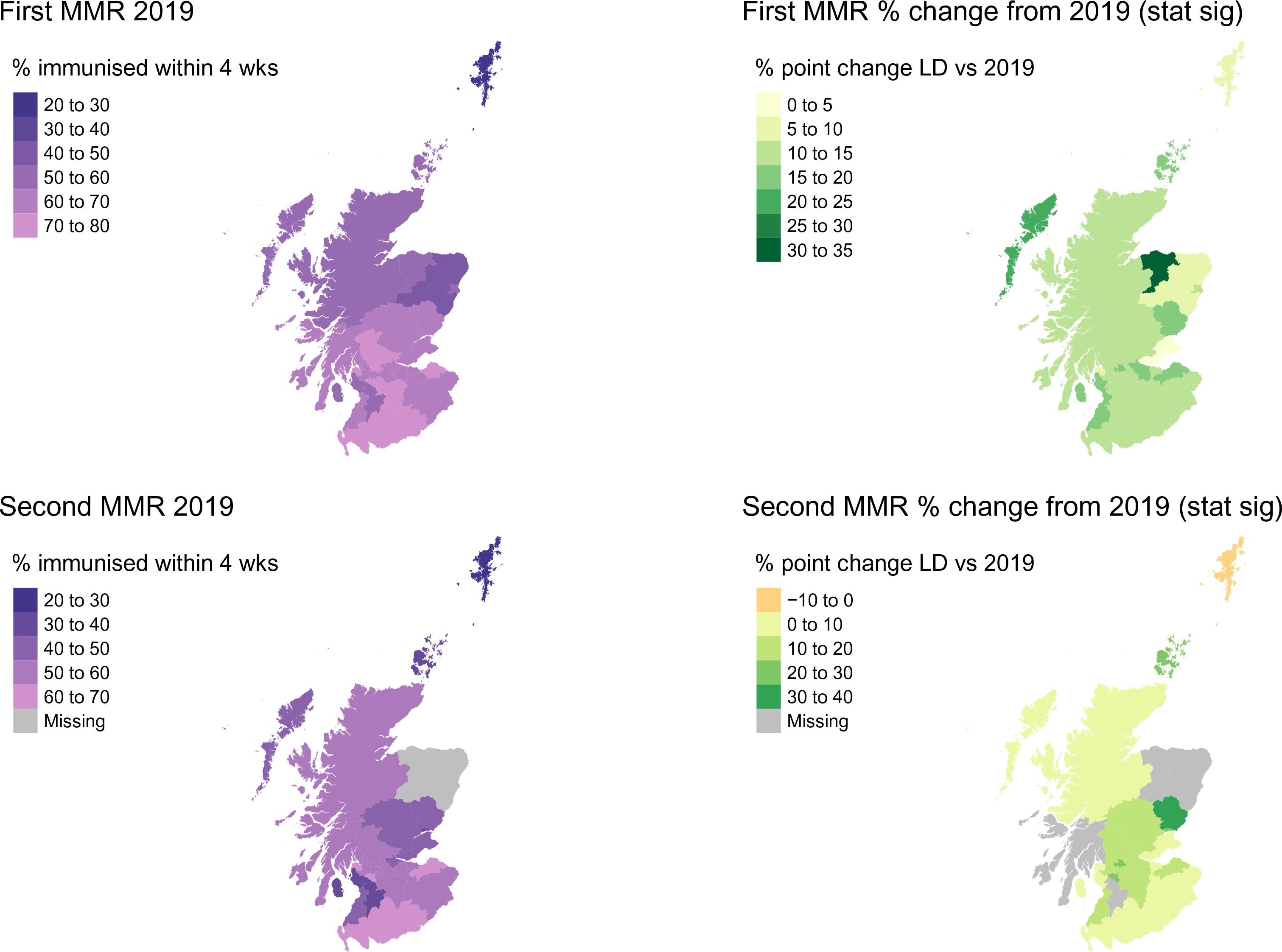
Choropleth maps showing baseline mean percentage uptake by HSCP. Left hand side = 2019, Right hand side = percentage point difference between 2019 and lockdown for areas in which the difference was statistically significant. Grey = no statistically significant changes in uptake between 2019 and lockdown, with the exception of Grampian HSCP for MMR dose 2 (see text).

### Preschool immunisation uptake increased across all deprivation levels

Percentage uptake within four weeks of becoming eligible rose across all SIMD quintiles, between 2019 and lockdown, for all immunisations (Fig3). The magnitude of this rise varied by quintile and vaccine (FigS3, Table S3) from 0.3% (SIMD 4, first 6in1 dose, OR 1.1, 95% CI 0.9-1.3) to 16.2% (SIMD 5, second MMR dose, OR 2.1, 95% CI 1.9-2.3). The increase in uptake between 2019 and lockdown was statistically significant for all except first dose 6in1 for the least deprived quintiles (4 and 5) (Table S3). In the post-lockdown period, percentage uptake remained significantly higher than the 2019 baseline for all quintiles for each vaccine except for the first 6in1 dose, in which only the most deprived quintile retained a significant increase (Fig S3, Table S3).

In keeping with previous observations (22), children in the least deprived quintiles were more likely to be immunised and this relationship was broadly maintained throughout the study period (Fig3 & FigS4). While all quintiles improved uptake between 2019 and lockdown, whether the inequality between most and least deprived increased or decreased varied by vaccine type. For all doses of the 6in1 vaccine there was a tendency to a convergent improvement i.e., the gap in percentage uptake between the quintiles lessened, while for both MMR doses there was further divergence in uptake rates between most and least deprived (Fig 3, FigS4, Table S4). The interaction of SIMD quintile and time period was non-significant for all 6in1 doses; that is to say all SIMD quintiles improved equally for this immunisation (Table S4). However, for the first MMR dose, the improvement in uptake was statistically greater for SIMD quintiles 3-5 compared to SIMD 1, and for the second MMR dose, SIMD 5 showed a significantly larger increase in uptake between 2019 and lockdown (Table S4). This suggests that for the MMR immunisation, the factors leading to an improvement in uptake had greater positive impact for children living in less deprived areas.

**Fig 3:**
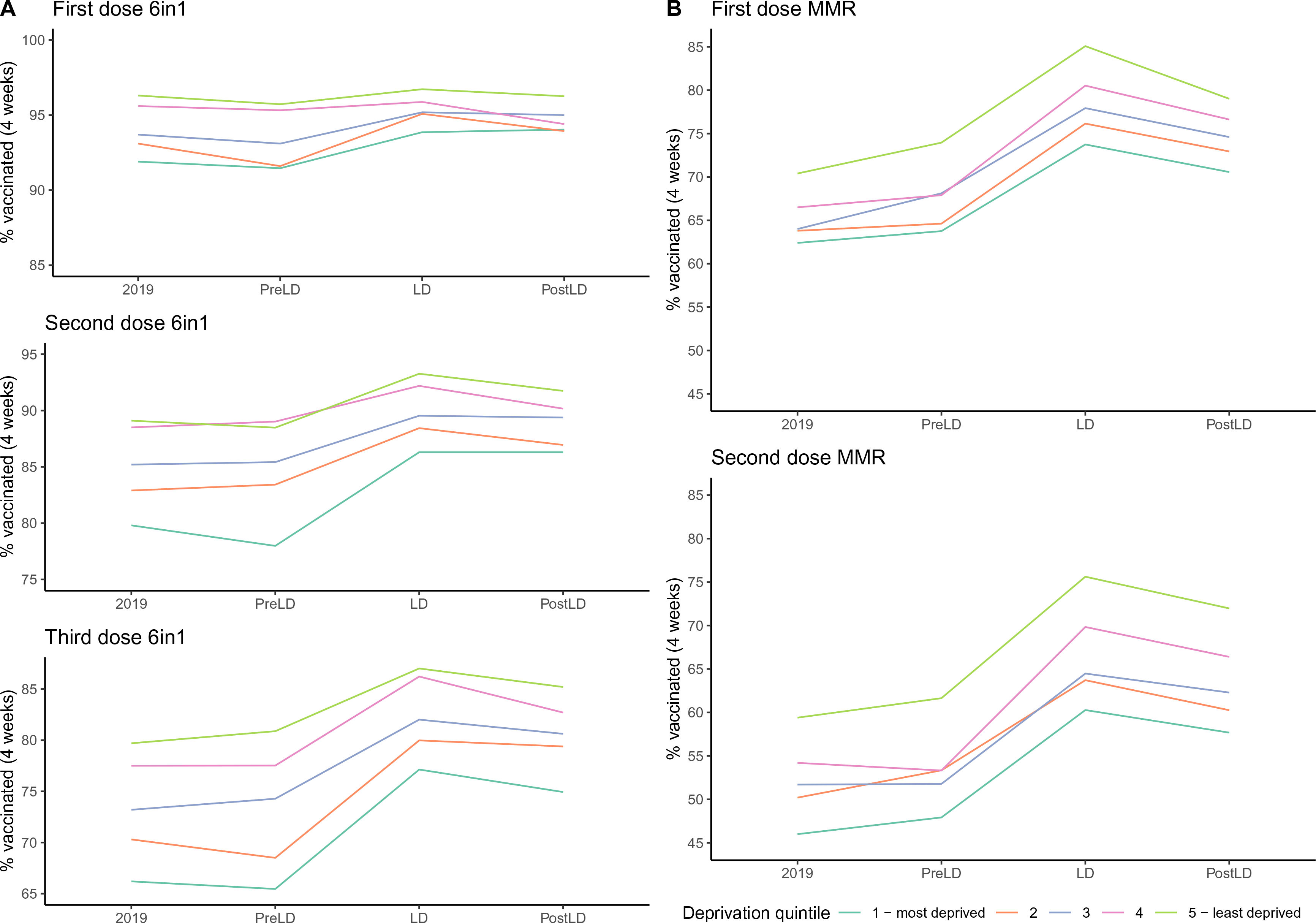
Mean percentage immunised by SIMD quintile. A= 6in1 vaccine, B= MMR vaccine. See Figure S3 and Table S3 for absolute percentage rise compared to 2019 and significance levels.

### ‘Catch up’ immunisation rates and comparison with data from England

Thus far, the Scottish data presented has related to children receiving their immunisations as per the recommended schedule (within four weeks of the child becoming eligible by age). representing a ‘gold standard’ in which the child is protected as early as possible. It is recognised that some children will receive their immunisations after this point. This ‘catch-up’ effect can be seen in uptakes rate for all Scottish children by the time they reached between six and eight months (6in1), 16 months (first MMR) or three years eight months (second MMR) (Fig 4A). Those becoming eligible during lockdown showed minimal change in this longer-term measure of uptake for the three doses of the 6in1 immunisation, a small increase in uptake of the first MMR, and a more substantial increase in uptake of the second MMR (Table S5). These data suggest that while lockdown was associated with a beneficial effect on timely uptake of all infant and pre-school immunisations, the impact on longer term or final achieved uptake was more variable, possibly reflecting a ceiling effect on maximal uptake, for the earliest immunisations.

**Fig 4:**
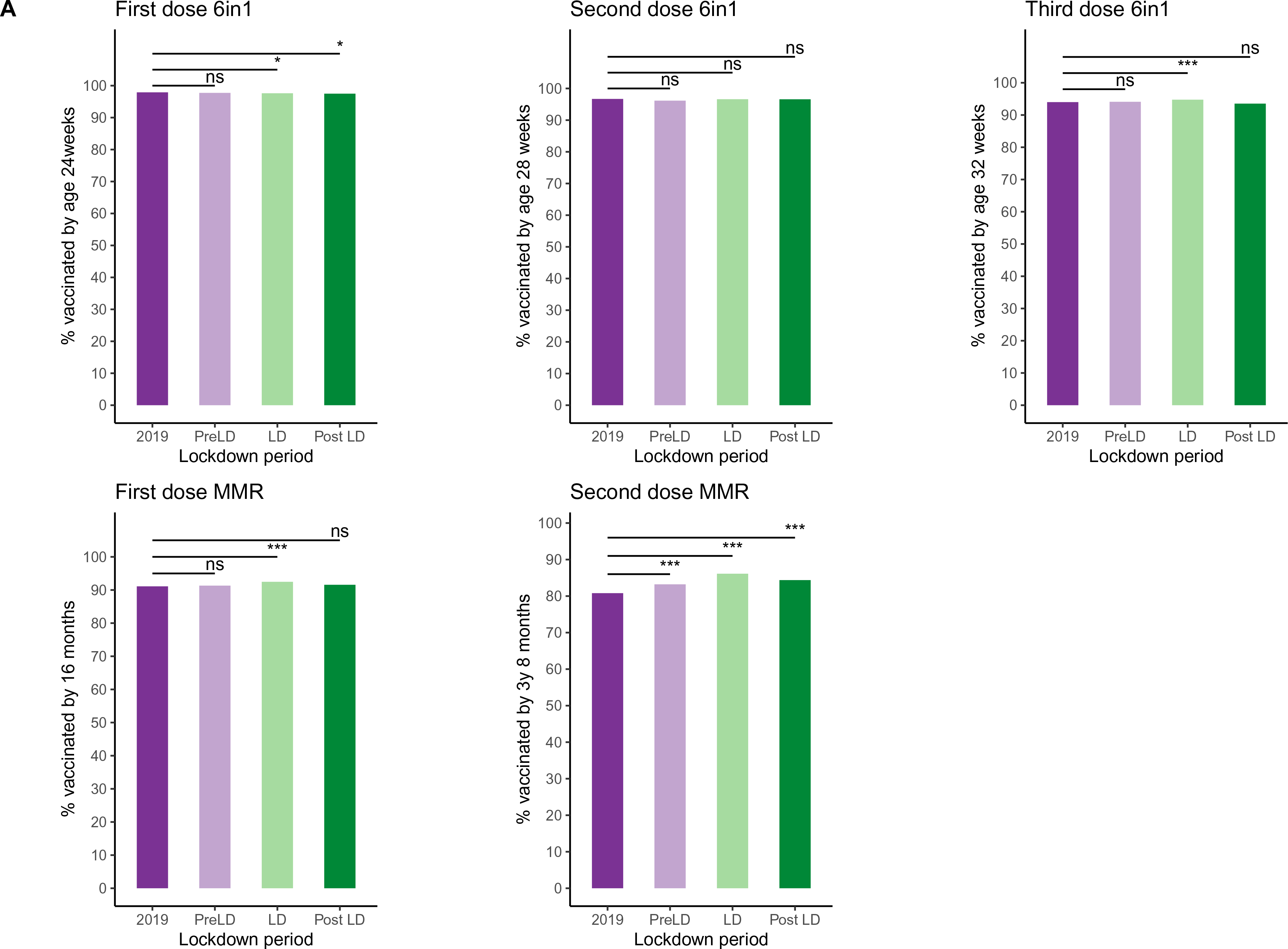
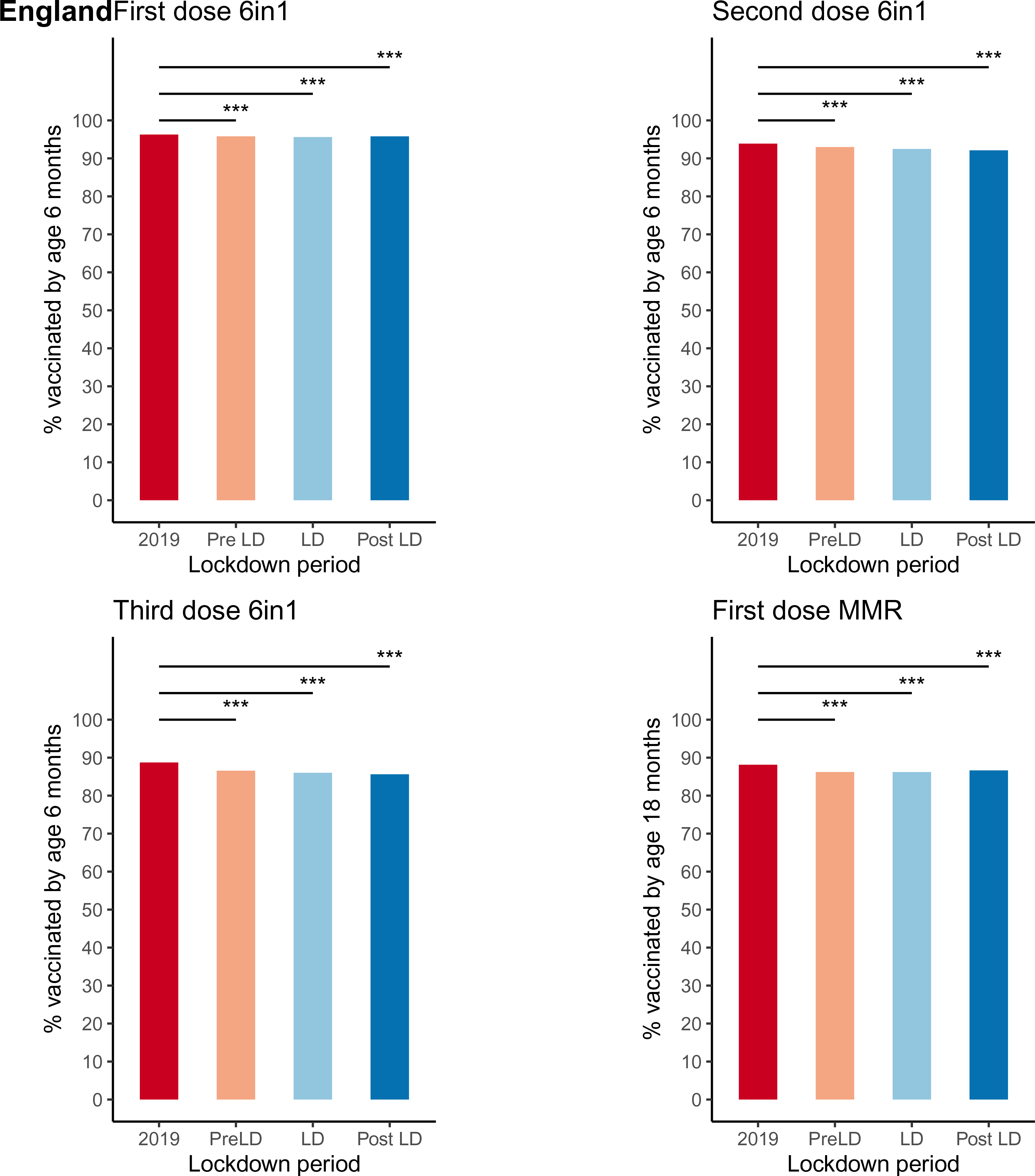
Percentage of children up to date at 6/18months. Overall mean percentage of children immunised by approximately 6 months of age (all doses of 6 in1, see y-axis for specific ages) or 16-18 months (first MMR) for Scotland and England. Each bar contains children who became eligible for the immunisation of interest during the time period indicated. A= Scotland, B = England *** *p*-value < 0.001, * *p*-value <0.05, ns = not significant

For England, broadly equivalent data were available for children aged six months (all doses 6in1) and 18 months (first dose MMR) who had become eligible for their immunisations during the time periods of interest (Fig 4B, Table S6). These data demonstrated a small, but statistically significant fall in uptake for all the immunisations in the lockdown periods compared to 2019, ranging from 0.5% (first dose 6in1, 95.8% vs 96.3%, OR 0.89, CI 0.86-0.91) to 2.1% (third dose 6in1, 86.6% vs 88.7, OR 0.82, CI 0.81-0.83) (Table S6). However, much of the fall in uptake took place in the pre-lockdown period, particularly for the third dose 6in1, with a gradual recovery seen over the lockdown period itself (FigS5). A general trend towards falling MMR uptake can also be seen to pre-date the COVID-19 pandemic (Fig S5).

## Discussion

Contrary to initial reports which focused only on the very early weeks of national lockdowns implemented in response to the COVID-19 pandemic, we found that early uptake of infant and preschool immunisations (within four weeks of a child becoming eligible) rose significantly for the duration of the first lockdown period in Scotland, resulting in thousands more children receiving their immunisations at the scheduled time. This is an important message to send to support public and professional confidence in the preschool immunisation programme and will help normalise timely immunisation uptake for both parents and health services. Improving public and professional confidence is particularly vital given in the current climate of promoting vaccination against SARS-CoV-2. Encouragingly, positive results were seen across all levels of deprivation, though some geographical variations were observed across Scotland. Improvement was also seen in longer term uptake of the first and second MMR immunisations (immunisation within four months of becoming eligible) in the lockdown period. Findings in England differed, with a small fall in longer term uptake of immunisations observed for the lockdown period.

### Strengths and limitations

It is important to acknowledge the limitations of the data presented, many of which arise from opportunistically using routinely collected data rather than that obtained from a specific study design. The SIRS electronic system is well-established and captures data on the entire child population in Scotland. However, the aggregate surveillance data derived from the system that we could access lacked detailed information on several potentially important factors, not least of which was ethnicity, which is known to affect both immunisation uptake and attitudes towards immunisation. (23) In the 2011 Scottish Census, 92% of the population of Scotland identified themselves as White Scottish/British, and only 4% as non-white, whereas in England, 81% described themselves as White British and 14% non-white. (24) It may not be appropriate to extrapolate these data to countries with a significantly different ethnic make-up and it is plausible that some of the difference seen between the Scottish and English data could be due to these factors.

In using the mean percentage of the entire year 2019 as our baseline comparator, we potentially run the risk of confusing normal seasonal variation in immunisation uptake with the impact of lockdown measures. Ideally, direct weekly or monthly comparisons would be made between 2019 and 2020, however quarterly trends published for previous years including 2019 do not show major difference in uptake throughout the year and in fact show a gradual decline in uptake year on year since 2015. (25) In addition, it is possible that 2020 uptake rates have been underestimated, due to a lag in data entry into SIRS which would particularly effect the ‘catch-up’ rates (Fig4A).

Therefore, caution must be taken not to over-interpret the results presented here or extrapolate to significant different populations with varying baseline immunisation uptakes rates and less robust immunisation programmes, the organisation of which may have been adversely affected by the pandemic. Nevertheless, this study has efficiently and quickly produced useful and valid results which have the potential to aid the development of future research and guide policy.

### Interpretation and implications for policy, practice and research

Despite these encouraging data, it is not possible to ascertain from the numbers alone which are the key contributing factors to improving uptake. This is a key avenue of future research as lessons learnt can then be taken forward to optimise future immunisation programmes, both within the pandemic setting and beyond. While the concept of vaccine hesitancy is a popular media topic, previous studies have shown that the reasons given by parents for not vaccinating their children are often much more practical. In fact, a pre-pandemic report by the Royal Society for Public Health in the UK found that the major barriers for parents who wanted to immunise their children were timing and availability of appointments and childcare duties. (26) Although for those who actively chose not to immunise their children, fear of side effects was a key concern and the negative effects of social media are increasing. (26) The lockdown has had a major effect on parental working patterns with almost 9 million UK employees furloughed and millions more working from home, often with the additional tasks of managing childcare as schools and nurseries were closed. (27) While this had made life significantly more challenging for many, more flexible working patterns may have made attending immunisation appointments easier for some.

Jarchow-MacDonald and colleagues (28) from NHS Lothian (which consists of HSCPs Edinburgh, West Lothian, East Lothian and Midlothian) have suggested that ensuring the accessibility of immunisation centres, either by public transport or by providing mobile services to shielding families, was important in maintaining uptake during the pandemic, as was directly communicating with families with a pre-appointment phone call and reminder postcards. This gave families an opportunity to discuss the immunisation with a healthcare professional, a strategy that has been showed to be important in addressing parental concerns. (29) In fact, the reminder alone may have been sufficient to encourage parents to attend the first appointment. (30) The clear commitment of the Scottish Government to maintain the immunisation programme was also felt to be an important factor. (28)

In England, Bell et al (31) conducted a large scale online survey of parents of children under the age of 18 months to assess their experiences of accessing immunisations during the early part of the UK lockdown (19^th^ April-11^th^ May 2020). They highlighted the uncertainty of some families about whether the immunisation service was continuing, particularly amongst non-white ethnic groups. While most felt it was important that their child was immunised on time, Bell et al reported that many parents felt the risk of catching a vaccine-preventable infectious disease was reduced due to limited mixing with others. This suggests that an enhanced appreciation of the utility of immunisation was not a major motivator for parents to ensure they attended the immunisation appointments. These issues may well be reflected in the fall in immunisation uptake in England during the earlier part of lockdown reported here and a delay in receiving the first dose 6-in-1 may well led to delays in subsequent doses meaning the infants were unable to ‘catch-up’ by six months of age.

Other factors which may have had an impact on promoting timely immunisation uptake could include a reduction in fever, cough and colds which may otherwise have caused parents to delay immunisations. Though specific data on this point are lacking, a significant fall in the detection of Rhinovirus in adults was observed in 2020 compared to 2019 (32) suggests that ‘normal’ childhood respiratory infection are likely to have decreased. More work is required to fully dissect the key factors in improving timely preschool immunisation uptake in Scotland. Clarity and publicity about the continuation of the immunisation programmes, telephone reminders and the opportunity to discuss with healthcare professions seem likely to have had the most impact.

## Conclusions

The ongoing COVID-19 pandemic continues to stretch health services and adversely affect all areas of life, with children disproportionately bearing the burden of the indirect consequences of pandemic control measures such as the closure of schools and limited social contact. However, opportunities have also been created in terms of enhanced surveillance of health programmes. In this study, we have used such data to investigate the effect of the pandemic on infant and preschool immunisation uptake. We have demonstrated that a robust child immunisation service can continue to deliver high and even increasing uptake rates. Families will respond despite the many difficulties they face, to ensure that children continue to be protected again vaccine-preventable diseases. The challenge now is to use and expand on this knowledge to promote future vaccination programmes, including those targeting SARS-CoV-2.

## Supporting information

prespecified analysis plan

RECORD

## Data Availability

All data used in this study are publicly available on the Public Health Scotland Wider impacts of COVID-19 dashboard which can be accessed via https://scotland.shinyapps.io/phs-covid-wider-impact/. The code used to analyse the data is publicly available via the EAVE II GitHub (https://github.com/EAVE-II).

## Acknowledgments

We thank Public Health Scotland for making the data publicly available and acknowledge the support of the HDR UK BREATHE Hub and EAVE II collaborators.

## Supporting information captions

**S1 Fig:** Percentage uptake by HSCP for 2019 (pale orange) and lockdown (dark orange) with HSCP ordered by uptake for 2019 (note: this varies by immunisation). Dashed horizontal lines indicated the mean uptake for all of Scotland for the time period of the corresponding colour. A = 6in1 immunisation, B = MMR.

**S2 Fig:** Choropleth maps showing all (significant or not) percentage point changes between 2019 and lockdown for each HSCP for all immunisations.

**S3 Fig:** Absolute percentage change in uptake compared to 2019 for each immunisations and SIMD for each time period (A = pre lockdown, B= lockdown, C= post lockdown). Significance rates varied by immunisation and SIMD, for details see table S3.

**S4 Fig:** Combined odds ratio plot with 95% confidence intervals comparing each SIMD quintile (2–5) to SIMD 1-most deprived for 2019 (dark blue) and LD (light blue).

**S5 Fig:** Percentage of children in England immunised by 6 months of age (first, second and third dose 6in1) or 18 months of age (first dose MMR) from January 2019 to September 2020. The start and end of the lockdown period is indicated by the dashed blue lines.

**S1 Table:** Percentage uptake of each immunisation by year (2019), month (Jan and Feb 2020) or week as per data availability. W/B = week beginning.

**S2 Tables A-E:** Percentage uptake, percent point change in uptake compared to 2019 and significance level for this change for each HSCP at each time-period. Each table shows results for a different immunisation. Results were considered significant if *p*-value <0.05 and 95% CI did not include 1. Statistically significant p values are shaded green and significant results for the 2019-LD comparisons are plotted on Figure 2. HSCP = Health and Social Care Partnership, OR = odds ratio, CI = confidence interval, NA = not applicable, PreLD = pre lockdown, LD = lockdown, PostLD = post lockdown. *p*-value rounded to 2 decimal places.

**S3 Table:** Uptake of pre-school immunisations by time period and SIMD and percent point change in uptake compared to baseline 2019. Odds ratio and 95% confidence intervals shown are for change in uptake compared to 2019. *p*-value rounded to 2 decimal places. LD = lockdown, NA = not applicable. Statistically significant change in uptake compared to 2019 are shaded green.

**S4 Table:** To assess whether the differences between change in uptake were statistically significant between SIMD quintiles, the interaction between time period and SIMD quintile was added into the model. The baseline comparisons showed are for time period 2019 and deprivation quintile SIMD 1. ROR = ratio of odds ratio, calculated by taking the exponential function of the coefficient of the interaction term from the interaction model. If the 95% confidence intervals did not include 1, the interaction of time period and SIMD was considered statistically significant, that is; there was a significant difference in the level of change (2019-time period) between the deprivation quintile and SIMD 1. For example, the increase in uptake during lockdown for SIMD 5 was statistically greater than the increase in uptake for SIMD 1. The ROR can be used to calculate the odds ratio for uptake compared to the baseline levels by multiplying the ROR with the relevant OR in table S3. LD= lockdown, SIMD = Scottish Index of Multiple Deprivation, ns = not statistically significant (coloured green) = interaction was statistically significant. *p*-value rounded to 2 decimal places.

**S5 Table:** Scotland. Uptake of pre-school immunisations at an older age by time period and point percentage change from 2019 with odds ratio and 95% confidence intervals compared to baseline of 2019. Children are categorised into the time-period at which they became eligible for the immunisation as before and uptake data were extracted at a later stage when they reached the ages indicated in the immunisation column. LD = lockdown, NA = not applicable. Statistically significant changes are coloured green.

**S6 Table:** England. Uptake of pre-school immunisations at an older age by time period and point percentage change from 2019 with odds ratio and 95% confidence intervals compared to baseline of 2019. Children are categorised into the time-period at which they became eligible for the immunisation as before and uptake data were extracted at a later stage when they reached the ages indicated in the immunisation column. LD = lockdown, NA = not applicable. Statistically significant changes are coloured green.

**S1 File:** Childhood immunisation V1.0 Final analysis plan S2 File: Supplemental methods

**S3 File:** RECORD plus STROBE checklist

## References

1. Prime minister’s statement on coronavirus (COVID 19): 23 March 2020 [press release]. 2020. Availale from https://www.gov.uk/government/speeches/pm-address-to-the-nation-on-coronavirus-23-march-2020, last accessed 25th May 2021

2. Scottish Government. Re-mobilise, Reciver, Re-design: th framework for NHS Scotland. 2020. Available from https://www.gov.scot/publications/re-mobilise-recover-re-design-framework-nhsscotland/, last accessed 25th May 2021

3. Mulholland RH, Wood R, Stagg HR, et al. Impact of COVID-19 on accident and emergency attendances and emergency and planned hospital admissions in Scotland: an interrupted time-series analysis. J R Soc Med. 2020;113(11):444–53.

4. Joint Committee on Vaccination and Immunisation. JCVI statement on immunisation prioritisation 2020. Available from: https://www.gov.uk/government/publications/jcvi-statement-on-immunisation-prioritisation/statement-from-jcvi-on-immunisation-prioritisation, last accessed 25th May 2021

5. Götzinger F, Santiago-García B, Noguera-Julián A, et al. COVID-19 in children and adolescents in Europe: a multinational, multicentre cohort study. Lancet Child Adolesc Health. 2020;4(9):653–61.

6. Viner RM, Mytton OT, Bonell C, et al. Susceptibility to SARS-CoV-2 Infection Among Children and Adolescents Compared With Adults: A Systematic Review and Meta-analysis. JAMA Pediatr. 2020.

7. Araújo LA, Veloso CF, Souza MC, Azevedo JMC, Tarro G. The potential impact of the COVID-19 pandemic on child growth and development: a systematic review. J Pediatr (Rio J). 2020.

8. RCPCH workforce team. The impact of COVID - 19 on child health services - report. 7th May 2020. Avilable from https://www.rcpch.ac.uk/sites/default/files/managed-pdf/Impact%20-of-COVID-19-child-health-services-web.pdf.pdf, last accessed 25th May 2021.

9. Saxena S, Skirrow H, Bedford H. Routine vaccination during covid-19 pandemic response. BMJ. 2020;369:m2392.

10. World Health Organization, WHO and UNICEF warn of a decline in vaccinations during COVID-19 [press release 15 July 2020]. Geneva/New York. Available from https://www.who.int/news/item/15-07-2020-who-and-unicef-warn-of-a-decline-in-vaccinations-during-covid-19, last accessed 25th May 2021.

11. McDonald HI, Tessier E, White JM, et al. Early impact of the coronavirus disease (COVID-19) pandemic and physical distancing measures on routine childhood vaccinations in England, January to April 2020. Euro Surveill. 2020;25(19).

12. Chandir S, Siddiqi DA, Mehmood M, et al. Impact of COVID-19 pandemic response on uptake of routine immunizations in Sindh, Pakistan: An analysis of provincial electronic immunization registry data. Vaccine. 2020;38(45):7146–55.

13. Jensen C, McKerrow NH. Child health services during a COVID-19 outbreak in KwaZulu-Natal Province, South Africa. S Afr Med J. 2020;0(0):13185.

14. Zhong Y, Clapham HE, Aishworiya R, et al. Childhood vaccinations: Hidden impact of COVID-19 on children in Singapore. Vaccine. 2021;39(5):780–5.

15. Santoli JM, Lindley MC, DeSilva MB, et al. Effects of the COVID-19 Pandemic on Routine Pediatric Vaccine Ordering and Administration - United States, 2020. MMWR Morb Mortal Wkly Rep. 2020;69(19):591-3.

16. Department of Health. Immunisation against infectious disease: The Green book. Public Health England; 2021.

17. PM annouces easing of lockdown restrictions: 23 June 2020 [press release]. Available from https://www.gov.uk/government/news/pm-announces-easing-of-lockdown-restr-ictions-23-june-2020, last accessed 25th May 2021

18. Scottish Government. Scotland’s routemap through and out of the crisis. 2020. Available from https://www.gov.scot/publications/coronavirus-covid-19-framework-decision-making-scotlands-route-map-through-out-crisis/pages/2/2020. last accessed 25th May 2020

19. Scottish Government. Introducing the Scottish Index of Multiple Deprivation 2020. National statistics publication; 2020.

20. von Elm E, Altman DG, Egger M, et al. The Strengthening the Reporting of Observational Studies in Epidemiology (STROBE) statement: guidelines for reporting observational studies. J Clin Epidemiol. 2008;61(4):344–9.

21. Benchimol EI, Smeeth L, Guttmann A, et al. The REporting of studies Conducted using Observational Routinely-collected health Data (RECORD) statement. PLoS Med. 2015;12(10):e1001885.

22. Haider EA, Willocks LJ, Anderson N. Identifying inequalities in childhood immunisation uptake and timeliness in southeast Scotland, 2008-2018: A retrospective cohort study. Vaccine. 2019;37(37):5614-24.

23. Forster AS, Rockliffe L, Chorley AJ, et al. Ethnicity-specific factors influencing childhood immunisation decisions among Black and Asian Minority Ethnic groups in the UK: a systematic review of qualitative research. J Epidemiol Community Health. 2017;71(6):544–9.

24. National Office of Statistics, National Records of Scotland 2011 census aggregate data Scotland NRo, Northern, 2016. Available from http://dx.doi.org/10.5257/census/aggregate-2011-1, last accessed 25th May 2021

25. Public Health Scotland Data and Intelligence. Childhood Immunisation Statistics Scotland - Quarter ending 30 September 2019. 2019. Available from https://www.isdscotland.org/Health-Topics/Child-Health/publications/data-tables2017.asp?id=2574#2574, last accessed 25th May 2021

26. Royal Society for Public Health. Moving the needle: Promoting vaccination uptake across the life course. 2019. Available from https://www.rsph.org.uk/static/uploaded/3b82db00-a7ef-494c-85451e78ce18a779.pdf, last accessed 25th May 2021

27. HM revenus and Customs. Coronavirus Job Retention Scheme Official Statistics. 2020. Available from https://assets.publishing.service.gov.uk/government/uploads/system/uploads/attachment_data/file/891249/Coronavirus_Job_Retention_Scheme_Statistics_June_2020.pdf, last accessed 25th May 2021

28. Jarchow-MacDonald AA, Burns R, Miller J, Kerr L, Willocks LJ. Keeping childhood immunisation rates stable during the COVID-19 pandemic. Lancet Infect Dis. 2021.

29. Campbell H, Edwards A, Letley L, Bedford H, Ramsay M, Yarwood J. Changing attitudes to childhood immunisation in English parents. Vaccine. 2017;35(22):2979–85.

30. Jacobson Vann JC, Szilagyi P. Patient reminder and patient recall systems to improve immunization rates. Cochrane Database Syst Rev. 2005(3):CD003941.

31. Bell S, Clarke R, Paterson P, Mounier-Jack S. Parents’ and guardians’ views and experiences of accessing routine childhood vaccinations during the coronavirus (COVID-19) pandemic: A mixed methods study in England. PLoS One. 2020;15(12):e0244049.

32. Poole S, Brendish NJ, Tanner AR, Clark TW. Physical distancing in schools for SARS-CoV-2 and the resurgence of rhinovirus. Lancet Respir Med. 2020;8(12):e92–e3.

