## Supplementary material for "Uptake of infant and pre-school immunisations in Scotland and England during the COVID-19 pandemic: an observational study of routinely collected data": prespecified analysis plan

Impact of COVID-19 and associated control measures on uptake of pre-school immunisations

Analysis Plan

| **Full Project Title** | Impact of COVID-19 and associated control measures on uptake of pre-school immunisations |
| --- | --- |
| **Version Number** | 1.0 |
| **Previous Versions** | 0.2 |
| **Effective Date** | 27/01/2021 |
| **EAVE II Sub-study** |  |
| **EAVE II Analyst(s)** | Rachel Mulholland (UofE) |
| **Co-authors** | Scotland:  Fiona McQuaid* (NHS Lothian, UofE)  Claire Cameron (PHS)  Cheryl Gibbon (PHS)  Aziz Sheikh (UofE)  Judith Tait (PHS)  Steve Turner (RCPCH, University of Aberdeen)  Jaime Villacampa Ortega (PHS)  Rachael Wood (PHS, UofE)  England:  Helen Bedford (ICH, London)  Elise Tessier (PHE)  Yuma Sanpang Rai (PHE)  Joanne White (PHE)  Partho Roy (PHE) |
| **Target journal** | BMJ |

*Corresponding author

### 1 Introduction

The COVID-19 pandemic and resulting national lockdown has had a profound impact on the delivery of healthcare with the reconfiguration of acute services to support the anticipated influx of COVID-19 patients, cancellation of most elective activity and pausing of screening programmes (1). There is also evidence of a change in healthcare-seeking behaviour, for example, in Scotland the uptake of both emergency and elective hospital based care dropped substantially over the lockdown period (2). However, within child health, key routine services such as childhood immunisations and health visiting continued across Scotland and a campaign launched in April 2020 urged the public to remember “The NHS is open” (https://www.gov.scot/news/urgent-medical-help-still-available).

It has become increasingly apparent that younger children are at low risk of severe disease due to SARS-CoV-2 (3, 4) and may be less susceptible to infection by the virus (5). Yet the wider impact of the pandemic on children in terms of education, mental and physical health and safeguarding is not yet fully understood, though it is likely to be profound (6, 7). One particular area of concern early in the lockdown period was the potential effect on the uptake of routine childhood immunisations (8). Maintaining high population vaccine coverage is vital for both direct and indirect (via herd immunity) protection against non-COVID-19 infectious diseases. The current pre-school UK vaccination schedule is shown in table 1 along with the disease protected against.

Table 1 UK immunisation schedule 2020 (9). NA= not referenced/used in this manuscript.

| Age due | Diseases protected against | Vaccine given | Referred in this manuscript as: |
| --- | --- | --- | --- |
| 8 weeks | Diphtheria, tetanus, pertussis (whooping cough), polio, Haemophilus influenzae type b (Hib) and hepatitis B | DTaP/IPV/Hib/HepB | First dose 6in1 |
|  | Meningococcal group B (MenB) | MenB | NA |
|  | Rotavirus gastroenteritis | Rotavirus | NA |
| 12 weeks | Diphtheria, tetanus, pertussis, polio, Hib and hepatitis B | DTaP/IPV/Hib/HepB | Second dose 6in1 |
|  | Pneumococcal (13 serotypes) | Pneumococcal conjugate vaccine (PCV) | NA |
|  | Rotavirus gastroenteritis | Rotavirus | NA |
| 16 weeks | Diphtheria, tetanus, pertussis, polio, Hib and hepatitis B | DTaP/IPV/Hib/HepB | Third dose 6in1 |
|  | Meningococcal group B (MenB) | MenB | NA |
| 1 year (after first birthday) | Hib and Meningococcal group C (MenC) | Hib/MenC | NA |
|  | Measles, mumps and rubella | MMR | First dose MMR |
|  | Pneumococcal | PCV booster | NA |
|  | MenB | MenB booster | NA |
| 3 years 4 months (or shortly after) | Diphtheria, tetanus, pertussis and polio | dTaP/IPV | NA |
|  | Measles, mumps and rubella | MMR booster | Second dose MMR |

Initial reports suggested a fall in children receiving their scheduled vaccinations in the very early lockdown period (8). The aim of this paper is to use routinely collected and publicly available child health surveillance data in Scotland to describe the pattern of pre-school vaccine uptake during the pandemic period, with comparisons to 2019 data, by geographical area and socio-economic index. Of note, this paper refers to the first national lockdown which began on 23^rd^ March 2020 with restrictions easing gradually from June 2020. The emergence in the UK of a new, highly transmissible, SARS-CoV-2 variant in late 2020 (10) has since prompted further control measures (essentially further lockdowns in Scotland and England with similar measure in Wales and Northern Ireland). Data continue to be collected on the impact on vaccine uptake and should be further evaluated when available, however this paper deals solely with the first national lockdown period.

We have contacted colleagues in child health surveillance from the other nations of the UK (England, Wales and Northern Ireland) to request access to equivalent data and should this become available, we will aim to describe the patterns of pre-school vaccine uptake as above for each of the nations which form the UK.

### 2 Aims and objectives

#### 2.1 Aims

We aim to describe the impact of COVID-19 and associated control measures on the uptake of pre-school immunisations in Scotland, with potential to expand to other UK nations should additional data become available.

#### 2.2 Objectives

We seek to:

a. Describe the impact of the COVID-19 pandemic on the uptake of selected immunisations provided at each of the five immunisation contacts offered to pre-school children (table 1)
b. Explore whether impact varied by: i) Health and social care partnership area of residence (HSCP- geographical areas closely related to local authority areas); and ii) socio-economic deprivation index
c. Use these data to inform future strategies to promote maximal childhood vaccine uptake and reduce barriers, both in the context of a pandemic and more generally.

### 3 Study Design

#### 3.1 Study design

Natural experiment, designed to take advantage of routinely collected data in the year prior to the COVID-19 pandemic and immediately before, during the first period of ‘lockdown’ (23 March-July 2020) imposed by the Scottish and UK and other devolved governments, and after the lockdown restrictions began to be eased (August- September 2020).

#### 3.2 Setting

Scotland, UK with potential scope to extend to England, Wales and Northern Ireland.

#### 3.3 Population

All children in Scotland (and if available; England, Wales and Northern Ireland) who become eligible (based on age), for routine immunisations as shown in Table 1 (specifically; first, second and third dose 6 in1, and first and second dose MMR) from January 2019 to September 2020.

#### 3.4 Data sources

The Public Health Scotland (PHS) [COVID-19 wider impacts dashboard](https://scotland.shinyapps.io/phs-covid-wider-impact/_w_c89e0e10/) (<https://scotland.shinyapps.io/phs-covid-wider-impact/>), which presents data drawn from the Scottish Immunisation & Recall System (SIRS). SIRS is the dataset which records all information on children eligible for and receiving routine preschool immunisation in Scotland. These data are publicly available via the link above.

Equivalent data has been sought from England, Wales and Northern Ireland.

In England, data are extracted from ImmForm, a Public Health England website used to collect vaccine coverage data and provide vaccine ordering facilities. At present, due to data sharing agreements, it is likely that data from England will be analysed “in-house” by scientists at Public Health England, and results shared with the wider team. This analysis plan and the code used to analysis the Scottish data will be shared and similar methods applied within the constraints of which data are available (likely uptake of 6in1 vaccine at 6 months of age, and MMR at 12 and 18 months).

In Northern Ireland, data are collected by the Northern Ireland Child health System, the format and accessibility of which is being explored.

In Wales, data are provided by NHS Wales Informatics Service from the National Community Child Health Database (NCCHD). This is sourced from Community Child Health databases maintained by local Child Health Office staff in Trusts throughout Wales based on regular returns from nurses and doctors who immunise or advise on immunisation.

#### 3.5 Ethical approval

Ethical approval for this specific study was not required as we are using publicly available, anonymised, aggregated data.

#### 3.6 Inclusion/exclusion criteria

All children living in Scotland (or other UK nation) who became eligible based on age for the relevant vaccine are included, with the exception of those registered to receive their second dose MMR immunisation in HSCPs associated with NHS Grampian. This is because this immunisation is offered to children when they turn 4 years of age in Grampian, rather than when they turn 3 years and 4 months of age as in all other Board areas.

Note 1: During this period, some children would become eligible for more than one set of immunisations (for example a child aged 8 weeks in January 2020, would become eligible for first, second and third doses of 6in1 vaccine during the study period). These children are included for each separate dose.

Note 2: A very small number of children would be medically exempt from the MMR vaccine as it is a live vaccine, however these children are still included as eligible for the purposes of this analysis.

Note 3: It is possible for parents to consent to receive selected vaccines only (for example to decline the MMR, but still receive the other vaccines offered at 1 year of age), therefore caution must be taken if extrapolating these data to represent uptake of the other pre-school immunisations for which data are not currently publicly available.

#### 3.7 Sample size calculation

Taking in entire eligible population

### 4 Data and data validation

#### 4.1 Data variables available

Table 2 Available variables (Scottish dataset)

| **Variable** | **Description** | **Values** |
| --- | --- | --- |
| Vaccine type | Type of preschool immunisation vaccine | First dose 6in1 (8 weeks)  Second dose 6in1 (12 weeks)  Third dose 6in1 (16 weeks)  First dose MMR (12 months)  Second dose MMR (3 years and 4 months) |
| Area name | Splits into Scotland as a whole, NHS Health Boards and HSCP | Scotland  NHS Health Boards  HSCP |
| Time-period | Time-period when preschool child was eligible for immunisation | 2019 (baseline),  January to September 2020 (Monthly)  Week beginning 02 March to 28 September 2020 (Weekly) |
| Total eligible | The total number of eligible preschool children | N |
| Total uptake within 4 weeks of eligibility (N)* | The number of preschool children who received vaccination within 4 weeks of becoming eligible | N |
| Total uptake within 4 weeks of eligibility (%) | The number of preschool children who received vaccination within 4 weeks of becoming eligible out of the total number of eligible preschool children | % [0,100] |
| SIMD Quintile (for Scotland level data only) | Scottish Index of Multiple Deprivation (SMID) index | 1 (most deprived), 2, 3, 4, 5 (least deprived) |

*Note: Data are available for total uptake at later ages (for example 6 months of age). However, we have chosen to examine uptake within 4 weeks of eligibility as this represents timely uptake of vaccinations as per the recommended schedule.

#### 4.2 Constructed variables

Table 3 Constructed variables

| **Variable** | **Description** | **Values** |
| --- | --- | --- |
| Absolute change from 2019 (%) | % uptake within 4 weeks of eligibility in 2020 – % uptake within 4 weeks of eligibility in 2019 | [-100,100] % |
| Relative change from 2019 (%) | Absolute change from 2019 (%) / % uptake within 4 weeks of eligibility in 2019 | [-1,Inf] % |

#### 4.3 Consistency and error checking

Data quality has already been checked by PHS.

### 5 Statistical analyses

The following sections are presented separately for each analytical objective. All analyses will be conducted for data from Scotland. Depending on the data available for the other UK nations, similar analyses will be conducted in parallel. At present, the variations in data (in terms of collection methods and availability and varying vaccination policies) suggest that pooling the UK data or conducting a meta-analysis would not be appropriate, however any differing trends may be discussed in a descriptive manner.

#### 5.1 Objective a) Describe the impact of the COVID-19 pandemic on the uptake of immunisations provided at each of the five immunisation contacts offered to pre-school children

##### 5.1.1 Outcome

The primary outcome is the % uptake of preschool immunisations (represented by uptake of one of the vaccines due at that age) within 4 weeks of eligibility. As a secondary outcome, the % uptake of preschool immunisations by 6 months of age (first dose 6in1) or 16 -18 months of age (first dose MMR) may also be considered to allow for comparisons with data from other nations (TBC).

##### 5.1.2 Exposures of interest

Time, specifically in time-periods: 2019 (baseline), 01 Jan- 22 Mar 2020 (pre-lockdown), 23 Mar-final week Jul 2020 (lockdown) and Aug-end of Sept 2020 (post lockdown). The start date to 23 Mar has been chosen to correspond with the beginning of the UK wide lockdown as announced by the UK government. The end of the lockdown period is less well-defined and varied both in approach and timescale between Scotland, England, Wales and Northern Ireland. Broadly speaking, by the end of July, there was a substantial reduction in ‘lockdown’ restrictions with the opening of many non-essential businesses and limited indoor meeting between households permitted, therefore a pragmatic approach has been taken to define the lockdown period as 23 Mar 2020 until end Jul 2020. The data included in each time period are shown in Table 4.

Table 4 Definition of time periods

| Time period | Data included |
| --- | --- |
| Baseline 2019 | Aggregate data for whole of 2019 |
| Pre-Lockdown 2020 | Monthly data for Jan 2020 and Feb 2020. Weekly data from W/B 2Mar2020 up to and including W/B 16Mar2020 |
| Lockdown 2020 | Weekly data from W/B 23Mar2020 up to and including W/B 27Jul2020 |
| Post lockdown 2020 | Weekly data from W/B 3Aug2020 up to and including W/B 28Sept2020 |

The primary outcome of interest is comparison between baseline 2019 and lockdown 2020 for each vaccine. Of secondary interest are comparisons between baseline 2019 and pre-lockdown 2020, baseline 2019 and post lockdown 2020, pre-lockdown 2020 and lockdown 2020, lockdown 2020 and post lockdown 2020.

##### 5.1.3 Analytical techniques

To compare the % uptake of preschool immunisation across time-periods we will visualise the total % uptake within 4 weeks of eligibility for each time-period by vaccine in a bar plot.

To statistically test whether these uptake rates are different, we will perform a binary logistical regression analysis for aggregate data, using time period as the explanatory variable, and vaccination status (vaccinated or unvaccinated) as the dependent variable. Separate analyses will be carried out for each vaccine. Odds ratios with 95% confidence intervals will be calculates using 2019 as the baseline comparator.

The following tables will be utilised:

Table 5 Odds of being vaccinated between time-periods for each vaccine visit

| Vaccine | Time-period comparison | Odds ratio | 95% CI |
| --- | --- | --- | --- |
| First dose 6in1 | 2019 vs pre-lockdown (secondary outcome) |  |  |
|  | **2019 vs lockdown (primary outcome)** |  |  |
|  | Pre-lockdown vs lockdown (secondary outcome) |  |  |
|  | 2019 vs post-lockdown (secondary outcome) |  |  |
|  | Lockdown vs post-lockdown (secondary outcome) |  |  |
| Second dose 6in1 | 2019 vs pre-lockdown (secondary outcome) |  |  |
|  | **2019 vs lockdown (primary outcome)** |  |  |
|  | Pre-lockdown vs lockdown (secondary outcome) |  |  |
|  | 2019 vs post-lockdown (secondary outcome) |  |  |
|  | Lockdown vs post-lockdown (secondary outcome) |  |  |
| … |  |  |  |
| Second dose MMR | 2019 vs pre-lockdown (secondary outcome) |  |  |
|  | **2019 vs lockdown (primary outcome)** |  |  |
|  | Pre-lockdown vs lockdown (secondary outcome) |  |  |
|  | 2019 vs post-lockdown (secondary outcome) |  |  |
|  | Lockdown vs post-lockdown (secondary outcome) |  |  |

##### 5.1.4 Potential confounders

None available since aggregated data

##### 5.1.5 Potential effect modifiers

None at this stage

##### 5.1.6 Sub-group analysis

None

##### 5.1.7 Corrections for multiple testing

None

##### 5.1.8 Sensitivity analysis

None

##### 5.1.9 Other analysis

None

#### 5.2 Objective bi) Explore the difference in impact between geographical areas

##### 5.2.1 Outcome

Absolute/relative difference in % uptake within 4 weeks of eligibility during lockdown vs 2019 for each Health and Social Care Partnership (HSCP).

##### 5.2.2 Exposures of interest

Geographical area, specifically HSCP (excluding partnership areas within NHS Grampian for the MMR second dose only).

##### 5.2.3 Analytical techniques

To visualise the spatial distribution of the difference in lockdown and 2019, the % differences will be plotted in a choropleth map of Scotland segmented into HSCPs. This map will have each HSCP area coloured according to the % difference. This will be repeated for each vaccine type.

To statistically test whether these uptake rates are different between time periods for each HSCP, we will perform a binary logistical regression analysis for aggregate data, using vaccination status (vaccinated or unvaccinated) as the dependent variable and time period as the explanatory variable, specifying an interaction with HSCP. Separate analyses will be carried out for each vaccine. Odds ratios with 95% confidence intervals will be calculated using 2019 as the baseline comparator and results will be visualised using forest plots. The following table will be used (one for each vaccine):

Table 6 Odds of being vaccinated between time-periods by HSCP

| Time-period | Variable | Difference % | Odds ratio | 95% CI |
| --- | --- | --- | --- | --- |
| 2019 vs lockdown | Aberdeen City |  |  |  |
|  | Aberdeenshire |  |  |  |
|  | Angus |  |  |  |
|  | … |  |  |  |

##### 5.2.4 Potential confounders

None

##### 5.2.5 Potential effect modifiers

None

##### 5.2.6 Sub-group analysis

None

##### 5.2.7 Corrections for multiple testing

None

##### 5.2.8 Sensitivity analysis

None

##### 5.2.9 Other analysis

None

#### 5.3 Objective bii) Explore the difference in impact between socio-economic deprivation

##### 5.3.1 Outcome

Absolute/relative difference in % uptake within 4 weeks of eligibility during lockdown vs total 2019 by SMID quintile.

##### 5.3.2 Exposures of interest

Scottish Index of Multiple Deprivation (SIMD) quintiles

##### 5.3.3 Analytical techniques

To explore the difference in the % uptake during lockdown (23Mar-Jul 2020) versus 2019 across the deprivation quintiles, we will plot the % difference for each vaccine by the SIMD quintile in a bar plot.

To statistically test whether these uptake rates are different between time periods for each SMID quintile, we will perform a binary logistical regression analysis for aggregate data, using time period as the explanatory variable, and vaccination status (vaccinated or unvaccinated) as the dependent variable, specifying an interaction with SIMD quintile. Separate analyses will be carried out for each vaccine. Odds ratios with 95% confidence intervals will be calculated using 2019 as the baseline comparator. The following tables will be utilised (one for each vaccine):

Table 7 Odds of being vaccinated between time-periods by SMID

| Time-period | Variable | Difference % | Odds ratio | 95% CI |
| --- | --- | --- | --- | --- |
| 2019 vs lockdown | SMID 1 |  |  |  |
|  | SMID 2 |  |  |  |
|  | SMID 3 |  |  |  |
|  | SMID 4 |  |  |  |
|  | SMID 5 |  |  |  |

A similar technique will be carried out to determine the odds of being vaccinated by SMID quintile at each time point (2019 and lockdown), using SMID 1 (most deprived) as the baseline comparator, to assess for change in the inequality between SMID quintiles between the two time periods.

##### 5.3.4 Potential confounders

None

##### 5.3.5 Potential effect modifiers

None

##### 5.3.6 Sub-group analysis

None

##### 5.3.7 Corrections for multiple testing

None

##### 5.3.8 Sensitivity analysis

None

##### 5.3.9 Other analysis

None

#### 5.4 Missing data

As noted in section 3.6, for analyses relating to the second dose of MMR only, data relating to children registered to receive their immunisations in HSCP within NHS Grampian will be excluded. This is because this immunisation is offered to children when they turn 4 years of age in Grampian, rather than when they turn 3 years and 4 months of age as in all other Board areas.

A small number of children have missing information on HSCP area of residence and/or SIMD quintile. Numbers with missing data will be provided as supporting information for the analyses by HSCP and SIMD. No information is provided through the PHS dashboard on immunisation uptake for children with missing HSCP/SIMD data so this will not be reported.

#### 5.5 Statistical software

All analyses will be performed on R/R Studio (4.0.3)

### 6 Reporting results

#### 6.1 Reporting guidelines and conventions

Results will be reported according to STROBE (11) and RECORD (12) (via the COVID-19 extension) guidelines. P-values will be quoted to two decimal places, unless they are less than 0.001 (whereby the p-value will be given as <0.001) or between <0.005 and >0.001, in which case they will be stated to three decimal places. We will report 95% confidence intervals and if the confidence intervals cross 1, the results will not be considered statistically significant.

#### 6.2 Dissemination

The analysis will be written in a manuscript and submitted to a peer reviewed journal. The chosen journal we have decided to aim for is the BMJ.

We will distribute finding to leads for immunisation policy and delivery in the Scottish government, Public Health Scotland, and territorial NHS Boards through the Scottish Immunisation Group. Results will also be shared with equivalent bodies of other UK nations. Findings will also be discussed with the Royal College of Paediatrics and Child Health and British Association for Child and Adolescent Public Health and may be presented at local, regional, national and/or international meetings.

Key messages will be made into an infographic to be disseminated on social media and communication channels. All code will be made publicly available via the EAVE II GitHub (<https://github.com/EAVE-II>).

### 7 References

1.	Scottish Government, Re-mobilise, Recoved, Re-design: the feamework for NHS Scotland, 2020.

2.	Mulholland RH, Wood R, Stagg HR, Fischbacher C, Villacampa J, Simpson CR, et al. Impact of COVID-19 on accident and emergency attendances and emergency and planned hospital admissions in Scotland: an interrupted time-series analysis. J R Soc Med. 2020;113(11):444-53.

3.	Swann OV, Holden KA, Turtle L, Pollock L, Fairfield CJ, Drake TM, et al. Clinical characteristics of children and young people admitted to hospital with covid-19 in United Kingdom: prospective multicentre observational cohort study. BMJ. 2020;370:m3249.

4.	Dong Y, Mo X, Hu Y, Qi X, Jiang F, Jiang Z, et al. Epidemiology of COVID-19 Among Children in China. Pediatrics. 2020;145(6).

5.	Viner RM, Mytton OT, Bonell C, Melendez-Torres GJ, Ward J, Hudson L, et al. Susceptibility to SARS-CoV-2 Infection Among Children and Adolescents Compared With Adults: A Systematic Review and Meta-analysis. JAMA Pediatr. 2020.

6.	Araújo LA, Veloso CF, Souza MC, Azevedo JMC, Tarro G. The potential impact of the COVID-19 pandemic on child growth and development: a systematic review. J Pediatr (Rio J). 2020.

7.	Royal College of Paediatrics and Child Health workforce team . The impact of COVID - 19 on child	health services	- report. 2020. Available from <https://www.rcpch.ac.uk/resources/impact-covid-19-child-health-services-report>

8.	Saxena S, Skirrow H, Bedford H. Routine vaccination during covid-19 pandemic response. BMJ. 2020;369:m2392.

9.	Public Health England. Complete routine immunisation schedule from January 2020 2020 [updated June 2020. Available from: <https://assets.publishing.service.gov.uk/government/uploads/system/uploads/attachment_data/file/899423/PHE_Complete_Immunisation_Schedule_Jun2020_05.pdf>.

10.	Public Health England. Investigation of novel SARS - C o V - 2	variant. 2020.

11.	von Elm E, Altman DG, Egger M, Pocock SJ, Gøtzsche PC, Vandenbroucke JP, et al. The Strengthening the Reporting of Observational Studies in Epidemiology (STROBE) statement: guidelines for reporting observational studies. J Clin Epidemiol. 2008;61(4):344-9.

12.	Benchimol EI, Smeeth L, Guttmann A, Harron K, Moher D, Petersen I, et al. The REporting of studies Conducted using Observational Routinely-collected health Data (RECORD) statement. PLoS Med. 2015;12(10):e1001885.
