## Supplementary material for "Uptake of infant and pre-school immunisations in Scotland and England during the COVID-19 pandemic: an observational study of routinely collected data": S2 File supplemental methods

*Additional inclusion/exclusion criteria*

During the study period, some children would become eligible for more than one set of immunisations (for example, a child aged 8 weeks in January 2020, would become eligible for first, second and third doses of 6in1 vaccine during the study period) and these children were included for each separate dose. Of note, a very small number of children would be medically exempt from the MMR vaccine as it is a live vaccine (15), however these children are still included as eligible for the purposes of this analysis. For the analyses of the second dose MMR only, we excluded children registered to receive this vaccine in Health and Social Partnerships associated with NHS Grampian (Aberdeen City, Aberdeenshire and Moray) as this immunisation is offered to children when they turn 4 years of age in Grampian, rather than when they turn 3 years and 4 months of age as in all other areas.

*Additional information on data sources*

*Scotland*

The “COVID19 wider impacts on the health care system” dashboard was set up by Public Health Scotland in response to the COVID-19 pandemic to provide near real-time updates from a range of national databases on topics ranging from child health to cancer, thus allowing for rapid analysis of trends (19). (18). Data for childhood immunisations were drawn monthly from the Scottish Immunisation & Recall System (SIRS), an electronic system used by all NHS boards in Scotland by which information on children eligible for and receiving routine preschool immunisation is recorded by administrative staff within the relevant NHS board (19). The code used by Public Health Scotland to produce the wider impacts dashboard can be accessed at <https://github.com/Public-Health-Scotland/covid-wider-impacts>.

Data were available for all children living in Scotland and eligible for the five immunisations of interest with the above noted exceptions. For 2019, aggregate data for the entire year were used, while for 2020, monthly data were used for January and February 2020, then weekly from 2^nd^ March 2020 (table S1). For the island HSCPs (Shetland, Orkney and the Western Isles), and for England, only monthly data were available therefore lockdown was defined as the months April to July inclusive.

Of note, data checking by Public Health Scotland revealed that a changes to the reporting database to include scheduling for some adult immunisations meant that a small number of older adults born in 1920 were included in the denominator for the January 2020 data (further details can be found in the 23 December release commentary on <https://scotland.shinyapps.io/phs-covid-wider-impact/>). It was not possible to remove these older adult records, therefore, uptake rates for January 2020 may have been slightly under-reported. This fault was rectified by the next dashboard update in February.
