## Supplementary figures and images for "Uptake of infant and pre-school immunisations in Scotland and England during the COVID-19 pandemic: an observational study of routinely collected data"

### FigS1

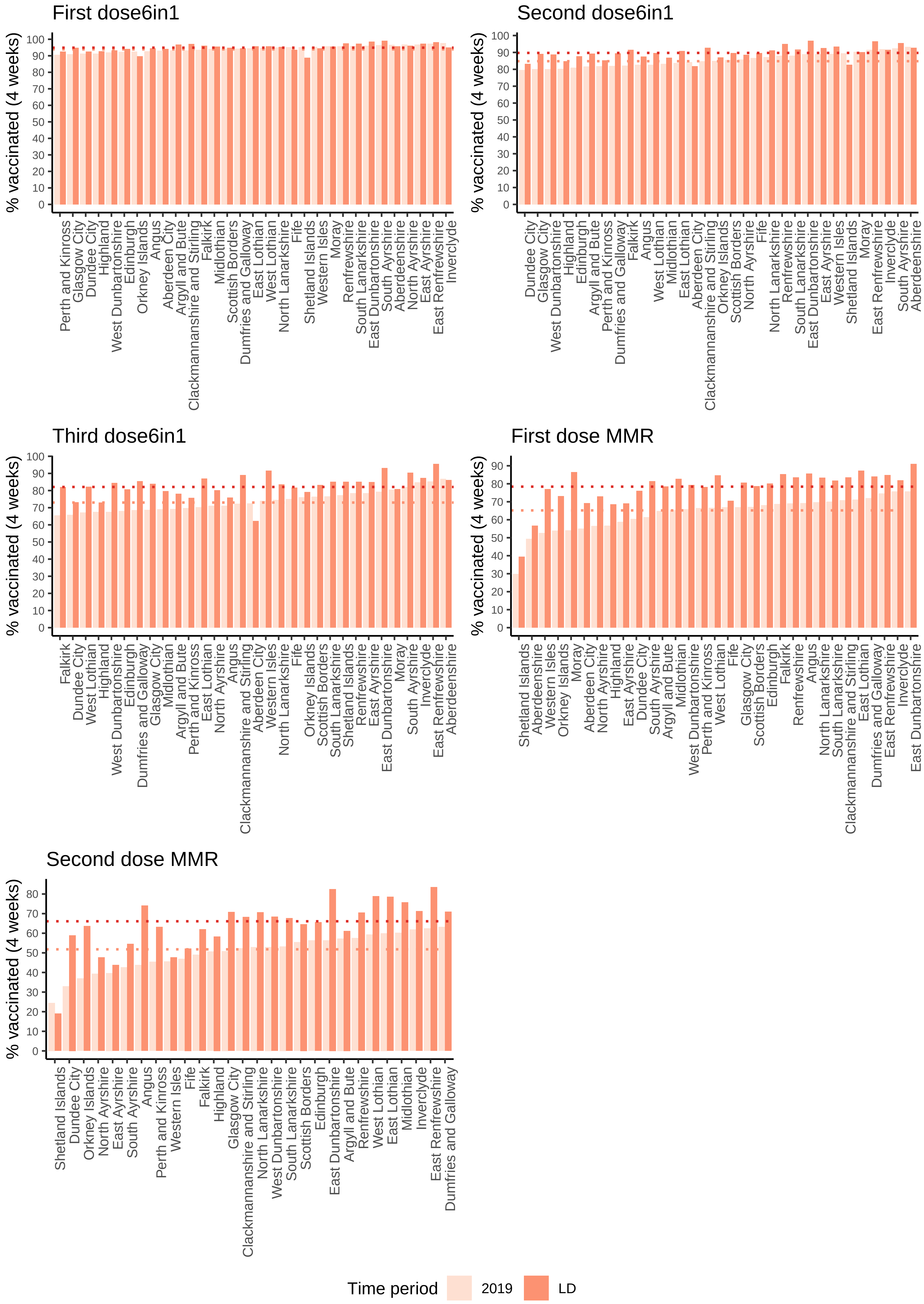

### FigS3

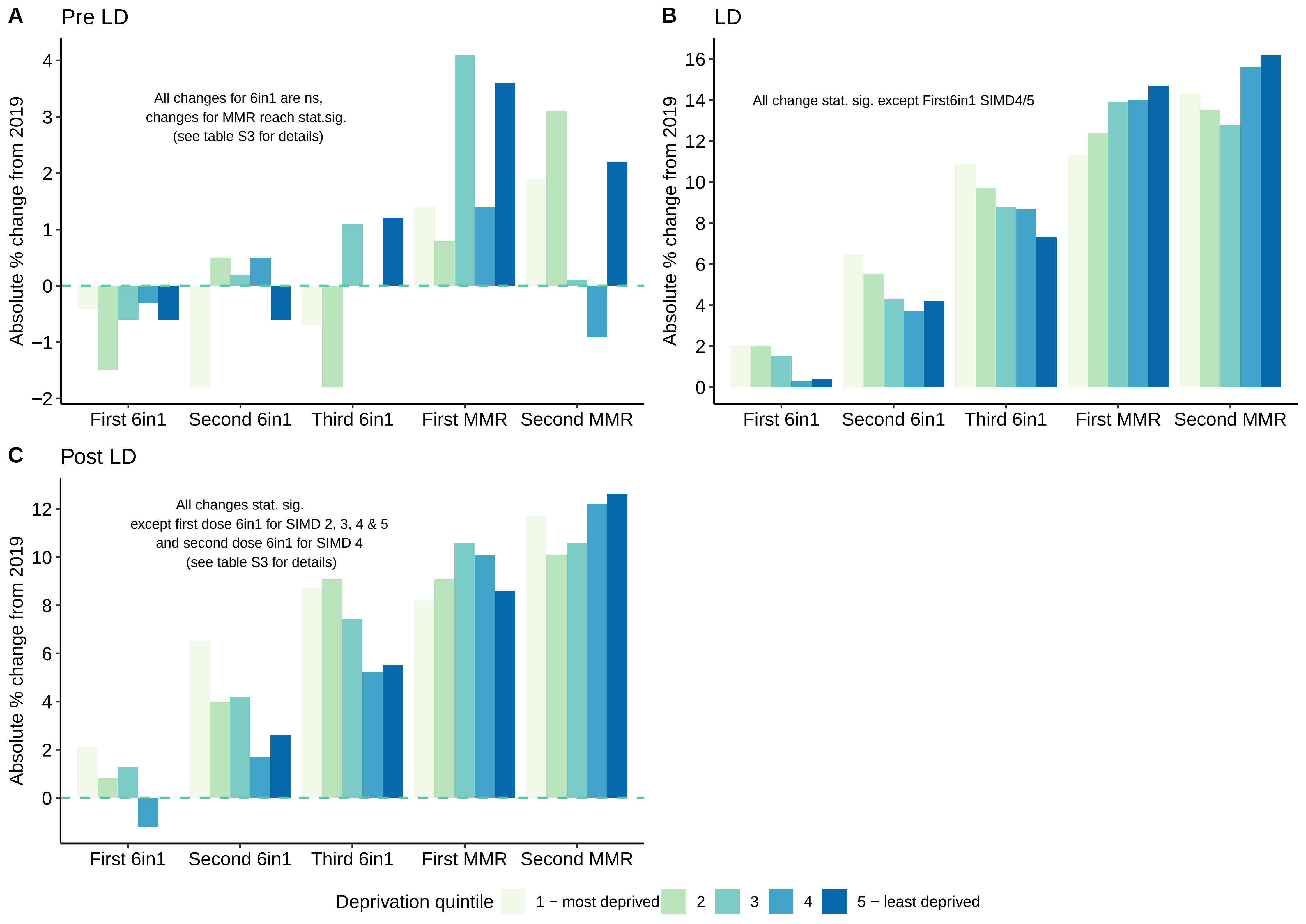

### FigS4

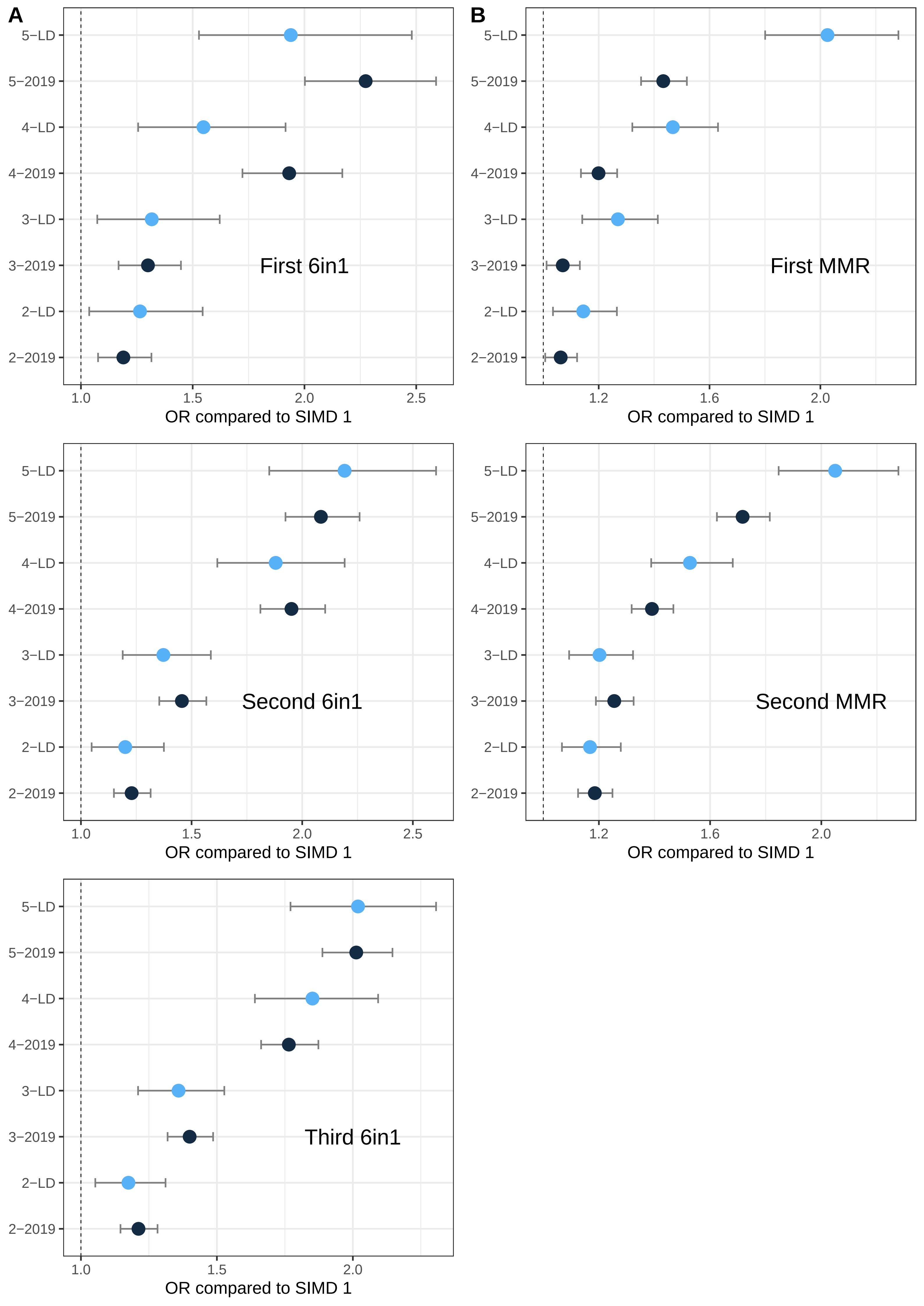

### FigS5

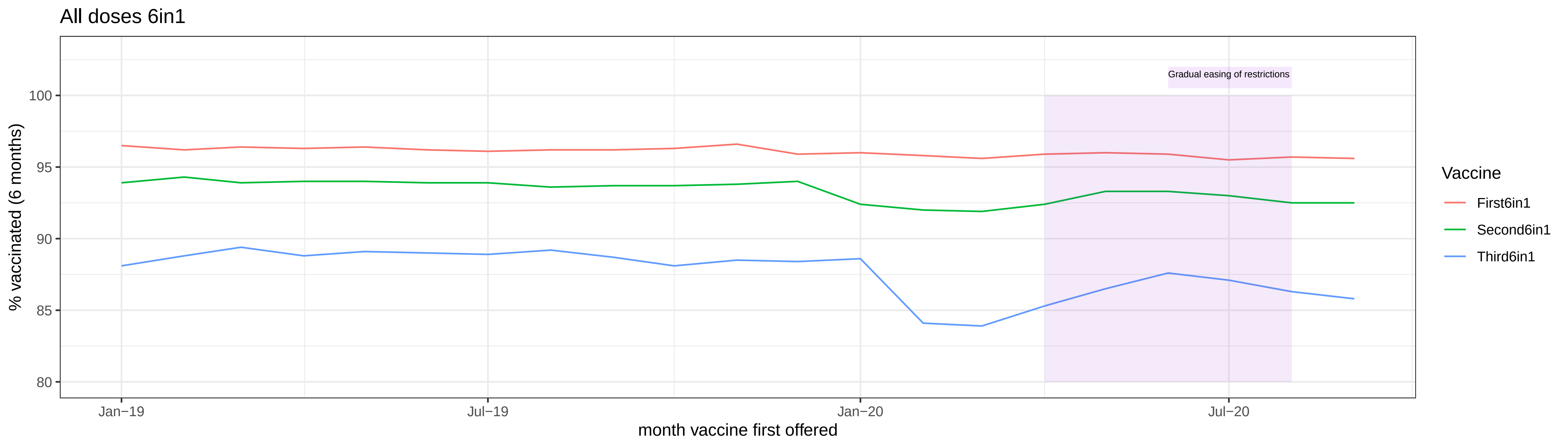

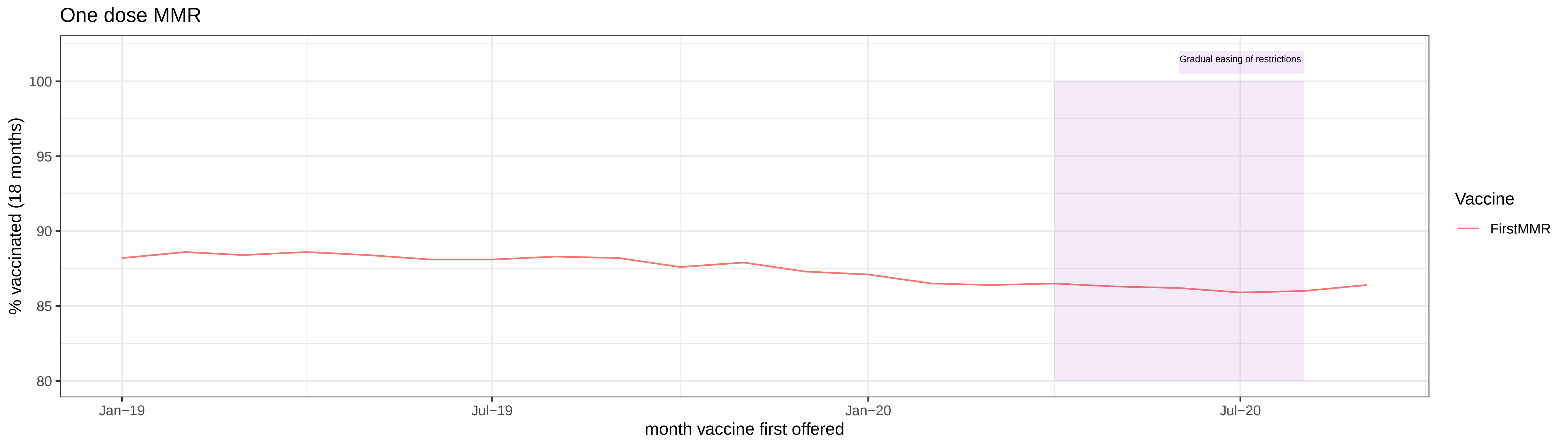
