## Supplementary material for "Uptake of infant and pre-school immunisations in Scotland and England during the COVID-19 pandemic: an observational study of routinely collected data": FigS2

### First 6in1 % change from 2019

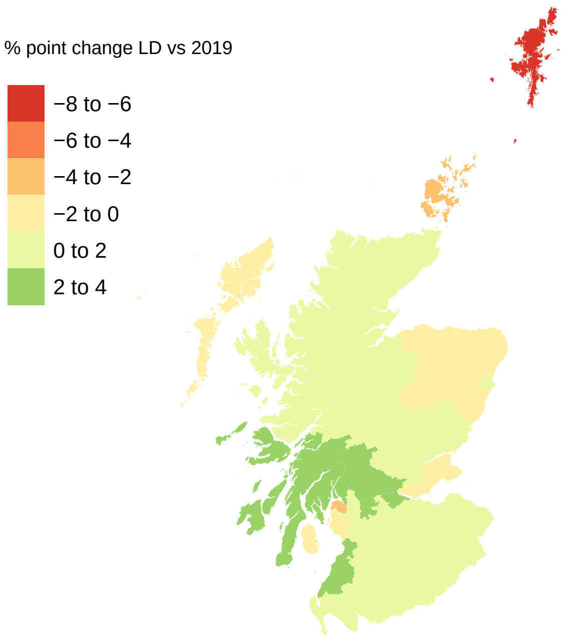


### Second 6in1 % change from 2019

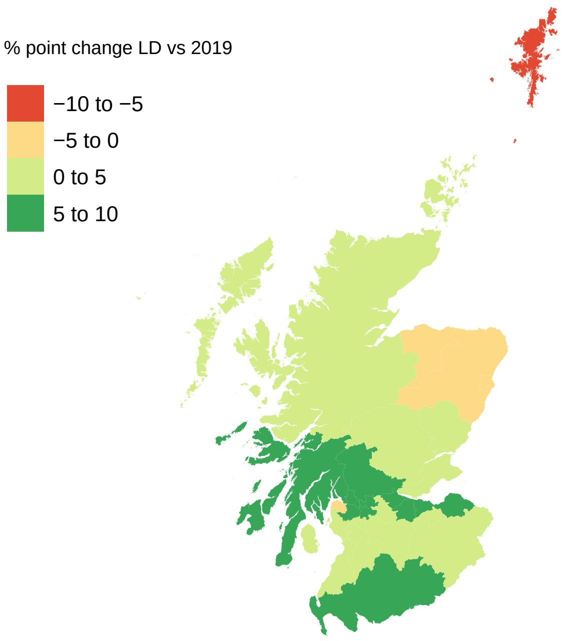


### Third 6in1 % change from 2019

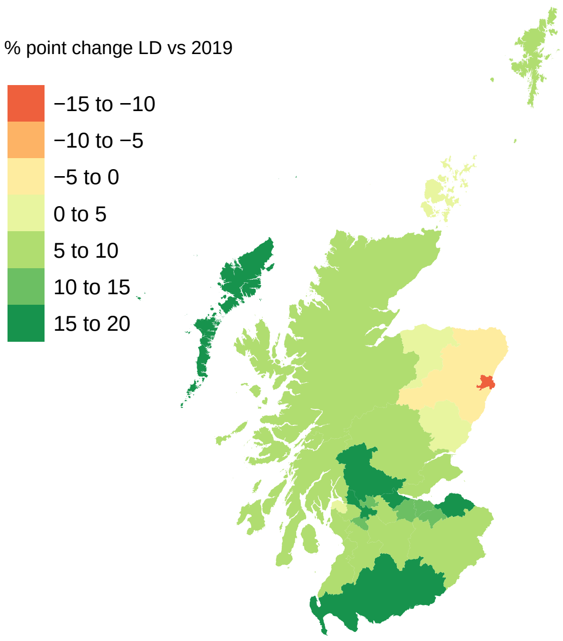


### First MMR % change from 2019

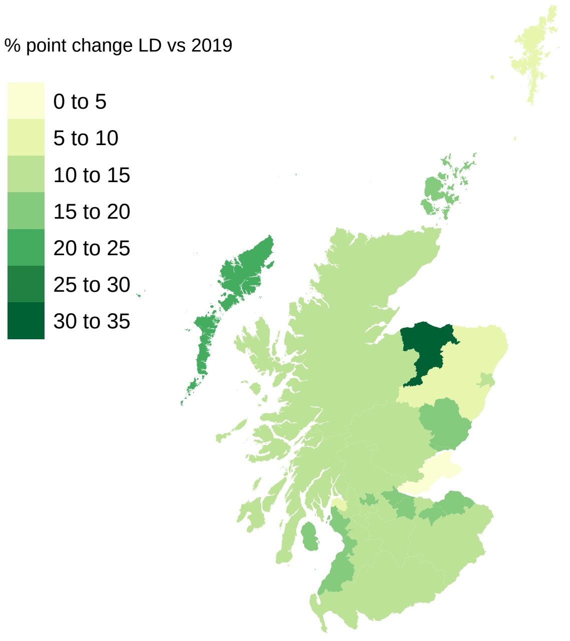


### Second MMR % change from 2019

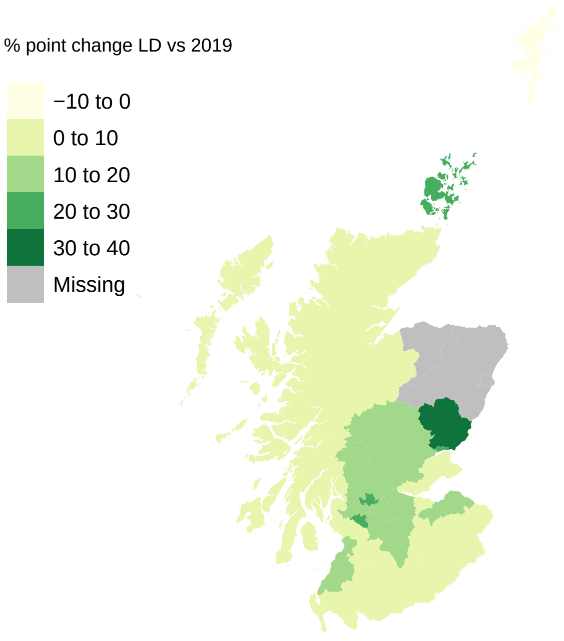
