## Supplementary material for "Uptake of infant and pre-school immunisations in Scotland and England during the COVID-19 pandemic: an observational study of routinely collected data": Table S

**Supplementary tables**

**Supplementary Table S1**

| **% uptake within 4 weeks of eligibility**  **(number received/total eligible)** | | | | | |
| --- | --- | --- | --- | --- | --- |
| **Time period** | **First 6in1** | **Second 6in1** | **Third 6in1** | **First MMR** | **Second MMR** |
| 2019 | 94  (47567/50609) | 84.8  (43221/50975) | 73  (37266/51083) | 65.2  (33935/52015) | 51.8  (25844/49940) |
| Jan-20 | 94.4  (3393/3593) | 84.1  (3101/3689) | 70.9  (2916/4112) | 68.4  (2578/3767) | 54.7  (2151/3934) |
| Feb-20 | 94.3  (4079/4325) | 86.7  (3878/4472) | 74  (3403/4600) | 69.8  (3309/4739) | 59.2  (2845/4804) |
| W/B 02-MAR-20 | 92.8  (813/876) | 84.6  (729/862) | 73.4  (628/856) | 67.2  (636/947) | 54.3  (491/904) |
| W/B 09-MAR-20 | 93.4  (883/945) | 84.3  (763/905) | 72.4  (624/862) | 66.1  (656/993) | 51  (474/930) |
| W/B 16-MAR-20 | 91.5  (935/1022) | 82.5  (635/770) | 73.1  (705/964) | 65.3  (603/924) | 46.5  (429/923) |
| W/B 23-MAR-20 | 93.2  (846/908) | 83.7  (761/909) | 73.3  (644/879) | 64.4  (588/913) | 46.9  (430/916) |
| W/B 30-MAR-20 | 95  (864/909) | 85.5  (749/876) | 75.5  (651/862) | 71.1  (692/973) | 48.6  (472/972) |
| W/B 06-APR-20 | 92.6  (892/963) | 86.9  (821/945) | 76  (688/905) | 69.7  (636/912) | 56  (509/909) |
| W/B 13-APR-20 | 93.9  (835/889) | 87.2  (891/1022) | 77  (593/770) | 76.9  (749/974) | 58.4  (558/955) |
| W/B 20-APR-20 | 95.1  (851/895) | 88.7  (805/908) | 78  (709/909) | 77.5  (732/945) | 63.3  (567/896) |
| W/B 27-APR-20 | 96  (881/918) | 91  (827/909) | 79.8  (699/876) | 80.3  (789/982) | 65.6  (563/858) |
| W/B 04-MAY-20 | 94.7  (838/885) | 88.1  (848/963) | 80  (756/945) | 80.4  (781/971) | 66.9  (590/882) |
| W/B 11-MAY-20 | 96.3  (894/928) | 89.1  (792/889) | 82.7  (845/1022) | 82.8  (815/984) | 70.6  (653/925) |
| W/B 18-MAY-20 | 95.5  (857/897) | 91.6  (820/895) | 84.7  (769/908) | 81.3  (784/964) | 70.6  (602/853) |
| W/B 25-MAY-20 | 96.2  (840/873) | 92.7  (851/918) | 86.1  (783/909) | 83.2  (820/986) | 74.2  (636/857) |
| W/B 01-JUN-20 | 97.1  (835/860) | 91  (805/885) | 83.1  (800/963) | 80.9  (728/900) | 73.1  602/824) |
| W/B 08-JUN-20 | 95  (899/946) | 93.5  (868/928) | 82.9  (737/889) | 81.9  (801/978) | 76.5  (739/966) |
| W/B 15-JUN-20 | 95.9  (880/918) | 91  (816/897) | 84.7  (758/895) | 81.7  (793/971) | 70.8  (644/909) |
| W/B 22-JUN-20 | 95.5  (804/842) | 92.1  (804/873) | 85.6  (786/918) | 80.6  (789/979) | 70.4  (650/923) |
| W/B 29-JUN-20 | 94.8  (795/839) | 92.9  (799/860) | 86.6  (766/885) | 79.3  (783/987) | 68.1  (608/893) |
| W/B 06-JUL-20 | 96.4  (863/895) | 90.5  (856/946) | 87.6  (813/928) | 76.8  (788/1026) | 71.4  (637/892) |
| W/B 13-JUL-20 | 95.2  (894/939) | 90.8  (834/918) | 85.1  (763/897) | 80.4  (805/1001) | 69.5  (629/905) |
| W/B 20-JUL-20 | 96.8  (864/893) | 89  (749/842) | 85.1  (743/873) | 80  (813/1016) | 68.2  (597/875) |
| W/B 27-JUL-20 | 94.6  (885/936) | 89  (747/839) | 85.6  (736/860) | 79.5  (796/1001) | 66  (617/935) |
| W/B 03-AUG-20 | 95  (916/964) | 90.4  (809/895) | 83.2  (787/946) | 78.3  (808/1032) | 70  (656/937) |
| W/B 10-AUG-20 | 95.1  (847/891) | 89.1  (837/939) | 82.4  (756/918) | 77.2  (761/986) | 65.9  (602/913) |
| W/B 17-AUG-20 | 95  (914/962) | 89.5  (799/893) | 80.6  (679/842) | 75.9  (749/987) | 65.8  (571/868) |
| W/B 24-AUG-20 | 94.7  (946/999) | 87.1  (815/936) | 81.3  (682/839) | 73.8  (721/977) | 61.4  (551/897) |
| W/B 31-AUG-20 | 94.9  (885/933) | 88.4  (852/964) | 81.2  (727/895) | 74  (709/958) | 61  (540/885) |
| W/B 07-SEP-20 | 95.1  (939/987) | 89.1  (794/891) | 79.7  (748/939) | 72.9  (725/995) | 63.5  (589/927) |
| W/B 14-SEP-20 | 94  (885/941) | 87.6  (843/962) | 79.3  (708/893) | 74  (751/1015) | 61.9  (569/919) |
| W/B 21-SEP-20 | 93.5  (864/924) | 88.3  (882/999) | 77.4  (724/936) | 73.4  (782/1066) | 58.8  (549/933) |
| W/B 28-SEP-20 | 94.5  (879/930) | 88.7  (828/933) | 77.2  (744/964) | 71.2  (734/1031) | 59.3  (544/917) |

Table S1: Percentage uptake of each immunisation by year (2019), month (Jan and Feb 2020) or week as per data availability. W/B = week beginning.

**Supplementary Tables S2**

A-E Percentage uptake, percent point change in uptake compared to 2019 and significance level for this change for each HSCP at each time-period. Each table shows results for a different immunisation. Results were considered significant if *p*-value <0.05 and 95% CI did not include 1. Statistically significant p values are shaded green and significant results for the 2019-LD comparisons are plotted on Figure 2. HSCP = Health and Social Care Partnership, OR = odds ratio, CI = confidence interval, NA = not applicable, PreLD = pre lockdown, LD = lockdown, PostLD = post lockdown. *p*-value rounded to 2 decimal places.

**A First dose 6in1**

| **HSCP** | **Time period** | **% uptake (within 4 weeks)** | **Number received** | **Number eligible** | **% point change from 2019** | **OR compared to 2019 (95% CI)** | **p value** |
| --- | --- | --- | --- | --- | --- | --- | --- |
| Aberdeen City | 2019 | 93.2 | 2077 | 2228 | NA | NA | NA |
|  | PreLD | 90.5 | 382 | 425 | -2.7 | 0.65  (0.45-0.92) | 0.02 |
|  | LD | 94.2 | 696 | 739 | 1 | 1.18  (0.83-1.67) | 0.36 |
|  | PostLD | 91.6 | 334 | 365 | -1.6 | 0.78  (0.52-1.17) | 0.24 |
| Aberdeenshire | 2019 | 96.6 | 2452 | 2538 | NA | NA | NA |
|  | PreLD | 95.8 | 513 | 540 | -0.8 | 0.67  (0.43-1.04) | 0.07 |
|  | LD | 95.7 | 793 | 830 | -0.9 | 0.75  (0.51-1.11) | 0.16 |
|  | PostLD | 92.1 | 354 | 385 | -4.5 | 0.4  (0.26-0.61) | <0.001 |
| Angus | 2019 | 92.7 | 939 | 1013 | NA | NA | NA |
|  | PreLD | 92.4 | 205 | 219 | -0.3 | 1.15  (0.64-2.08) | 0.63 |
|  | LD | 94.5 | 312 | 330 | 1.8 | 1.37  (0.8-2.32) | 0.25 |
|  | PostLD | 93.9 | 158 | 169 | 1.2 | 1.13  (0.59-2.18) | 0.71 |
| Argyll and Bute | 2019 | 93.3 | 608 | 652 | NA | NA | NA |
|  | PreLD | 93.4 | 140 | 150 | 0.1 | 1.01  (0.5-2.06) | 0.97 |
|  | LD | 96.8 | 234 | 243 | 3.5 | 1.88  (0.9-3.92) | 0.09 |
|  | PostLD | 91.3 | 96 | 106 | -2 | 0.69  (0.34-1.43) | 0.32 |
| Clackmannanshire and Stirling | 2019 | 93.3 | 1092 | 1170 | NA | NA | NA |
|  | PreLD | 92.9 | 272 | 288 | -0.4 | 1.21  (0.7-2.11) | 0.49 |
|  | LD | 97.3 | 398 | 408 | 4 | 2.84  (1.46-5.55) | <0.001 |
|  | PostLD | 95.9 | 191 | 199 | 2.6 | 1.71  (0.81-3.59) | 0.16 |
| Dumfries and Galloway | 2019 | 94.4 | 1116 | 1182 | NA | NA | NA |
|  | PreLD | 91.1 | 231 | 247 | -3.3 | 0.85  (0.49-1.5) | 0.58 |
|  | LD | 94.6 | 404 | 429 | 0.2 | 0.96  (0.59-1.54) | 0.85 |
|  | PostLD | 93.7 | 194 | 206 | -0.7 | 0.96  (0.51-1.8) | 0.89 |
| Dundee City | 2019 | 91.4 | 1289 | 1411 | NA | NA | NA |
|  | PreLD | 89.1 | 266 | 290 | -2.3 | 1.05  (0.66-1.66) | 0.84 |
|  | LD | 92.5 | 426 | 461 | 1.1 | 1.15  (0.78-1.7) | 0.48 |
|  | PostLD | 92.1 | 206 | 225 | 0.7 | 1.03  (0.62-1.7) | 0.92 |
| East Ayrshire | 2019 | 96.9 | 1122 | 1158 | NA | NA | NA |
|  | PreLD | 95.1 | 241 | 252 | -1.8 | 0.7  (0.35-1.4) | 0.32 |
|  | LD | 97.4 | 420 | 431 | 0.5 | 1.23  (0.62-2.43) | 0.56 |
|  | PostLD | 94.1 | 181 | 193 | -2.8 | 0.48  (0.25-0.95) | 0.03 |
| East Dunbartonshire | 2019 | 96.1 | 958 | 997 | NA | NA | NA |
|  | PreLD | 95.7 | 187 | 193 | -0.4 | 1.27  (0.53-3.04) | 0.59 |
|  | LD | 98.6 | 339 | 344 | 2.5 | 2.76  (1.08-7.06) | 0.03 |
|  | PostLD | 99.4 | 154 | 155 | 3.3 | 6.27  (0.86-45.96) | 0.07 |
| East Lothian | 2019 | 94.6 | 1031 | 1090 | NA | NA | NA |
|  | PreLD | 93.9 | 194 | 204 | -0.7 | 1.11  (0.56-2.21) | 0.77 |
|  | LD | 95.8 | 330 | 346 | 1.2 | 1.18  (0.67-2.08) | 0.57 |
|  | PostLD | 95.3 | 194 | 203 | 0.7 | 1.23  (0.6-2.53) | 0.57 |
| East Renfrewshire | 2019 | 97.4 | 868 | 891 | NA | NA | NA |
|  | PreLD | 96 | 193 | 201 | -1.4 | 0.64  (0.28-1.45) | 0.28 |
|  | LD | 98.3 | 290 | 295 | 0.9 | 1.54  (0.58-4.08) | 0.39 |
|  | PostLD | 97.4 | 149 | 153 | 0 | 0.99  (0.34-2.89) | 0.98 |
| Edinburgh | 2019 | 92.5 | 4130 | 4464 | NA | NA | NA |
|  | PreLD | 89.9 | 883 | 964 | -2.6 | 0.88  (0.68-1.14) | 0.33 |
|  | LD | 94.2 | 1517 | 1613 | 1.7 | 1.28  (1.01-1.62) | 0.04 |
|  | PostLD | 94.4 | 751 | 796 | 1.9 | 1.35  (0.98-1.86) | 0.07 |
| Falkirk | 2019 | 93.7 | 1382 | 1475 | NA | NA | NA |
|  | PreLD | 95 | 291 | 308 | 1.3 | 1.15  (0.68-1.96) | 0.6 |
|  | LD | 96.2 | 513 | 534 | 2.5 | 1.64  (1.01-2.67) | 0.04 |
|  | PostLD | 95.8 | 245 | 256 | 2.1 | 1.5  (0.79-2.84) | 0.21 |
| Fife | 2019 | 95.2 | 3234 | 3396 | NA | NA | NA |
|  | PreLD | 93.4 | 700 | 744 | -1.8 | 0.8  (0.57-1.12) | 0.19 |
|  | LD | 93.7 | 1078 | 1150 | -1.5 | 0.75  (0.56-1) | 0.05 |
|  | PostLD | 92.3 | 520 | 562 | -2.9 | 0.62  (0.44-0.88) | 0.01 |
| Glasgow City | 2019 | 91 | 5855 | 6435 | NA | NA | NA |
|  | PreLD | 92.5 | 1254 | 1346 | 1.5 | 1.35  (1.07-1.7) | 0.01 |
|  | LD | 95 | 2029 | 2136 | 4 | 1.88  (1.52-2.32) | <0.001 |
|  | PostLD | 94.8 | 1069 | 1128 | 3.8 | 1.79  (1.36-2.36) | <0.001 |
| Highland | 2019 | 91.4 | 1821 | 1992 | NA | NA | NA |
|  | PreLD | 88.8 | 389 | 433 | -2.6 | 0.83  (0.59-1.18) | 0.3 |
|  | LD | 92.8 | 587 | 635 | 1.4 | 1.15  (0.82-1.6) | 0.42 |
|  | PostLD | 92.9 | 336 | 360 | 1.5 | 1.31  (0.84-2.05) | 0.23 |
| Inverclyde | 2019 | 97.8 | 618 | 632 | NA | NA | NA |
|  | PreLD | 96.9 | 146 | 151 | -0.9 | 0.66  (0.23-1.87) | 0.43 |
|  | LD | 95.1 | 217 | 227 | -2.7 | 0.49  (0.22-1.12) | 0.09 |
|  | PostLD | 97.1 | 96 | 99 | -0.7 | 0.72  (0.2-2.57) | 0.62 |
| Midlothian | 2019 | 94.1 | 1051 | 1117 | NA | NA | NA |
|  | PreLD | 94.4 | 212 | 226 | 0.3 | 0.95  (0.52-1.72) | 0.87 |
|  | LD | 95.5 | 336 | 352 | 1.4 | 1.32  (0.75-2.31) | 0.33 |
|  | PostLD | 95.8 | 191 | 200 | 1.7 | 1.33  (0.65-2.72) | 0.43 |
| Moray | 2019 | 95.7 | 788 | 823 | NA | NA | NA |
|  | PreLD | 95.2 | 209 | 222 | -0.5 | 0.71  (0.37-1.37) | 0.31 |
|  | LD | 95.6 | 295 | 310 | -0.1 | 0.87  (0.47-1.62) | 0.67 |
|  | PostLD | 97.4 | 141 | 145 | 1.7 | 1.57  (0.55-4.47) | 0.4 |
| North Ayrshire | 2019 | 96.8 | 1082 | 1118 | NA | NA | NA |
|  | PreLD | 98.2 | 261 | 270 | 1.4 | 0.96  (0.46-2.03) | 0.92 |
|  | LD | 96 | 371 | 386 | -0.8 | 0.82  (0.45-1.52) | 0.53 |
|  | PostLD | 93.7 | 182 | 193 | -3.1 | 0.55  (0.28-1.1) | 0.09 |
| North Lanarkshire | 2019 | 95.1 | 3337 | 3510 | NA | NA | NA |
|  | PreLD | 95.6 | 696 | 724 | 0.5 | 1.29  (0.86-1.94) | 0.22 |
|  | LD | 95.5 | 1049 | 1098 | 0.4 | 1.11  (0.8-1.54) | 0.53 |
|  | PostLD | 97.1 | 609 | 627 | 2 | 1.75  (1.07-2.87) | 0.03 |
| Orkney Islands | 2019 | 92.6 | 189 | 204 | NA | NA | NA |
|  | PreLD | 100 | 28 | 28 | 7.4 | 1.16  (0.55-2.44) | 0.69 |
|  | LD | 89.7 | 53 | 59 | -2.9 | 1.27  (0.81-2.01) | 0.3 |
|  | PostLD | 100 | 37 | 37 | 7.4 | 0.8  (0.48-1.33) | 0.39 |
| Perth and Kinross | 2019 | 90.6 | 1124 | 1240 | NA | NA | NA |
|  | PreLD | 87.4 | 225 | 255 | -3.2 | 0.77  (0.51-1.19) | 0.24 |
|  | LD | 92.5 | 411 | 444 | 1.9 | 1.29  (0.86-1.92) | 0.22 |
|  | PostLD | 93.2 | 191 | 206 | 2.6 | 1.31  (0.75-2.3) | 0.34 |
| Renfrewshire | 2019 | 95.7 | 1617 | 1689 | NA | NA | NA |
|  | PreLD | 97.8 | 377 | 385 | 2.1 | 2.1  (1-4.39) | 0.05 |
|  | LD | 97.6 | 587 | 602 | 1.9 | 1.74  (0.99-3.06) | 0.05 |
|  | PostLD | 96.6 | 239 | 248 | 0.9 | 1.18  (0.58-2.4) | 0.64 |
| Scottish Borders | 2019 | 94.3 | 838 | 889 | NA | NA | NA |
|  | PreLD | 93.9 | 183 | 193 | -0.4 | 1.11  (0.56-2.23) | 0.76 |
|  | LD | 94.9 | 291 | 306 | 0.6 | 1.18  (0.65-2.13) | 0.58 |
|  | PostLD | 95.8 | 142 | 148 | 1.5 | 1.44  (0.61-3.42) | 0.41 |
| Shetland Islands | 2019 | 95.3 | 203 | 213 | NA | NA | NA |
|  | PreLD | 86.4 | 22 | 25 | -8.9 | 0.36  (0.09-1.41) | 0.14 |
|  | LD | 89 | 54 | 60 | -6.3 | 1.27  (0.81-2.01) | 0.3 |
|  | PostLD | 89.6 | 33 | 37 | -5.8 | 0.8  (0.48-1.33) | 0.39 |
| South Ayrshire | 2019 | 96.5 | 844 | 875 | NA | NA | NA |
|  | PreLD | 94.1 | 187 | 198 | -2.4 | 0.62  (0.31-1.26) | 0.19 |
|  | LD | 99.2 | 274 | 277 | 2.7 | 3.35  (1.02-11.06) | 0.05 |
|  | PostLD | 97.4 | 148 | 152 | 0.9 | 1.36  (0.47-3.91) | 0.57 |
| South Lanarkshire | 2019 | 96 | 3066 | 3194 | NA | NA | NA |
|  | PreLD | 94.6 | 622 | 646 | -1.4 | 1.08  (0.69-1.69) | 0.73 |
|  | LD | 97.4 | 1084 | 1113 | 1.4 | 1.56  (1.04-2.35) | 0.03 |
|  | PostLD | 96.4 | 484 | 502 | 0.4 | 1.12  (0.68-1.86) | 0.65 |
| West Dunbartonshire | 2019 | 92.1 | 796 | 864 | NA | NA | NA |
|  | PreLD | 90.3 | 158 | 172 | -1.8 | 0.96  (0.53-1.76) | 0.9 |
|  | LD | 93.4 | 274 | 292 | 1.3 | 1.3  (0.76-2.23) | 0.34 |
|  | PostLD | 95.8 | 144 | 150 | 3.7 | 2.05  (0.87-4.81) | 0.1 |
| West Lothian | 2019 | 94.7 | 1754 | 1852 | NA | NA | NA |
|  | PreLD | 93.9 | 342 | 363 | -0.8 | 0.91  (0.56-1.48) | 0.7 |
|  | LD | 95.7 | 547 | 571 | 1 | 1.27  (0.81-2.01) | 0.3 |
|  | PostLD | 93.4 | 272 | 291 | -1.3 | 0.8  (0.48-1.33) | 0.39 |
| Western Isles | 2019 | 95.4 | 188 | 197 | NA | NA | NA |
|  | PreLD | 95.8 | 33 | 35 | 0.4 | 0.79  (0.16-3.82) | 0.77 |
|  | LD | 94.6 | 56 | 59 | -0.8 | 1.18  (0.83-1.67) | 0.36 |
|  | PostLD | 95.4 | 23 | 24 | 0 | 0.78  (0.52-1.17) | 0.24 |

**B Second dose 6in1**

| **HSCP** | **Time period** | **% uptake (within 4 weeks)** | **Number received** | **Number eligible** | **% point change from 2019** | **OR compared to 2019**  **(95% CI)** | **p value** |
| --- | --- | --- | --- | --- | --- | --- | --- |
| Aberdeen City | 2019 | 84.3 | 1891 | 2244 | NA | NA | NA |
|  | PreLD | 74.7 | 358 | 459 | -9.6 | 0.66  (0.52-0.85) | <0.001 |
|  | LD | 81.9 | 603 | 733 | -2.4 | 0.87  (0.69-1.08) | 0.2 |
|  | PostLD | 82 | 281 | 344 | -2.3 | 0.83  (0.62-1.12) | 0.23 |
| Aberdeenshire | 2019 | 93.3 | 2411 | 2584 | NA | NA | NA |
|  | PreLD | 87.4 | 470 | 530 | -5.9 | 0.56  (0.41-0.77) | <0.001 |
|  | LD | 92.9 | 753 | 811 | -0.4 | 0.93  (0.68-1.27) | 0.65 |
|  | PostLD | 90.7 | 365 | 404 | -2.6 | 0.67  (0.47-0.97) | 0.03 |
| Angus | 2019 | 82.7 | 843 | 1019 | NA | NA | NA |
|  | PreLD | 80 | 188 | 223 | -2.7 | 1.12  (0.75-1.67) | 0.57 |
|  | LD | 87.6 | 288 | 330 | 4.9 | 1.43  (1-2.06) | 0.05 |
|  | PostLD | 85.7 | 136 | 160 | 3 | 1.18  (0.74-1.88) | 0.48 |
| Argyll and Bute | 2019 | 81.7 | 523 | 640 | NA | NA | NA |
|  | PreLD | 84.4 | 137 | 167 | 2.7 | 1.02  (0.66-1.59) | 0.92 |
|  | LD | 89.3 | 214 | 240 | 7.6 | 1.84  (1.17-2.9) | 0.01 |
|  | PostLD | 83.8 | 86 | 107 | 2.1 | 0.92  (0.55-1.54) | 0.74 |
| Clackmannanshire and Stirling | 2019 | 84.9 | 1003 | 1182 | NA | NA | NA |
|  | PreLD | 91.2 | 231 | 259 | 6.3 | (1.47  (0.96-2.25) | 0.07 |
|  | LD | 92.8 | 416 | 448 | 7.9 | 2.32  (1.57-3.44) | <0.001 |
|  | PostLD | 92.5 | 173 | 188 | 7.6 | 2.06  (1.19-3.57) | 0.01 |
| Dumfries and Galloway | 2019 | 82 | 978 | 1193 | NA | NA | NA |
|  | PreLD | 87.2 | 217 | 251 | 5.2 | 1.4  (0.95-2.07) | 0.09 |
|  | LD | 89.5 | 363 | 409 | 7.5 | 1.73  (1.23-2.44) | <0.001 |
|  | PostLD | 90.3 | 197 | 218 | 8.3 | 2.06  (1.28-3.31) | <0.001 |
| Dundee City | 2019 | 79.6 | 1133 | 1423 | NA | NA | NA |
|  | PreLD | 75.9 | 226 | 284 | -3.7 | 1  (0.73-1.37) | 0.99 |
|  | LD | 83.2 | 385 | 463 | 3.6 | 1.26  (0.96-1.66) | 0.1 |
|  | PostLD | 83.9 | 199 | 238 | 4.3 | 1.31  (0.91-1.88) | 0.15 |
| East Ayrshire | 2019 | 89 | 1052 | 1182 | NA | NA | NA |
|  | PreLD | 88.2 | 214 | 242 | -0.8 | 0.94  (0.61-1.46) | 0.8 |
|  | LD | 92.7 | 400 | 429 | 3.7 | 1.7  (1.12-2.59) | 0.01 |
|  | PostLD | 92.8 | 187 | 202 | 3.8 | 1.54  (0.88-2.69) | 0.13 |
| East Dunbartonshire | 2019 | 88.9 | 893 | 1005 | NA | NA | NA |
|  | PreLD | 96.4 | 186 | 196 | 7.5 | 2.33  (1.2-4.54) | 0.01 |
|  | LD | 97 | 331 | 342 | 8.1 | 3.77  (2.01-7.1) | <0.001 |
|  | PostLD | 95.4 | 168 | 176 | 6.5 | 2.63  (1.26-5.5) | 0.01 |
| East Lothian | 2019 | 83.8 | 932 | 1112 | NA | NA | NA |
|  | PreLD | 87.1 | 169 | 198 | 3.3 | 1.13  (0.74-1.72) | 0.59 |
|  | LD | 90.9 | 326 | 360 | 7.1 | 1.85  (1.26-2.73) | <0.001 |
|  | PostLD | 88.8 | 152 | 170 | 5 | 1.63  (0.98-2.73) | 0.06 |
| East Renfrewshire | 2019 | 91.6 | 834 | 910 | NA | NA | NA |
|  | PreLD | 95.8 | 171 | 181 | 4.2 | 1.56  (0.79-3.07) | 0.2 |
|  | LD | 96.6 | 282 | 292 | 5 | 2.57  (1.31-5.04) | 0.01 |
|  | PostLD | 96.3 | 156 | 162 | 4.7 | 2.37  (1.01-5.54) | 0.05 |
| Edinburgh | 2019 | 81 | 3650 | 4505 | NA | NA | NA |
|  | PreLD | 83.3 | 814 | 980 | 2.3 | 1.15  (0.96-1.38) | 0.14 |
|  | LD | 87.7 | 1387 | 1582 | 6.7 | 1.67  (1.41-1.97) | <0.001 |
|  | PostLD | 83.9 | 679 | 810 | 2.9 | 1.21  (0.99-1.48) | 0.06 |
| Falkirk | 2019 | 82.3 | 1243 | 1511 | NA | NA | NA |
|  | PreLD | 82.8 | 260 | 315 | 0.5 | 1.02  (0.74-1.4) | 0.91 |
|  | LD | 91.7 | 473 | 517 | 9.4 | 2.32  (1.66-3.24) | <0.001 |
|  | PostLD | 92.1 | 231 | 250 | 9.8 | 2.62  (1.61-4.26) | <0.001 |
| Fife | 2019 | 86.8 | 2966 | 3418 | NA | NA | NA |
|  | PreLD | 83.7 | 627 | 732 | -3.1 | 0.91  (0.72-1.14) | 0.42 |
|  | LD | 89.6 | 1030 | 1151 | 2.8 | 1.3  (1.05-1.6) | 0.02 |
|  | PostLD | 90.2 | 509 | 562 | 3.4 | 1.46  (1.08-1.98) | 0.01 |
| Glasgow City | 2019 | 80.2 | 5196 | 6480 | NA | NA | NA |
|  | PreLD | 83.2 | 1116 | 1347 | 3 | 1.19  (1.02-1.39) | 0.02 |
|  | LD | 89.1 | 1894 | 2125 | 8.9 | 2.03  (1.74-2.35) | <0.001 |
|  | PostLD | 90.8 | 971 | 1069 | 10.6 | 2.45  (1.97-3.04) | <0.001 |
| Highland | 2019 | 80.3 | 1600 | 1992 | NA | NA | NA |
|  | PreLD | 75.5 | 332 | 419 | -4.8 | 0.93  (0.72-1.21) | 0.61 |
|  | LD | 84.8 | 565 | 669 | 4.5 | 1.33  (1.05-1.69) | 0.02 |
|  | PostLD | 80.1 | 258 | 321 | -0.2 | 1  (0.75-1.35) | 0.98 |
| Inverclyde | 2019 | 91.9 | 575 | 626 | NA | NA | NA |
|  | PreLD | 91.2 | 142 | 156 | -0.7 | 0.9  (0.48-1.67) | 0.74 |
|  | LD | 91.6 | 207 | 226 | -0.3 | 0.97  (0.56-1.68) | 0.9 |
|  | PostLD | 88.7 | 91 | 101 | -3.2 | 0.81  (0.4-1.65) | 0.56 |
| Midlothian | 2019 | 83.3 | 935 | 1122 | NA | NA | NA |
|  | PreLD | 84.1 | 181 | 218 | 0.8 | 0.98  (0.66-1.44) | 0.91 |
|  | LD | 86.9 | 308 | 354 | 3.6 | 1.34  (0.95-1.9) | 0.1 |
|  | PostLD | 93.9 | 172 | 183 | 10.6 | 3.13  (1.67-5.87) | <0.001 |
| Moray | 2019 | 90.4 | 751 | 831 | NA | NA | NA |
|  | PreLD | 88.8 | 171 | 197 | -1.6 | 0.7  (0.44-1.12) | 0.14 |
|  | LD | 90.2 | 279 | 311 | -0.2 | 0.93  (0.6-1.43) | 0.74 |
|  | PostLD | 94.2 | 146 | 155 | 3.8 | 1.73  (0.85-.52) | 0.13 |
| North Ayrshire | 2019 | 85.8 | 952 | 1109 | NA | NA | NA |
|  | PreLD | 82.7 | 221 | 257 | -3.1 | 1.01  (0.69-1.5) | 0.95 |
|  | LD | 88.7 | 354 | 399 | 2.9 | 1.3  (0.91-1.85) | 0.15 |
|  | PostLD | 85.4 | 171 | 201 | -0.4 | 0.94  (0.62-1.44) | 0.77 |
| North Lanarkshire | 2019 | 87.2 | 3090 | 3542 | NA | NA | NA |
|  | PreLD | 85.6 | 623 | 724 | -1.6 | 0.9  (0.72-1.14) | 0.39 |
|  | LD | 91.3 | 1037 | 1136 | 4.1 | 1.53  (1.22-1.93) | <0.001 |
|  | PostLD | 89.3 | 508 | 566 | 2.1 | 1.28  (0.96-1.71) | 0.09 |
| Orkney Islands | 2019 | 85 | 175 | 206 | NA | NA | NA |
|  | PreLD | 93.2 | 29 | 31 | 8.2 | 2.57  (0.58-11.32) | 0.21 |
|  | LD | 87 | 53 | 61 | 2 | 1.88  (1.39-2.53) | <0.001 |
|  | PostLD | 97 | 26 | 27 | 12 | 1.48  (1.02-2.14) | 0.04 |
| Perth and Kinross | 2019 | 81.8 | 1016 | 1242 | NA | NA | NA |
|  | PreLD | 76.9 | 211 | 264 | -4.9 | 0.89  (0.63-1.24) | 0.48 |
|  | LD | 85.3 | 382 | 446 | 3.5 | 1.33  (0.98-1.79) | 0.07 |
|  | PostLD | 85.5 | 171 | 200 | 3.7 | 1.31  (0.86-1.99) | 0.2 |
| Renfrewshire | 2019 | 88.2 | 1508 | 1710 | NA | NA | NA |
|  | PreLD | 90.8 | 334 | 366 | 2.6 | 1.4  (0.95-2.07) | 0.09 |
|  | LD | 95 | 593 | 623 | 6.8 | 2.65  (1.78-3.93) | <0.001 |
|  | PostLD | 92.2 | 246 | 269 | 4 | 1.43  (0.91-2.25) | 0.12 |
| Scottish Borders | 2019 | 85.6 | 740 | 864 | NA | NA | NA |
|  | PreLD | 83.3 | 178 | 212 | -2.3 | 0.88  (0.58-1.33) | 0.53 |
|  | LD | 89.6 | 283 | 315 | 4 | 1.48  (0.98-2.24) | 0.06 |
|  | PostLD | 86.8 | 134 | 153 | 1.2 | 1.18  (0.7-1.98) | 0.53 |
| Shetland Islands | 2019 | 89.5 | 197 | 220 | NA | NA | NA |
|  | PreLD | 83.8 | 21 | 25 | -5.8 | 0.61  (0.19-1.94) | 0.41 |
|  | LD | 82.8 | 56 | 68 | -6.7 | 1.88  (1.39-2.53) | <0.001 |
|  | PostLD | 77.3 | 25 | 32 | -12.2 | 1.48  (1.02-2.14) | 0.04 |
| South Ayrshire | 2019 | 92.4 | 830 | 898 | NA | NA | NA |
|  | PreLD | 89.5 | 165 | 184 | -2.9 | 0.71  (0.42-1.22) | 0.21 |
|  | LD | 95.5 | 277 | 290 | 3.1 | 1.75  (0.95-3.21) | 0.07 |
|  | PostLD | 92.3 | 121 | 132 | -0.1 | 0.9  (0.46-1.75) | 0.76 |
| South Lanarkshire | 2019 | 88.2 | 2821 | 3199 | NA | NA | NA |
|  | PreLD | 85.9 | 590 | 666 | -2.3 | 1.04  (0.8-1.35) | 0.77 |
|  | LD | 91.8 | 1014 | 1104 | 3.6 | 1.51  (1.19-1.92) | <0.001 |
|  | PostLD | 90.8 | 489 | 539 | 2.6 | 1.31  (0.96-1.79) | 0.09 |
| West Dunbartonshire | 2019 | 80.2 | 693 | 864 | NA | NA | NA |
|  | PreLD | 85.5 | 131 | 160 | 5.3 | 1.11  (0.72-1.72) | 0.63 |
|  | LD | 88.7 | 263 | 296 | 8.5 | 1.97  (1.32-2.93) | <0.001 |
|  | PostLD | 85 | 128 | 149 | 4.8 | 1.5  (0.92-2.46) | 0.1 |
| West Lothian | 2019 | 82.7 | 1525 | 1845 | NA | NA | NA |
|  | PreLD | 85.3 | 318 | 368 | 2.6 | 1.33  (0.97-1.84) | 0.08 |
|  | LD | 89.6 | 510 | 567 | 6.9 | 1.88  (1.39-2.53) | <0.001 |
|  | PostLD | 87.5 | 254 | 290 | 4.8 | 1.48  (1.02-2.14) | 0.04 |
| Western Isles | 2019 | 89.2 | 173 | 194 | NA | NA | NA |
|  | PreLD | 83.3 | 30 | 36 | -5.9 | 0.61  (0.23-1.63) | 0.32 |
|  | LD | 93.5 | 60 | 64 | 4.3 | 0.87  (0.69-1.08) | 0.2 |
|  | PostLD | 88.9 | 24 | 28 | -0.3 | 0.83  (0.62-1.12) | 0.23 |

**C Third dose 6in1**

| **HSCP** | **Time period** | **% uptake (within 4 weeks)** | **Number received** | **Number eligible** | **% point change from 2019** | **OR compared to 2019**  **(95% CI)** | **p value** |
| --- | --- | --- | --- | --- | --- | --- | --- |
| Aberdeen City | 2019 | 72.4 | 1609 | 2222 | NA | NA | NA |
|  | PreLD | 65.8 | 345 | 520 | -6.6 | 0.75  (0.61-0.92) | 0.01 |
|  | LD | 62.3 | 450 | 719 | -10.1 | 0.64  (0.53-0.76) | <0.001 |
|  | PostLD | 75 | 252 | 336 | 2.6 | 1.14  (0.88-1.49) | 0.32 |
| Aberdeenshire | 2019 | 86.8 | 2266 | 2611 | NA | NA | NA |
|  | PreLD | 82 | 446 | 536 | -4.8 | 0.75  (0.59-0.97) | 0.03 |
|  | LD | 86.1 | 721 | 838 | -0.7 | 0.94  (0.75-1.18) | 0.58 |
|  | PostLD | 80.8 | 336 | 416 | -6 | 0.64  (0.49-0.84) | <0.001 |
| Angus | 2019 | 71.2 | 738 | 1036 | NA | NA | NA |
|  | PreLD | 76 | 159 | 214 | 4.8 | 1.17  (0.84-1.63) | 0.37 |
|  | LD | 75.9 | 261 | 345 | 4.7 | 1.25  (0.95-1.66) | 0.11 |
|  | PostLD | 79 | 121 | 154 | 7.8 | 1.48  (0.98-2.23) | 0.06 |
| Argyll and Bute | 2019 | 69.3 | 443 | 639 | NA | NA | NA |
|  | PreLD | 72.9 | 120 | 170 | 3.6 | 1.06  (0.73-1.54) | 0.75 |
|  | LD | 78.2 | 178 | 230 | 8.9 | 1.51  (1.07-2.15) | 0.02 |
|  | PostLD | 78.8 | 94 | 118 | 9.5 | 1.73  (1.07-2.8) | 0.02 |
| Clackmannanshire and Stirling | 2019 | 72.3 | 860 | 1189 | NA | NA | NA |
|  | PreLD | 74.4 | 217 | 282 | 2.1 | 1.28  (0.94-1.73) | 0.12 |
|  | LD | 89 | 384 | 432 | 16.7 | 3.06  (2.21-4.24) | <0.001 |
|  | PostLD | 86.1 | 167 | 193 | 13.8 | 2.46  (1.59-3.79) | <0.001 |
| Dumfries and Galloway | 2019 | 68.5 | 805 | 1175 | NA | NA | NA |
|  | PreLD | 66.4 | 187 | 280 | -2.1 | 0.92  (0.7-1.22) | 0.58 |
|  | LD | 85.5 | 339 | 401 | 17 | 2.51  (1.87-3.38) | <0.001 |
|  | PostLD | 81 | 177 | 221 | 12.5 | 1.85  (1.3-2.63) | <0.001 |
| Dundee City | 2019 | 65.9 | 933 | 1416 | NA | NA | NA |
|  | PreLD | 62 | 213 | 325 | -3.9 | 0.98  (0.76-1.27) | 0.9 |
|  | LD | 73.2 | 326 | 442 | 7.3 | 1.45  (1.15-1.85) | <0.001 |
|  | PostLD | 72.5 | 170 | 235 | 6.6 | 1.35  (1-1.84) | 0.05 |
| East Ayrshire | 2019 | 78.5 | 930 | 1184 | NA | NA | NA |
|  | PreLD | 78.4 | 204 | 260 | -0.1 | 0.99  (0.72-1.38) | 0.98 |
|  | LD | 84.9 | 357 | 420 | 6.4 | 1.55  (1.14-2.09) | <0.001 |
|  | PostLD | 87 | 172 | 198 | 8.5 | 1.81  (1.17-2.79) | 0.01 |
| East Dunbartonshire | 2019 | 79.3 | 808 | 1019 | NA | NA | NA |
|  | PreLD | 86.3 | 180 | 213 | 7 | 1.42  (0.95-2.13) | 0.08 |
|  | LD | 93.2 | 304 | 326 | 13.9 | 3.61  (2.28-5.71) | <0.001 |
|  | PostLD | 92.4 | 152 | 165 | 13.1 | 3.05  (1.7-5.49) | <0.001 |
| East Lothian | 2019 | 70.2 | 786 | 1119 | NA | NA | NA |
|  | PreLD | 67.9 | 153 | 225 | -2.3 | 0.9  (0.66-1.23) | 0.5 |
|  | LD | 86.9 | 298 | 345 | 16.7 | 2.69  (1.92-3.75) | <0.001 |
|  | PostLD | 82.9 | 140 | 167 | 12.7 | 2.2  (1.43-3.38) | <0.001 |
| East Renfrewshire | 2019 | 85.3 | 795 | 932 | NA | NA | NA |
|  | PreLD | 84.5 | 145 | 167 | -0.8 | 1.14  (0.7-1.84) | 0.61 |
|  | LD | 95.6 | 294 | 310 | 10.3 | 3.17  (1.85-5.41) | <0.001 |
|  | PostLD | 89.6 | 134 | 147 | 4.3 | 1.78  (0.98-3.23) | 0.06 |
| Edinburgh | 2019 | 68 | 3058 | 4494 | NA | NA | NA |
|  | PreLD | 68.1 | 721 | 1043 | 0.1 | 1.05  (0.91-1.22) | 0.5 |
|  | LD | 80.7 | 1253 | 1545 | 12.7 | 2.02  (1.75-2.32) | <0.001 |
|  | PostLD | 75.8 | 583 | 771 | 7.8 | 1.46  (1.22-1.74) | <0.001 |
| Falkirk | 2019 | 65.5 | 969 | 1480 | NA | NA | NA |
|  | PreLD | 73.7 | 241 | 337 | 8.2 | 1.32  (1.02-1.72) | 0.03 |
|  | LD | 82 | 412 | 503 | 16.5 | 2.39  (1.86-3.07) | <0.001 |
|  | PostLD | 83.3 | 212 | 254 | 17.8 | 2.66  (1.88-3.77) | <0.001 |
| Fife | 2019 | 75 | 2571 | 3426 | NA | NA | NA |
|  | PreLD | 73.5 | 556 | 768 | -1.5 | 0.87  (0.73-1.04) | 0.13 |
|  | LD | 81.7 | 940 | 1151 | 6.7 | 1.48  (1.25-1.75) | <0.001 |
|  | PostLD | 86.4 | 484 | 561 | 11.4 | 2.09  (1.62-2.69) | <0.001 |
| Glasgow City | 2019 | 68.7 | 4454 | 6484 | NA | NA | NA |
|  | PreLD | 72.9 | 1011 | 1424 | 4.2 | 1.12  (0.98-1.27) | 0.09 |
|  | LD | 83.9 | 1796 | 2140 | 15.2 | 2.38  (2.1-2.7) | <0.001 |
|  | PostLD | 84.4 | 842 | 998 | 15.7 | 2.46  (2.06-2.94) | <0.001 |
| Highland | 2019 | 67.5 | 1340 | 1984 | NA | NA | NA |
|  | PreLD | 61.4 | 279 | 461 | -6.1 | 0.74  (0.6-0.91) | <0.001 |
|  | LD | 73.1 | 480 | 653 | 5.6 | 1.33  (1.09-1.62) | <0.001 |
|  | PostLD | 63.9 | 199 | 311 | -3.6 | 0.85  (0.67-1.1) | 0.22 |
| Inverclyde | 2019 | 84.8 | 542 | 639 | NA | NA | NA |
|  | PreLD | 78.6 | 121 | 151 | -6.2 | 0.72  (0.46-1.14) | 0.16 |
|  | LD | 87.4 | 206 | 238 | 2.6 | 1.15  (0.75-1.77) | 0.52 |
|  | PostLD | 84.6 | 91 | 105 | -0.2 | 1.16  (0.64-2.13) | 0.62 |
| Midlothian | 2019 | 69 | 791 | 1146 | NA | NA | NA |
|  | PreLD | 66.6 | 151 | 222 | -2.4 | 0.95  (0.7-1.3) | 0.77 |
|  | LD | 79.7 | 288 | 360 | 10.7 | 1.8  (1.35-2.39) | <0.001 |
|  | PostLD | 82.4 | 143 | 173 | 13.4 | 2.14  (1.42-3.23) | <0.001 |
| Moray | 2019 | 80.5 | 680 | 845 | NA | NA | NA |
|  | PreLD | 66.2 | 129 | 188 | -14.3 | 0.53  (0.37-0.75) | <0.001 |
|  | LD | 80.9 | 259 | 320 | 0.4 | 1.03  (0.74-1.43) | 0.86 |
|  | PostLD | 86.8 | 134 | 155 | 6.3 | 1.55  (0.95-2.53) | 0.08 |
| North Ayrshire | 2019 | 71.1 | 781 | 1099 | NA | NA | NA |
|  | PreLD | 74.3 | 198 | 268 | 3.2 | 1.15  (0.85-1.56) | 0.36 |
|  | LD | 80.2 | 334 | 417 | 9.1 | 1.55  (0.95-2.53) | 0.08 |
|  | PostLD | 67 | 117 | 175 | -4.1 | 0.82  (0.58-1.15) | 0.26 |
| North Lanarkshire | 2019 | 74.5 | 2651 | 3558 | NA | NA | NA |
|  | PreLD | 77.7 | 579 | 758 | 3.2 | 1.11  (0.92-1.33) | 0.28 |
|  | LD | 83.6 | 959 | 1148 | 9.1 | 1.74  (1.46-2.06) | <0.001 |
|  | PostLD | 77.8 | 404 | 517 | 3.3 | 1.22  (0.98-1.53) | 0.07 |
| Orkney Islands | 2019 | 76 | 155 | 204 | NA | NA | NA |
|  | PreLD | 82.2 | 27 | 33 | 6.2 | 1.42  (0.56-3.65) | 0.46 |
|  | LD | 79.2 | 39 | 51 | 3.2 | 2.22  (1.76-2.79) | <0.001 |
|  | PostLD | 94.4 | 28 | 29 | 18.5 | 1.74  (1.27-2.39) | <0.001 |
| Perth and Kinross | 2019 | 69.8 | 847 | 1213 | NA | NA | NA |
|  | PreLD | 57.3 | 204 | 313 | -12.5 | 0.81  (0.62-1.05) | 0.11 |
|  | LD | 75.7 | 317 | 415 | 5.9 | 1.4  (1.08-1.81) | 0.01 |
|  | PostLD | 74.8 | 166 | 219 | 5 | 1.35  (0.97-1.89) | 0.07 |
| Renfrewshire | 2019 | 78.4 | 1342 | 1712 | NA | NA | NA |
|  | PreLD | 81.3 | 307 | 386 | 2.9 | 1.07  (0.82-1.41) | 0.62 |
|  | LD | 85.1 | 537 | 630 | 6.7 | 1.59  (1.24-2.04) | <0.001 |
|  | PostLD | 84.5 | 228 | 271 | 6.1 | 1.46  (1.03-2.07) | 0.03 |
| Scottish Borders | 2019 | 76.4 | 662 | 867 | NA | NA | NA |
|  | PreLD | 68.3 | 165 | 239 | -8.1 | 0.69  (0.5-0.95) | 0.02 |
|  | LD | 83.3 | 250 | 303 | 6.9 | 1.46  (1.04-2.04) | 0.03 |
|  | PostLD | 73.5 | 103 | 137 | -2.9 | 0.94  (0.62-1.43) | 0.76 |
| Shetland Islands | 2019 | 77.2 | 169 | 219 | NA | NA | NA |
|  | PreLD | 87 | 27 | 31 | 9.8 | 2  (0.67-5.98) | 0.22 |
|  | LD | 85.2 | 56 | 65 | 8 | 2.22  (1.76-2.79) | <0.001 |
|  | PostLD | 42.8 | 13 | 31 | -34.5 | 1.74  (1.27-2.39) | <0.001 |
| South Ayrshire | 2019 | 82.1 | 760 | 926 | NA | NA | NA |
|  | PreLD | 79.6 | 149 | 180 | -2.5 | 1.05  (0.69-1.6) | 0.82 |
|  | LD | 90.5 | 271 | 301 | 8.4 | 1.97  (1.31-2.98) | <0.001 |
|  | PostLD | 85 | 103 | 122 | 2.9 | 1.18  (0.71-1.99) | 0.52 |
| South Lanarkshire | 2019 | 76.6 | 2469 | 3225 | NA | NA | NA |
|  | PreLD | 75 | 587 | 745 | -1.6 | 1.14  (0.94-1.38) | 0.19 |
|  | LD | 85.1 | 893 | 1043 | 8.5 | 1.82  (1.51-2.21) | <0.001 |
|  | PostLD | 80.6 | 438 | 545 | 4 | 1.25  (1-1.57) | 0.05 |
| West Dunbartonshire | 2019 | 67.6 | 585 | 866 | NA | NA | NA |
|  | PreLD | 75.7 | 129 | 190 | 8.1 | 1.02  (0.73-1.42) | 0.93 |
|  | LD | 84.5 | 238 | 281 | 16.9 | 2.66  (1.86-3.79) | <0.001 |
|  | PostLD | 78.8 | 116 | 147 | 11.2 | 1.8  (1.18-2.74) | 0.01 |
| West Lothian | 2019 | 67.2 | 1244 | 1850 | NA | NA | NA |
|  | PreLD | 73.7 | 266 | 382 | 6.5 | 1.12  (0.88-1.42) | 0.36 |
|  | LD | 82.1 | 487 | 594 | 14.9 | 2.22  (1.76-2.79) | <0.001 |
|  | PostLD | 77.6 | 193 | 247 | 10.4 | 1.74  (1.27-2.39) | <0.001 |
| Western Isles | 2019 | 74.1 | 149 | 201 | NA | NA | NA |
|  | PreLD | 60.8 | 18 | 30 | -13.3 | 0.52  (0.24-1.16) | 0.11 |
|  | LD | 91.6 | 62 | 69 | 17.5 | 0.64  (0.53-0.76) | <0.001 |
|  | PostLD | 76.7 | 23 | 30 | 2.6 | 1.14  (0.88-1.49) | 0.32 |

**D First dose MMR**

| **HSCP** | **Time period** | **% uptake (within 4 weeks)** | **Number received** | **Number eligible** | **% point change from 2019** | **OR compared to 2019**  **(94% CI)** | **p value** |
| --- | --- | --- | --- | --- | --- | --- | --- |
| Aberdeen City | 2019 | 55.1 | 1235 | 2243 | NA | NA | NA |
|  | PreLD | 53.4 | 259 | 501 | -1.7 | 0.87  (0.72-1.06) | 0.17 |
|  | LD | 69.3 | 553 | 795 | 14.2 | 1.87  (1.57-2.22) | <0.001 |
|  | PostLD | 71.5 | 285 | 399 | 16.4 | 2.04  (1.62-2.57) | <0.001 |
| Aberdeenshire | 2019 | 49.4 | 1383 | 2798 | NA | NA | NA |
|  | PreLD | 48 | 254 | 554 | -1.4 | 0.87  (0.72-1.04) | 0.12 |
|  | LD | 56.8 | 536 | 936 | 7.4 | 1.37  (1.18-1.59) | <0.001 |
|  | PostLD | 48.9 | 221 | 448 | -0.5 | 1  (0.82-1.22) | 0.97 |
| Angus | 2019 | 69.3 | 681 | 982 | NA | NA | NA |
|  | PreLD | 72.1 | 182 | 250 | 2.8 | 1.18  (0.87-1.61) | 0.29 |
|  | LD | 85.6 | 311 | 365 | 16.3 | 2.55  (1.85-3.5) | <0.001 |
|  | PostLD | 80 | 129 | 162 | 10.7 | 1.73  (1.15-2.59) | 0.01 |
| Argyll and Bute | 2019 | 64.8 | 440 | 679 | NA | NA | NA |
|  | PreLD | 74 | 112 | 157 | 9.2 | 1.35  (0.92-1.98) | 0.12 |
|  | LD | 78.6 | 178 | 226 | 13.8 | 2.01  (1.41-2.87) | <0.001 |
|  | PostLD | 72.9 | 74 | 102 | 8.1 | 1.44  (0.9-2.28) | 0.13 |
| Clackmannanshire and Stirling | 2019 | 70.8 | 902 | 1274 | NA | NA | NA |
|  | PreLD | 73.4 | 179 | 242 | 2.6 | 1.17  (0.86-1.6) | 0.32 |
|  | LD | 83.5 | 377 | 449 | 12.7 | 2.16  (1.63-2.86) | <0.001 |
|  | PostLD | 86.8 | 178 | 205 | 16 | 2.72  (1.78-4.15) | <0.001 |
| Dumfries and Galloway | 2019 | 72 | 907 | 1259 | NA | NA | NA |
|  | PreLD | 71.7 | 198 | 272 | -0.3 | 1.04  (0.77-1.39) | 0.8 |
|  | LD | 84 | 338 | 401 | 12 | 2.08  (1.55-2.8) | <0.001 |
|  | PostLD | 79.7 | 178 | 224 | 7.7 | 1.5  (1.06-2.12) | 0.02 |
| Dundee City | 2019 | 60.5 | 857 | 1416 | NA | NA | NA |
|  | PreLD | 60.3 | 188 | 293 | -0.2 | 1.17  (0.9-1.52) | 0.24 |
|  | LD | 76 | 388 | 510 | 15.5 | 2.07  (1.65-2.61) | <0.001 |
|  | PostLD | 74.5 | 198 | 265 | 14 | 1.93  (1.43-2.59) | <0.001 |
| East Ayrshire | 2019 | 58.9 | 703 | 1194 | NA | NA | NA |
|  | PreLD | 56.3 | 166 | 260 | -2.6 | 1.23  (0.93-1.63) | 0.14 |
|  | LD | 69 | 306 | 441 | 10.1 | 1.58  (1.25-2) | <0.001 |
|  | PostLD | 70.4 | 154 | 218 | 11.5 | 1.68  (1.23-2.3) | <0.001 |
| East Dunbartonshire | 2019 | 75.8 | 803 | 1060 | NA | NA | NA |
|  | PreLD | 79.3 | 188 | 240 | 3.5 | 1.16  (0.83-1.62) | 0.4 |
|  | LD | 91.1 | 303 | 335 | 15.3 | 3.03  (2.05-4.48) | <0.001 |
|  | PostLD | 86.8 | 162 | 187 | 11 | 2.07  (1.33-3.23) | <0.001 |
| East Lothian | 2019 | 71.2 | 800 | 1124 | NA | NA | NA |
|  | PreLD | 70.7 | 166 | 233 | -0.5 | 1  (0.73-1.37) | 0.98 |
|  | LD | 87.3 | 352 | 405 | 16.1 | 2.69  (1.96-3.69) | <0.001 |
|  | PostLD | 80.2 | 157 | 198 | 9 | 1.55  (1.07-2.24) | 0.02 |
| East Renfrewshire | 2019 | 74.6 | 745 | 998 | NA | NA | NA |
|  | PreLD | 76.6 | 157 | 199 | 2 | 1.27  (0.88-1.84) | 0.21 |
|  | LD | 84.8 | 280 | 327 | 10.2 | 2.02  (1.44-2.84) | <0.001 |
|  | PostLD | 85.2 | 136 | 159 | 10.6 | 2.01  (1.26-3.19) | <0.001 |
| Edinburgh | 2019 | 68.1 | 3140 | 4611 | NA | NA | NA |
|  | PreLD | 68.1 | 709 | 1037 | 0 | 1.01  (0.88-1.17) | 0.86 |
|  | LD | 80.1 | 1315 | 1642 | 12 | 1.88  (1.64-2.16) | <0.001 |
|  | PostLD | 73.1 | 562 | 766 | 5 | 1.29  (1.09-1.53) | <0.001 |
| Falkirk | 2019 | 68.8 | 1052 | 1530 | NA | NA | NA |
|  | PreLD | 67.8 | 245 | 343 | -1 | 1.14  (0.88-1.47) | 0.33 |
|  | LD | 85.4 | 450 | 530 | 16.6 | 2.56  (1.97-3.32) | <0.001 |
|  | PostLD | 80.8 | 216 | 265 | 12 | 2  (1.44-2.78) | <0.001 |
| Fife | 2019 | 67 | 2379 | 3553 | NA | NA | NA |
|  | PreLD | 70.9 | 535 | 762 | 3.9 | 1.16  (0.98-1.38) | 0.08 |
|  | LD | 70.5 | 856 | 1214 | 3.5 | 1.18  (1.02-1.36) | 0.02 |
|  | PostLD | 68 | 414 | 608 | 1 | 1.05  (0.88-1.27) | 0.58 |
| Glasgow City | 2019 | 67 | 4209 | 6281 | NA | NA | NA |
|  | PreLD | 76.4 | 1079 | 1413 | 9.4 | 1.59  (1.39-1.82) | <0.001 |
|  | LD | 80.6 | 1942 | 2404 | 13.6 | 2.07  (1.85-2.32) | <0.001 |
|  | PostLD | 80.4 | 921 | 1146 | 13.4 | 2.02  (1.73-2.35) | <0.001 |
| Highland | 2019 | 56.7 | 1156 | 2039 | NA | NA | NA |
|  | PreLD | 55.1 | 254 | 434 | -1.6 | 1.08  (0.87-1.33) | 0.48 |
|  | LD | 68.5 | 511 | 746 | 11.8 | 1.66  (1.39-1.98) | <0.001 |
|  | PostLD | 58 | 216 | 371 | 1.3 | 1.06  (0.85-1.33) | 0.58 |
| Inverclyde | 2019 | 75.7 | 535 | 707 | NA | NA | NA |
|  | PreLD | 62.6 | 100 | 151 | -13.1 | 0.63  (0.43-0.92) | 0.02 |
|  | LD | 82 | 161 | 195 | 6.3 | 1.52  (1.01-2.29) | 0.04 |
|  | PostLD | 82.1 | 87 | 106 | 6.4 | 1.47  (0.87-2.49) | 0.15 |
| Midlothian | 2019 | 65.3 | 733 | 1122 | NA | NA | NA |
|  | PreLD | 64.8 | 177 | 258 | -0.5 | 1.16  (0.87-1.55) | 0.32 |
|  | LD | 82.8 | 349 | 421 | 17.5 | 2.57  (1.94-3.41) | <0.001 |
|  | PostLD | 83.5 | 163 | 196 | 18.2 | 2.62  (1.77-3.89) | <0.001 |
| Moray | 2019 | 54.1 | 447 | 827 | NA | NA | NA |
|  | PreLD | 62.7 | 111 | 181 | 8.6 | 1.35  (0.97-1.87) | 0.08 |
|  | LD | 86.4 | 264 | 308 | 32.3 | 5.1  (3.6-7.22) | <0.001 |
|  | PostLD | 86.3 | 130 | 150 | 32.2 | 5.53  (3.38-9.02) | <0.001 |
| North Ayrshire | 2019 | 56.5 | 648 | 1146 | NA | NA | NA |
|  | PreLD | 58.6 | 144 | 253 | 2.1 | 1.02  (0.77-1.34) | 0.91 |
|  | LD | 73 | 272 | 373 | 16.5 | 2.07  (1.6-2.67) | <0.001 |
|  | PostLD | 61.2 | 144 | 243 | 4.7 | 1.12  (0.84-1.48) | 0.44 |
| North Lanarkshire | 2019 | 69.8 | 2448 | 3509 | NA | NA | NA |
|  | PreLD | 68.3 | 566 | 791 | -1.5 | 1.09  (0.92-1.29) | 0.32 |
|  | LD | 83.4 | 1078 | 1286 | 13.6 | 2.25  (1.9-2.65) | <0.001 |
|  | PostLD | 78.4 | 488 | 624 | 8.6 | 1.56  (1.27-1.91) | <0.001 |
| Orkney Islands | 2019 | 53.9 | 111 | 206 | NA | NA | NA |
|  | PreLD | 55.7 | 19 | 33 | 1.8 | 1.16  (0.55-2.44) | 0.69 |
|  | LD | 73.2 | 50 | 67 | 19.3 | 2.8  (2.23-3.53) | <0.001 |
|  | PostLD | 71.6 | 29 | 41 | 17.7 | 1.89  (1.42-2.51) | <0.001 |
| Perth and Kinross | 2019 | 66.4 | 855 | 1287 | NA | NA | NA |
|  | PreLD | 65.6 | 177 | 263 | -0.8 | 1.04  (0.78-1.38) | 0.79 |
|  | LD | 78.2 | 351 | 448 | 11.8 | 1.83  (1.42-2.35) | <0.001 |
|  | PostLD | 72.7 | 156 | 217 | 6.3 | 1.29  (0.94-1.78) | 0.11 |
| Renfrewshire | 2019 | 69.1 | 1210 | 1750 | NA | NA | NA |
|  | PreLD | 73.2 | 301 | 400 | 4.1 | 1.36  (1.06-1.74) | 0.02 |
|  | LD | 83.5 | 507 | 607 | 14.4 | 2.26  (1.78-2.87) | <0.001 |
|  | PostLD | 74.9 | 216 | 287 | 5.8 | 1.36  (1.02-1.81) | 0.04 |
| Scottish Borders | 2019 | 67.1 | 658 | 981 | NA | NA | NA |
|  | PreLD | 66.9 | 134 | 204 | -0.2 | 0.94  (0.68-1.29) | 0.7 |
|  | LD | 78.7 | 250 | 317 | 11.6 | 1.83  (1.36-2.47) | <0.001 |
|  | PostLD | 70.8 | 114 | 154 | 3.7 | 1.4  (0.95-2.05) | 0.09 |
| Shetland Islands | 2019 | 29.8 | 68 | 228 | NA | NA | NA |
|  | PreLD | 54.1 | 19 | 35 | 24.3 | 2.79  (1.36-5.76) | 0.01 |
|  | LD | 39.5 | 27 | 69 | 9.7 | 2.8  (2.23-3.53) | <0.001 |
|  | PostLD | 24.5 | 12 | 41 | -5.3 | 1.89  (1.42-2.51) | <0.001 |
| South Ayrshire | 2019 | 61.5 | 583 | 948 | NA | NA | NA |
|  | PreLD | 70.8 | 128 | 182 | 9.3 | 1.48  (1.05-2.09) | 0.02 |
|  | LD | 81.4 | 274 | 333 | 19.9 | 2.91  (2.13-3.97) | <0.001 |
|  | PostLD | 78.6 | 132 | 169 | 17.1 | 2.23  (1.52-3.29) | <0.001 |
| South Lanarkshire | 2019 | 70.1 | 2271 | 3241 | NA | NA | NA |
|  | PreLD | 71.3 | 548 | 731 | 1.2 | 1.28  (1.06-1.54) | 0.01 |
|  | LD | 81.8 | 960 | 1171 | 11.7 | 1.94  (1.64-2.3) | <0.001 |
|  | PostLD | 81 | 460 | 567 | 10.9 | 1.84  (1.47-2.3) | <0.001 |
| West Dunbartonshire | 2019 | 65.8 | 573 | 871 | NA | NA | NA |
|  | PreLD | 75.6 | 136 | 185 | 9.8 | 1.44  (1.01-2.06) | 0.04 |
|  | LD | 79.4 | 249 | 311 | 13.6 | 2.09  (1.53-2.85) | <0.001 |
|  | PostLD | 78 | 128 | 164 | 12.2 | 1.85  (1.25-2.75) | <0.001 |
| West Lothian | 2019 | 66.7 | 1238 | 1857 | NA | NA | NA |
|  | PreLD | 70.1 | 301 | 418 | 3.4 | 1.29  (1.02-1.63) | 0.04 |
|  | LD | 84.6 | 583 | 687 | 17.9 | 2.8  (2.23-3.53) | <0.001 |
|  | PostLD | 79.1 | 257 | 325 | 12.4 | 1.89  (1.42-2.51) | <0.001 |
| Western Isles | 2019 | 52.6 | 113 | 215 | NA | NA | NA |
|  | PreLD | 64.6 | 24 | 38 | 12 | 1.55  (0.76-3.15) | 0.23 |
|  | LD | 77.1 | 55 | 72 | 24.5 | 1.87  (1.57-.22) | <0.001 |
|  | PostLD | 63.2 | 23 | 35 | 10.6 | 2.04  (1.62-2.57) | <0.001 |

**E Second dose MMR**

| **HSCP** | **Time period** | **% uptake (within 4 weeks)** | **Number received** | **Number eligible** | **% point change from 2019** | **OR compared to 2019**  **(95% CI)** | **p value** |
| --- | --- | --- | --- | --- | --- | --- | --- |
| Angus | 2019 | 43.8 | 498 | 1137 | NA | NA | NA |
|  | PreLD | 54.7 | 147 | 275 | 10.9 | 1.47  (1.13-1.92) | <0.001 |
|  | LD | 74.1 | 266 | 367 | 30.3 | 3.38  (2.61-4.37) | <0.001 |
|  | PostLD | 62.7 | 118 | 189 | 18.9 | 2.13  (1.55-2.93) | <0.001 |
| Argyll and Bute | 2019 | 57.3 | 436 | 761 | NA | NA | NA |
|  | PreLD | 55.8 | 92 | 166 | -1.5 | 0.93  (0.66-1.3) | 0.66 |
|  | LD | 61.2 | 169 | 279 | 3.9 | 1.15  (0.87-1.51) | 0.34 |
|  | PostLD | 60.8 | 65 | 108 | 3.5 | 1.13  (0.75-1.7) | 0.57 |
| Clackmannanshire and Stirling | 2019 | 52.3 | 770 | 1471 | NA | NA | NA |
|  | PreLD | 54.7 | 193 | 320 | 2.4 | 1.38  (1.08-1.77) | 0.01 |
|  | LD | 68.3 | 341 | 504 | 16 | 1.9  (1.54-2.36) | <0.001 |
|  | PostLD | 60.5 | 144 | 237 | 8.2 | 1.41  (1.07-1.87) | 0.02 |
| Dumfries and Galloway | 2019 | 63.2 | 871 | 1379 | NA | NA | NA |
|  | PreLD | 60.6 | 199 | 314 | -2.6 | 1.01  (0.78-1.3) | 0.94 |
|  | LD | 71 | 308 | 437 | 7.8 | 1.39  (1.1-1.76) | 0.01 |
|  | PostLD | 72.5 | 187 | 257 | 9.3 | 1.56  (1.16-2.09) | <0.001 |
| Dundee City | 2019 | 32.9 | 485 | 1476 | NA | NA | NA |
|  | PreLD | 39.4 | 143 | 346 | 6.5 | 1.44  (1.13-1.83) | <0.001 |
|  | LD | 58.9 | 309 | 523 | 26 | 2.95  (2.4-3.62) | <0.001 |
|  | PostLD | 52.8 | 137 | 256 | 19.9 | 2.35  (1.8-3.08) | <0.001 |
| East Ayrshire | 2019 | 39.7 | 522 | 1316 | NA | NA | NA |
|  | PreLD | 26.6 | 100 | 304 | -13.1 | 0.75  (0.57-0.97) | 0.03 |
|  | LD | 43.8 | 192 | 445 | 4.1 | 1.15  (0.93-1.44) | 0.2 |
|  | PostLD | 43.5 | 86 | 198 | 3.8 | 1.17  (0.86-1.58) | 0.31 |
| East Dunbartonshire | 2019 | 56.4 | 674 | 1196 | NA | NA | NA |
|  | PreLD | 63.6 | 197 | 300 | 7.2 | 1.48  (1.14-1.93) | <0.001 |
|  | LD | 82.6 | 370 | 447 | 26.2 | 3.72  (2.84-4.88) | <0.001 |
|  | PostLD | 85.1 | 188 | 223 | 28.7 | 4.16  (2.85-6.07) | <0.001 |
| East Lothian | 2019 | 60 | 762 | 1271 | NA | NA | NA |
|  | PreLD | 56.2 | 161 | 271 | -3.8 | 0.98  (0.75-1.28) | 0.87 |
|  | LD | 78.5 | 315 | 401 | 18.5 | 2.45  (1.88-3.18) | <0.001 |
|  | PostLD | 59.1 | 112 | 184 | -0.9 | 1.04  (0.76-1.43) | 0.81 |
| East Renfrewshire | 2019 | 62.5 | 731 | 1169 | NA | NA | NA |
|  | PreLD | 63.3 | 176 | 258 | 0.8 | 1.29  (0.96-1.71) | 0.09 |
|  | LD | 83.5 | 309 | 369 | 21 | 3.09  (2.28-4.17) | <0.001 |
|  | PostLD | 84.1 | 156 | 186 | 21.6 | 3.12  (2.07-4.69) | <0.001 |
| Edinburgh | 2019 | 56.3 | 2727 | 4846 | NA | NA | NA |
|  | PreLD | 60.5 | 660 | 1117 | 4.2 | 1.12  (0.98-1.28) | 0.09 |
|  | LD | 65.6 | 1103 | 1679 | 9.3 | 1.49  (1.33-1.67) | <0.001 |
|  | PostLD | 61.3 | 492 | 806 | 5 | 1.22  (1.05-1.42) | 0.01 |
| Falkirk | 2019 | 49 | 793 | 1619 | NA | NA | NA |
|  | PreLD | 47.6 | 194 | 366 | -1.4 | 1.17  (0.94-1.47) | 0.16 |
|  | LD | 62 | 353 | 579 | 13 | 1.63  (1.34-1.97) | <0.001 |
|  | PostLD | 62.8 | 178 | 284 | 13.8 | 1.75  (1.35-2.27) | <0.001 |
| Fife | 2019 | 47 | 1799 | 3826 | NA | NA | NA |
|  | PreLD | 50.4 | 446 | 906 | 3.4 | 1.09  (0.95-1.26) | 0.23 |
|  | LD | 52.2 | 673 | 1285 | 5.2 | 1.24  (1.09-1.41) | <0.001 |
|  | PostLD | 44.7 | 257 | 573 | -2.3 | 0.92  (0.77-1.09) | 0.33 |
| Glasgow City | 2019 | 50.9 | 3262 | 6411 | NA | NA | NA |
|  | PreLD | 61.8 | 960 | 1543 | 10.9 | 1.59  (1.42-1.78) | <0.001 |
|  | LD | 70.8 | 1682 | 2378 | 19.9 | 2.33  (2.11-2.58) | <0.001 |
|  | PostLD | 72.6 | 782 | 1077 | 21.7 | 2.56  (2.22-2.95) | <0.001 |
| Highland | 2019 | 50.7 | 1144 | 2257 | NA | NA | NA |
|  | PreLD | 31.8 | 223 | 516 | -18.9 | 0.74  (0.61-0.9) | <0.001 |
|  | LD | 58.4 | 474 | 803 | 7.7 | 1.4  (1.19-1.65) | <0.001 |
|  | PostLD | 52.3 | 192 | 356 | 1.6 | 1.14  (0.91-1.43) | 0.26 |
| Inverclyde | 2019 | 61.9 | 445 | 719 | NA | NA | NA |
|  | PreLD | 63.2 | 111 | 177 | 1.3 | 1.04  (0.74-1.45) | 0.84 |
|  | LD | 71.3 | 153 | 214 | 9.4 | 1.54  (1.11-2.15) | 0.01 |
|  | PostLD | 66.4 | 91 | 128 | 4.5 | 1.51  (1-2.28) | 0.05 |
| Midlothian | 2019 | 60.2 | 736 | 1223 | NA | NA | NA |
|  | PreLD | 56.8 | 168 | 293 | -3.4 | 0.89  (0.69-1.15) | 0.37 |
|  | LD | 75.8 | 299 | 405 | 15.6 | 1.87  (1.45-2.4) | <0.001 |
|  | PostLD | 68 | 137 | 204 | 7.8 | 1.35  (0.99-1.85) | 0.06 |
| North Ayrshire | 2019 | 39.4 | 526 | 1334 | NA | NA | NA |
|  | PreLD | 35.4 | 100 | 280 | -4 | 0.85  (0.65-1.12) | 0.25 |
|  | LD | 47.7 | 205 | 427 | 8.3 | 1.42  (1.14-1.77) | <0.001 |
|  | PostLD | 42.7 | 94 | 224 | 3.3 | 1.11  (0.83-1.48) | 0.47 |
| North Lanarkshire | 2019 | 53 | 1986 | 3749 | NA | NA | NA |
|  | PreLD | 53.1 | 508 | 845 | 0.1 | 1.34  (1.15-1.56) | <0.001 |
|  | LD | 70.7 | 888 | 1252 | 17.7 | 2.17  (1.89-2.49) | <0.001 |
|  | PostLD | 66.7 | 436 | 656 | 13.7 | 1.76  (1.48-2.09) | <0.001 |
| Orkney Islands | 2019 | 37 | 71 | 192 | NA | NA | NA |
|  | PreLD | 50.6 | 16 | 33 | 13.6 | 1.6  (0.76-3.37) | 0.21 |
|  | LD | 63.6 | 43 | 66 | 26.6 | 2.46  (2.02-2.99) | <0.001 |
|  | PostLD | 77.3 | 24 | 31 | 40.3 | 1.6  (1.25-2.04) | <0.001 |
| Perth and Kinross | 2019 | 45.5 | 654 | 1436 | NA | NA | NA |
|  | PreLD | 51.5 | 190 | 356 | 6 | 1.37  (1.08-1.73) | 0.01 |
|  | LD | 63.2 | 284 | 465 | 17.7 | 1.88  (1.52-2.32) | <0.001 |
|  | PostLD | 59.1 | 121 | 203 | 13.6 | 1.76  (1.31-2.38) | <0.001 |
| Renfrewshire | 2019 | 57.5 | 1064 | 1852 | NA | NA | NA |
|  | PreLD | 60.9 | 252 | 408 | 3.4 | 1.2  (0.96-1.49) | 0.11 |
|  | LD | 70.6 | 445 | 627 | 13.1 | 1.81  (1.49-2.2) | <0.001 |
|  | PostLD | 68.8 | 215 | 310 | 11.3 | 1.68  (1.29-2.17) | <0.001 |
| Scottish Borders | 2019 | 55.5 | 616 | 1110 | NA | NA | NA |
|  | PreLD | 51.9 | 122 | 224 | -3.6 | 0.96  (0.72-1.28) | 0.78 |
|  | LD | 64.6 | 218 | 335 | 9.1 | 1.49  (1.16-1.93) | <0.001 |
|  | PostLD | 59.1 | 111 | 189 | 3.6 | 1.14  (0.83-1.56) | 0.41 |
| Shetland Islands | 2019 | 24.5 | 68 | 278 | NA | NA | NA |
|  | PreLD | 38 | 16 | 42 | 13.5 | 1.9  (0.96-3.75) | 0.06 |
|  | LD | 19 | 16 | 70 | -5.5 | 2.46  (2.02-2.99) | <0.001 |
|  | PostLD | 19.9 | 9 | 44 | -4.6 | 1.6  (1.25-2.04) | <0.001 |
| South Ayrshire | 2019 | 42.6 | 447 | 1050 | NA | NA | NA |
|  | PreLD | 37.3 | 100 | 244 | -5.3 | 0.94  (0.71-1.24) | 0.65 |
|  | LD | 54.6 | 187 | 354 | 12 | 1.51  (1.19-1.92) | <0.001 |
|  | PostLD | 56.4 | 94 | 166 | 13.8 | 1.76  (1.27-2.45) | <0.001 |
| South Lanarkshire | 2019 | 53.3 | 1839 | 3452 | NA | NA | NA |
|  | PreLD | 52 | 453 | 798 | -1.3 | 1.15  (0.99-1.35) | 0.07 |
|  | LD | 67.7 | 844 | 1247 | 14.4 | 1.84  (1.6-2.1) | <0.001 |
|  | PostLD | 64.5 | 364 | 563 | 11.2 | 1.6  (1.33-1.93) | <0.001 |
| West Dunbartonshire | 2019 | 53 | 529 | 998 | NA | NA | NA |
|  | PreLD | 62 | 125 | 200 | 9 | 1.48  (1.08-2.02) | 0.01 |
|  | LD | 68.4 | 203 | 294 | 15.4 | 1.98  (1.5-2.61) | <0.001 |
|  | PostLD | 71 | 111 | 152 | 18 | 2.4  (1.64-3.51) | <0.001 |
| West Lothian | 2019 | 59.3 | 1238 | 2089 | NA | NA | NA |
|  | PreLD | 59.3 | 291 | 493 | 0 | 0.99  (0.81-1.21) | 0.9 |
|  | LD | 79 | 576 | 737 | 19.7 | 2.46  (2.02-2.99) | <0.001 |
|  | PostLD | 69.9 | 239 | 342 | 10.6 | 1.6  (1.25-2.04) | <0.001 |
| Western Isles | 2019 | 45.6 | 104 | 228 | NA | NA | NA |
|  | PreLD | 41.4 | 20 | 48 | -4.2 | 0.85  (0.45-1.6) | 0.62 |
|  | LD | 47.8 | 43 | 89 | 2.1 | 3.38  (2.61-4.37) | <0.001 |
|  | PostLD | 64.1 | 27 | 42 | 18.5 | 2.13  (1.55-2.93) | <0.001 |

**Supplementary Table S3**

| **Immunisation** | **Deprivation quintile** | **Time period** | **% received within 4 weeks**  **(Number received/total eligible)** | **% point change from 2019** | **OR (95%CI) for uptake compared to 2019** | **p-value** |
| --- | --- | --- | --- | --- | --- | --- |
| **First 6in1** | 1 - most deprived | 2019 | 91.9  (11025/11996) | NA | NA | NA |
|  | 1 - most deprived | PreLD | 91.5  (2383/2580) | -0.4 | 1.1 (0.9-1.3) | 0.44 |
|  | 1 - most deprived | LD | 93.9  (3815/4067) | 2 | 1.3 (1.2-1.5) | <0.001 |
|  | 1 - most deprived | PostLD | 94  (1906/2026) | 2.1 | 1.4 (1.2-1.7) | <0.001 |
|  | 2 | 2019 | 93.1  (9740/10461) | NA | NA | NA |
|  | 2 | PreLD | 91.6  (2119/2289) | -1.5 | 0.9 (0.8-1.1) | 0.36 |
|  | 2 | LD | 95.1  (3311/3484) | 2 | 1.4 (1.2-1.7) | <0.001 |
|  | 2 | PostLD | 93.9  (1641/1746) | 0.8 | 1.2 (0.9-1.4) | 0.18 |
|  | 3 | 2019 | 93.7  (8457/9030) | NA | NA | NA |
|  | 3 | PreLD | 93.1  (1814/1931) | -0.6 | 1.1 (0.9-1.3) | 0.64 |
|  | 3 | LD | 95.2  (3070/3224) | 1.5 | 1.4 (1.1-1.6) | <0.001 |
|  | 3 | PostLD | 95  (1468/1546) | 1.3 | 1.3 (1-1.6) | 0.05 |
|  | 4 | 2019 | 95.6  (9804/10251) | NA | NA | NA |
|  | 4 | PreLD | 95.3  (2038/2142) | -0.3 | 0.9 (0.7-1.1) | 0.31 |
|  | 4 | LD | 95.9  (3328/3470) | 0.3 | 1.1 (0.9-1.3) | 0.5 |
|  | 4 | PostLD | 94.4  (1677/1776) | -1.2 | 0.8 (0.6-1) | 0.02 |
|  | 5 - least deprived | 2019 | 96.3  (8442/8769) | NA | NA | NA |
|  | 5 - least deprived | PreLD | 95.7  (1724/1794) | -0.6 | 1(0.7-1.3) | 0.73 |
|  | 5 - least deprived | LD | 96.7  (2759/2853) | 0.4 | 1.1 (0.9-1.4) | 0.28 |
|  | 5 - least deprived | PostLD | 96.3  (1373/1427) | 0 | 1 (0.7-1.3) | 0.92 |
| **Second 6in1** | 1 - most deprived | 2019 | 79.8  (9633/12078) | NA | NA | NA |
|  | 1 - most deprived | PreLD | 78  (1984/2492) | -1.8 | 1(0.9-1.1) | 0.87 |
|  | 1 - most deprived | LD | 86.3  (3533/4096) | 6.5 | 1.6 (1.4-1.8) | <0.001 |
|  | 1 - most deprived | PostLD | 86.3  (1737/2013) | 6.5 | 1.6 (1.4-1.8) | <0.001 |
|  | 2 | 2019 | 82.9  (8702/10499) | NA | NA | NA |
|  | 2 | PreLD | 83.4  (1880/2256) | 0.5 | 1 (0.9-1.2) | 0.61 |
|  | 2 | LD | 88.4  (3133/3549) | 5.5 | 1.6 (1.4-1.7) | <0.001 |
|  | 2 | PostLD | 86.9  (1509/1736) | 4 | 1.4 (1.2-1.6) | <0.001 |
|  | 3 | 2019 | 85.2  (7711/9055) | NA | NA | NA |
|  | 3 | PreLD | 85.4  (1675/1968) | 0.2 | 1 (0.9-1.1) | 0.96 |
|  | 3 | LD | 89.5  (2851/3182) | 4.3 | 1.5 (1.3-1.7) | <0.001 |
|  | 3 | PostLD | 89.4  (1387/1553) | 4.2 | 1.5 (1.2-1.7) | <0.001 |
|  | 4 | 2019 | 88.5  (9157/10348) | NA | NA | NA |
|  | 4 | PreLD | 89  (1892/2121) | 0.5 | 1.1 (0.9-1.3) | 0.35 |
|  | 4 | LD | 92.2  (3221/3494) | 3.7 | 1.5 (1.3-1.8) | <0.001 |
|  | 4 | PostLD | 90.2  (1540/1708) | 1.7 | 1.2 (1-1.4) | 0.04 |
|  | 5 - least deprived | 2019 | 89.1  (7925/8890) | NA | NA | NA |
|  | 5 - least deprived | PreLD | 88.5  (1655/1837) | -0.6 | 1.1 (0.9-1.3) | 0.23 |
|  | 5 - least deprived | LD | 93.3  (2668/2862) | 4.2 | 1.7 (1.4-2) | <0.001 |
|  | 5 - least deprived | PostLD | 91.7  (1280/1395) | 2.6 | 1.4 (1.1-1.7) | <0.001 |
| **Third 6in1** | 1 - most deprived | 2019 | 66.2  (8007/12102) | NA | NA | NA |
|  | 1 - most deprived | PreLD | 65.5  (1727/2640) | -0.7 | 1 (0.9-1.1) | 0.46 |
|  | 1 - most deprived | LD | 77.1  (3175/4114) | 10.9 | 1.7 (1.6-1.9) | <0.001 |
|  | 1 - most deprived | PostLD | 74.9  (1427/1905) | 8.7 | 1.5 (1.4-1.7) | <0.001 |
|  | 2 | 2019 | 70.3  (7432/10569) | NA | NA | NA |
|  | 2 | PreLD | 68.5  (1607/2331) | -1.8 | 0.9 (0.9-1) | 0.19 |
|  | 2 | LD | 80  (2836/3550) | 9.7 | 1.7 (1.5-1.8) | <0.001 |
|  | 2 | PostLD | 79.4  (1336/1683) | 9.1 | 1.6 (1.4-1.8) | <0.001 |
|  | 3 | 2019 | 73.2  (6670/9107) | NA | NA | NA |
|  | 3 | PreLD | 74.3  (1499/2045) | 1.1 | 1 (0.9-1.1) | 0.96 |
|  | 3 | LD | 82  (2610/3178) | 8.8 | 1.7 (1.5-1.9) | <0.001 |
|  | 3 | PostLD | 80.6  (1242/1542) | 7.4 | 1.5 (1.3-1.7) | <0.001 |
|  | 4 | 2019 | 77.5  (7991/10307) | NA | NA | NA |
|  | 4 | PreLD | 77.5  (1799/2322) | 0 | 1 (0.9-1.1) | 0.96 |
|  | 4 | LD | 86.2  (2911/3376) | 8.7 | 1.8 (1.6-2) | <0.001 |
|  | 4 | PostLD | 82.7  (1414/1709) | 5.2 | 1.4 (1.2-1.6) | <0.001 |
|  | 5 - least deprived | 2019 | 79.7  (7091/8893) | NA | NA | NA |
|  | 5 - least deprived | PreLD | 80.9  (1626/2032) | 1.2 | 1 (0.9-1.1) | 0.77 |
|  | 5 - least deprived | LD | 87  (2478/2841) | 7.3 | 1.7 (1.5-2) | <0.001 |
|  | 5 - least deprived | PostLD | 85.2  (1121/1316) | 5.5 | 1.5 (1.2-1.7) | <0.001 |
| **First MMR** | 1 - most deprived | 2019 | 62.4  (7561/12121) | NA | NA | NA |
|  | 1 - most deprived | PreLD | 63.8  (1790/2721) | 1.4 | 1.2 (1.1-1.3) | <0.001 |
|  | 1 - most deprived | LD | 73.7  3217/4358) | 11.3 | 1.7 (1.6-1.8) | <0.001 |
|  | 1 - most deprived | PostLD | 70.6  (1529/2168) | 8.2 | 1.4 (1.3-1.6) | <0.001 |
|  | 2 | 2019 | 63.8  (6720/10533) | NA | NA | NA |
|  | 2 | PreLD | 64.6  (1579/2378) | 0.8 | 1.1 (1-1.2) | 0.02 |
|  | 2 | LD | 76.2  (2920/3825) | 12.4 | 1.8 (1.7-2) | <0.001 |
|  | 2 | PostLD | 72.9  (1325/1814) | 9.1 | 1.5 (1.4-1.7) | <0.001 |
|  | 3 | 2019 | 64  (5936/9281) | NA | NA | NA |
|  | 3 | PreLD | 68.1  (1376/2024) | 4.1 | 1.2 (1.1-1.3) | <0.001 |
|  | 3 | LD | 77.9  (2541/3251) | 13.9 | 2 (1.8-2.2) | <0.001 |
|  | 3 | PostLD | 74.6  (1260/1685) | 10.6 | 1.7 (1.5-1.9) | <0.001 |
|  | 4 | 2019 | 66.5  (7102/10673) | NA | NA | NA |
|  | 4 | PreLD | 67.9  (1597/2292) | 1.4 | 1.2 (1-1.3) | <0.001 |
|  | 4 | LD | 80.5  (3041/3776) | 14 | 2.1 (1.9-2.3) | <0.001 |
|  | 4 | PostLD | 76.6  (1379/1801) | 10.1 | 1.6 (1.5-1.8) | <0.001 |
|  | 5 - least deprived | 2019 | 70.4  (6563/9325) | NA | NA | NA |
|  | 5 - least deprived | PreLD | 74  (1423/1927) | 3.6 | 1.2 (1.1-1.3) | <0.001 |
|  | 5 - least deprived | LD | 85.1  (2730/3208) | 14.7 | 2.4 (2.2-2.7) | <0.001 |
|  | 5 - least deprived | PostLD | 79  (1238/1566) | 8.6 | 1.6 (1.4-1.8) | <0.001 |
| **Second MMR** | 1 - most deprived | 2019 | 46  (5785/12580) | NA | NA | NA |
|  | 1 - most deprived | PreLD | 47.9  (1482/2935) | 1.9 | 1.2 (1.1-1.3) | <0.001 |
|  | 1 - most deprived | LD | 60.3  (2727/4538) | 14.3 | 1.8 (1.7-1.9) | <0.001 |
|  | 1 - most deprived | PostLD | 57.7  (1236/2139) | 11.7 | 1.6 (1.5-1.8) | <0.001 |
|  | 2 | 2019 | 50.2  (5169/10291) | NA | NA | NA |
|  | 2 | PreLD | 53.3  (1270/2300) | 3.1 | 1.2 (1.1-1.3) | <0.001 |
|  | 2 | LD | 63.7  (2276/3570) | 13.5 | 1.7 (1.6-1.9) | <0.001 |
|  | 2 | PostLD | 60.3  (1001/1661) | 10.1 | 1.5 (1.4-1.7) | <0.001 |
|  | 3 | 2019 | 51.7  (4701/9100) | NA | NA | NA |
|  | 3 | PreLD | 51.8  (1127/2099) | 0.1 | 1.2 (1.1-1.3) | <0.001 |
|  | 3 | LD | 64.5  (1937/3007) | 12.8 | 2 (1.8-2.2) | <0.001 |
|  | 3 | PostLD | 62.3  (916/1470) | 10.6 | 1.7 (1.5-1.9) | <0.001 |
|  | 4 | 2019 | 54.2  (4998/9219) | NA | NA | NA |
|  | 4 | PreLD | 53.3  (1219/2135) | -0.9 | 1.1 (1-1.2) | 0.02 |
|  | 4 | LD | 69.8  (2238/3211) | 15.6 | 1.9 (1.8-2.1) | <0.001 |
|  | 4 | PostLD | 66.4  (1014/1528) | 12.2 | 1.7 (1.5-1.9) | <0.001 |
|  | 5 - least deprived | 2019 | 59.4  (5148/8670) | NA | NA | NA |
|  | 5 - least deprived | PreLD | 61.6  (1281/2010) | 2.2 | 1.2 (1.1-1.3) | <0.001 |
|  | 5 - least deprived | LD | 75.6  (2105/2787) | 16.2 | 2.1 (1.9-2.3) | <0.001 |
|  | 5 - least deprived | PostLD | 72  (1000/1391) | 12.6 | 1.7 (1.5-2) | <0.001 |

Table S3: Uptake of pre-school immunisations by time period and SIMD and percent point change in uptake compared to baseline 2019. Odds ratio and 95% confidence intervals shown are for change in uptake compared to 2019. *p*-value rounded to 2 decimal places. LD = lockdown, NA = not applicable. Statistically significant change in uptake compared to 2019 are shaded green.

**Supplementary Table S4**

| **Immunisation** | **Interaction term**  **(baseline comparisons = 2019, SIMD 1)** | **ROR**  **(exp of coeff of interaction model)** | **95% Confidence intervals** | ***p* -value** |
| --- | --- | --- | --- | --- |
| **First 6in1** | PreLD:SIMD2 | 0.87 | (0.68-1.1)  ns | 0.23 |
|  | LD:SIMD2 | 1.06 | (0.85-1.33)  ns | 0.59 |
|  | PostLD:SIMD2 | 0.83 | (0.62-1.1)  ns | 0.2 |
|  | PreLD:SIMD3 | 0.99 | (0.76-1.28)  ns | 0.92 |
|  | LD:SIMD3 | 1.01 | (0.8-1.28)  ns | 0.91 |
|  | PostLD:SIMD3 | 0.91 | (0.67-1.25)  ns | 0.56 |
|  | PreLD:SIMD4 | 0.84 | (0.64-1.1)  ns | 0.2 |
|  | LD:SIMD4 | 0.8 | (0.63-1.02)  ns | 0.07 |
|  | PostLD:SIMD4 | 0.55 | (0.41-0.74)  sig | <0.001 |
|  | PreLD:SIMD5 | 0.9 | (0.66-1.22)  ns | 0.48 |
|  | LD:SIMD5 | 0.85 | (0.65-1.12)  ns | 0.25 |
|  | PostLD:SIMD5 | 0.7 | (0.5-1.01)  ns | 0.05 |
| **Second 6in1** | PreLD:SIMD2 | 1.04 | (0.89-1.23)  ns | 0.62 |
|  | LD:SIMD2 | 0.98 | (0.84-1.14)  ns | 0.76 |
|  | PostLD:SIMD2 | 0.86 | (0.7-1.05)  ns | 0.14 |
|  | PreLD:SIMD3 | 1.01 | (0.85-1.2)  ns | 0.95 |
|  | LD:SIMD3 | 0.94 | (0.8-1.11)  ns | 0.47 |
|  | PostLD:SIMD3 | 0.91 | (0.73-1.13)  ns | 0.41 |
|  | PreLD:SIMD4 | 1.08 | (0.9-1.3)  ns | 0.39 |
|  | LD:SIMD4 | 0.96 | (0.81-1.14)  ns | 0.67 |
|  | PostLD:SIMD4 | 0.75 | (0.6-0.93)  sig | 0.01 |
|  | PreLD:SIMD5 | 1.12 | (0.92-1.36)  ns | 0.27 |
|  | LD:SIMD5 | 1.05 | (0.87-1.27)  ns | 0.6 |
|  | PostLD:SIMD5 | 0.85 | (0.67-1.08)  ns | 0.18 |
| **Third 6in1** | PreLD:SIMD2 | 0.97 | (0.85-1.1)  ns | 0.63 |
|  | LD:SIMD2 | 0.97 | (0.86-1.1)  ns | 0.62 |
|  | PostLD:SIMD2 | 1.06 | (0.9-1.26)  ns | 0.46 |
|  | PreLD:SIMD3 | 1.04 | (0.9-1.19)  ns | 0.61 |
|  | LD:SIMD3 | 0.97 | (0.85-1.11)  ns | 0.66 |
|  | PostLD:SIMD3 | 0.99 | (0.83-1.18)  ns | 0.92 |
|  | PreLD:SIMD4 | 1.03 | (0.9-1.19)  ns | 0.67 |
|  | LD:SIMD4 | 1.05 | (0.92-1.2)  ns | 0.49 |
|  | PostLD:SIMD4 | 0.91 | (0.77-1.08)  ns | 0.29 |
|  | PreLD:SIMD5 | 1.05 | (0.91-1.22)  ns | 0.51 |
|  | LD:SIMD5 | 1 | (0.87-1.16)  ns | 0.97 |
|  | PostLD:SIMD5 | 0.96 | (0.79-1.16)  ns | 0.66 |
| **First MMR** | PreLD:SIMD2 | 0.97 | (0.85-1.1)  ns | 0.61 |
|  | LD:SIMD2 | 1.08 | (0.96-1.21)  ns | 0.21 |
|  | PostLD:SIMD2 | 1.07 | (0.92-1.24)  ns | 0.4 |
|  | PreLD:SIMD3 | 1.03 | (0.9-1.18)  ns | 0.65 |
|  | LD:SIMD3 | 1.19 | (1.05-1.34)  sig | 0.01 |
|  | PostLD:SIMD3 | 1.16 | (0.99-1.35)  ns | 0.06 |
|  | PreLD:SIMD4 | 1 | (0.87-1.14)  ns | 0.96 |
|  | LD:SIMD4 | 1.22 | (1.09-1.38)  sig | <0.001 |
|  | PostLD:SIMD4 | 1.14 | (0.98-1.33)  ns | 0.1 |
|  | PreLD:SIMD5 | 1.02 | (0.89-1.18)  ns | 0.73 |
|  | LD:SIMD5 | 1.41 | (1.24-1.61)  sig | <0.001 |
|  | PostLD:SIMD5 | 1.1 | (0.94-1.3)  ns | 0.25 |
| **Second MMR** | PreLD:SIMD2 | 1.02 | (0.9-1.15)  ns | 0.75 |
|  | LD:SIMD2 | 0.99 | (0.89-1.09)  ns | 0.78 |
|  | PostLD:SIMD2 | 0.93 | (0.81-1.08)  ns | 0.35 |
|  | PreLD:SIMD3 | 0.91 | (0.8-1.03)  ns | 0.12 |
|  | LD:SIMD3 | 0.96 | (0.86-1.07)  ns | 0.44 |
|  | PostLD:SIMD3 | 0.96 | (0.83-1.11)  ns | 0.61 |
|  | PreLD:SIMD4 | 0.94 | (0.83-1.06)  ns | 0.31 |
|  | LD:SIMD4 | 1.1 | (0.98-1.23)  ns | 0.09 |
|  | PostLD:SIMD4 | 1.04 | (0.9-1.2)  ns | 0.63 |
|  | PreLD:SIMD5 | 1 | (0.88-1.14)  ns | 0.96 |
|  | LD:SIMD5 | 1.19 | (1.06-1.34)  sig | <0.001 |
|  | PostLD:SIMD5 | 1.09 | (0.93-1.27)  ns | 0.29 |

Table S4: To assess whether the differences between change in uptake were statistically significant between SIMD quintiles, the interaction between time period and SIMD quintile was added into the model. The baseline comparisons showed are for time period 2019 and deprivation quintile SIMD 1. ROR = ratio of odds ratio, calculated by taking the exponential function of the coefficient of the interaction term from the interaction model. If the 95% confidence intervals did not include 1, the interaction of time period and SIMD was considered statistically significant, that is; there was a significant difference in the level of change (2019- time period) between the deprivation quintile and SIMD 1. For example, the increase in uptake during lockdown for SIMD 5 was statistically greater than the increase in uptake for SIMD 1. The ROR can be used to calculate the odds ratio for uptake compared to the baseline levels by multiplying the ROR with the relevant OR in table S3. LD= lockdown, SIMD = Scottish Index of Multiple Deprivation, ns = not statistically significant (coloured green) = interaction was statistically significant. *p*-value rounded to 2 decimal places.

**Appendix 4**

**Supplementary table S5**

| **Immunisation** | **Time period** | **% uptake**  **(no received/no eligible)** | **% point change from 2019** | **OR for uptake compared to 2019**  **(95% CI)** | ***p*-value** |
| --- | --- | --- | --- | --- | --- |
| First6in1  (uptake by age 24weeks) | 2019 | 97.9  (49542/50609) | NA | NA | NA |
|  | Pre LD | 97.7  (10514/10761) | -0.2 | 0.92  (0.8-1.06) | 0.22 |
|  | LD | 97.6  (16724/17133) | -0.3 | 0.88  (0.79-0.99) | 0.03 |
|  | Post LD | 97.5  (8319/8531) | -0.4 | 0.85  (0.73-0.98) | 0.03 |
| Second6in1  (uptake by age 28 weeks) | 2019 | 96.7  (49291/50975) | NA | NA | NA |
|  | Pre LD | 96.2  (10306/10698) | -0.5 | 0.9  (0.8-1.01) | 0.06 |
|  | LD | 96.6  (16639/17222) | -0.1 | 0.98  (0.89-1.07) | 0.61 |
|  | Post LD | 96.6  (8125/8412) | -0.1 | 0.97  (0.85-1.1) | 0.61 |
| Third6in1  (uptake by age 32 weeks) | 2019 | 94  (48029/51085) | NA | NA | NA |
|  | Pre LD | 94.1  (10728/11394) | 0.1 | 1.02  (0.94-1.12) | 0.59 |
|  | LD | 94.8  (16199/17093) | 0.8 | 1.15  (1.07-1.24) | <0.001 |
|  | Post LD | 93.5  (7644/8172) | -0.5 | 0.92  (0.84-1.01) | 0.09 |
| FirstMMR  (uptake by age 16 months) | 2019 | 91.1  (47386/52015) | NA | NA | NA |
|  | Pre LD | 91.3  (10389/11370) | 0.2 | 1.03  (0.96-1.11) | 0.36 |
|  | LD | 92.5  (17076/18463) | 1.4 | 1.2  (1.13-1.28) | <0.001 |
|  | Post LD | 91.6  (8285/9047) | 0.5 | 1.06  (0.98-1.15) | 0.14 |
| SecondMMR  (uptake by age 3years 8 months) | 2019 | 80.8  (40376/49940) | NA | NA | NA |
|  | Pre LD | 83.2  (9471/11495) | 2.4 | 1.11  (1.05-1.17) | <0.001 |
|  | LD | 86.1  (14763/17145) | 5.3 | 1.47  (1.4-1.54) | <0.001 |
|  | Post LD | 84.4  (6915/8196) | 3.6 | 1.28  (1.2-1.36) | <0.001 |

Table S5: Scotland. Uptake of pre-school immunisations at an older age by time period and point percentage change from 2019 with odds ratio and 95% confidence intervals compared to baseline of 2019. Children are categorised into the time-period at which they became eligible for the immunisation as before and uptake data were extracted at a later stage when they reached the ages indicated in the immunisation column. LD = lockdown, NA = not applicable. Statistically significant changes are coloured green.

**Supplementary table S6**

| **Immunisation** | **Time period** | **% uptake**  **(no received/no eligible)** | **% point change from 2019** | **OR for uptake compared to 2019**  **(95% CI)** | ***p*-value** |
| --- | --- | --- | --- | --- | --- |
| First6in1  (uptake by age 6 months) | 2019 | 96.3  (571531/593700) | NA | NA | NA |
|  | Pre LD | 95.8  (132779/138608) | 0.5 | 0.88 (0.86-0.91) | <0.001 |
|  | LD | 95.8  (171660/179180) | 0.5 | 0.89 (0.86-0.91) | <0.001 |
|  | Post LD | 95.6  (90702/94862) | 0.7 | 0.85 (0.82 -0.87) | <0.001 |
| Second6in1  (uptake by age 6 months) | 2019 | 93.9  (559382/595815) | NA | NA | NA |
|  | Pre LD | 92.1  (131578/142833) | 1.8 | 0.76 (0.74-0.78) | <0.001 |
|  | LD | 93.0  (164780/177221) | 0.9 | 0.86 (0.84-0.88) | <0.001 |
|  | Post LD | 92.5  (87818/94960) | 1.4 | 0.80 (0.78-0.82) | <0.001 |
| Third6in1  (uptake by age 6 months) | 2019 | 88.7  (529163/596355) | NA | NA | NA |
|  | Pre LD | 85.6  (126284/147479) | 3.1 | 0.76 (0.74-0.77) | <0.001 |
|  | LD | 86.6  (154912/178953) | 2.1 | 0.82 (0.81-0.83) | <0.001 |
|  | Post LD | 86.0  (79991/92990) | 2.7 | 0.78 (0.77-0.80) | <0.001 |
| FirstMMR  (uptake by age 18 months) | 2019 | 88.1  (541107/613923) | NA | NA | NA |
|  | Pre LD | 86.7  (119885/138353) | 1.4 | 0.87(0.86-0.89) | <0.001 |
|  | LD | 86.2  (173451/201164) | 1.9 | 0.84(0.83-0.85) | <0.001 |
|  | Post LD | 86.2  (85939/99685) | 1.9 | 0.84(0.83-0.86) | <0.001 |

Table S6: England. Uptake of pre-school immunisations at an older age by time period and point percentage change from 2019 with odds ratio and 95% confidence intervals compared to baseline of 2019. Children are categorised into the time-period at which they became eligible for the immunisation as before and uptake data were extracted at a later stage when they reached the ages indicated in the immunisation column. LD = lockdown, NA = not applicable. Statistically significant changes are coloured green.
